# Costs and Benefits of Malaria Elimination in Kenya by Means of IVM Implementation

**DOI:** 10.64898/2026.06.19.26355954

**Authors:** Santiago Movilla Blanco

## Abstract

1

**Introduction:** Integrated Vector Management (IVM) (Beier et al., 2008) is considered to be one of the most cost-effective tools to reduce malaria transmission. This rational decision-making tool supports the design of an intelligent and optimal management of resources meant for malaria prevention and vector control. Some of the most widespread components of IVM include the distribution of bed nets, spraying operations and environmental management to reduce mosquito density.

IVM was launched in Kenya in 2001, and during the last two decades the country has advocated for the use of this tool to combat malaria. IVM, however, requires a customized choice of interventions in order to achieve the optimal use of resources for vector control in the area of interest.

**Methods:** In this article, we present a malaria simulation model that helps us assess the potential impact of different future IVM strategies to achieve optimal malaria reduction results in Kenya.

The model used for this analysis is inspired by the Malaria Management Model (MMM) (Pedercini, Movilla Blanco, & Kopainsky, 2011), which has been developed by means of the System Dynamics methodology. The version of the model presented in this paper is adapted for Kenya, and the study includes the assessment of the cost and main socio-economic benefits from eliminating malaria in the country. The model also integrates the main impacts of malaria transmission into the socio-economic development, evaluating the effect of malaria on GDP production, literacy rate and life expectancy, among others.

**Results:** Our model includes data collected from 1980 until 2018, before the RTS,S/AS01 vaccine was piloted in Kenya, and the simulation results confirm that the IVM programs in Kenya would not achieve malaria elimination within this decade. In addition to a substantial increase in the malaria budget, a reallocation of such a budget across interventions would be necessary in order to reach a malaria-free country by 2030 (Kenya Malaria Strategic Plan).

The results also confirm that, despite the need for further efforts, malaria reduction in Kenya has provided sufficient economic benefits to cover the costs of malaria control.

**Conclusions:** The System Dynamics simulation model illustrates how a computer-aided scenario analysis tool can inform the design of malaria control policies and assist decision-makers. Simulation models create systematic mechanisms for analyzing alternative interventions and inform about the different tradeoffs.

## 2. Introduction

Malaria is one of the world’s most deadly diseases, and it is especially dangerous for pregnant women and children under 5 years of age. Apart of the consequences in mortality and the subsequent reduction in life expectancy, malaria affects people’s physical conditions, making them more vulnerable to other diseases.

As a result, malaria leads to anemia, malnutrition and many other health problems that also affect the number of hours worked per day by the labor force in a country, and may thus have an impact on their productivity, thus reducing a country’s economic growth prospects. In addition, malaria reduces students’ attendance at school, affecting their education and productivity in the long run. Malaria prevention and treatment also absorb a large amount of funds that could otherwise be used for investment in productive activities.

Kenya is one of the sub-Saharan countries where malaria is still endemic in some of its regions. In collaboration with partners, the Government of Kenya (GoK) has made substantial efforts to control malaria transmission in the whole country, reaching significant reductions in malaria prevalence (population fraction with malaria at any given point in time), especially along the coast (Kibe et al., 2006) [https](https://pubmed.ncbi.nlm.nih.gov/17604514/).

Interventions to reduce malaria prevalence and transmission focus on both, case management and prevention. The first type of intervention deals with diagnosis and treatment of the disease. Case management is especially effective to control malaria in low prevalence areas, although is a necessary measure in any malaria risk area. The second type of intervention is preventive, and includes very diverse methods, all of them aiming at reducing bites from Anopheles mosquitoes (the vector of malaria transmission) to humans. Plasmodium falciparum is the species most frequently associated with severe malaria and accounts for 80-90% of cases in Kenya. During the last two decades, prevention in the country has been mostly based on bed net distribution and insecticide indoor spraying operations. But other interventions such as environmental management, larviciding, or sensitization^1^ have gained momentum (National Malaria Control Programme - NMCP/Kenya, Kenya National Bureau of Statistics - KNBS, & ICF International, 2016) [https](https://dhsprogram.com/pubs/pdf/MIS22/MIS22.pdf).

As no intervention, in isolation, is able to control malaria transmission and, given that mosquitoes adapt to some interventions, integration and coordination of various prevention methods is essential in order to reach malaria elimination. In addition, one important lesson of the first Global Malaria Eradication effort, is that elimination cannot be achieved everywhere with the same interventions within the same time frame (Snow et al., 2015) [https](https://pmc.ncbi.nlm.nih.gov/articles/PMC4479373/).

In this regard, the Government of Kenya is advocating for the use of Integrated Vector Management (IVM), which is a rational decision-making tool designed to provide intelligent and optimal management of resources meant for malaria prevention and vector control (Beier et al., 2008) [https](https://www.who.int/malaria/vector_control/ivm/en/). Apart from achieving optimal use of existing resources to fight malaria, IVM also aims at increasing social mobilization and capacity building (Beier et al., 2008).

In this context, during the last two decades the GoK has significantly reduced malaria transmissions in the country through IVM implementation, achieving states of pre-elimination or even elimination in places where twenty years ago malaria was endemic.

The present study evaluates the potential impact of different IVM strategies in the future, aiming to achieve optimal results in malaria reduction. The analysis was conducted using a model inspired by the Malaria Management Model (MMM) developed by the Millennium Institute (Pedercini et al., 2011) [https](https://journals.plos.org/plosone/article?id=10.1371/journal.pone.0027771).

The scope of MMM is the whole sub-Saharan region, and it contains a representation of malaria transmission processes, vector control, and case management. The model also links dynamically these components to population development, health, education, and economic production.

Using the MMM as a point of departure, we have developed the Kenya Malaria Model (KMM), which is a national level model that can be used as a decision support tool to facilitate the design of effective malaria policies, as well as to capture the major dynamics of malaria and there impacts on other socio-economic sectors.

The resulting simulations from the model aim to evaluate different IVM interventions under different scenarios of development prospects, climate change, or effectiveness of anti-malaria methods. Such scenarios can help to identify adequate interventions to maximize the reduction in malaria transmission while observing their repercussions, for example, on population development, health, education, or economic production.

Finally, the model allows for estimating the possible cost of eliminating malaria in Kenya on a mid-term horizon under different scenarios and considering different combinations of IVM interventions.

## 3. Situation of Malaria in Kenya

### 3.1. Malaria Prevalence

The Kenya National Malaria Strategy (KNMS) has been developed in line with the Government’s first Medium-Term Plan of Kenya Vision 2030 and the Millennium Development Goals (MDGs), as well as Roll Back Malaria partnership goals and targets for malaria control. KNMS has been designed to cover the period 2009-2017, and the plan envisions achieving a malaria-free Kenya (Kenya Division of Malaria Control & Ministry of Public Health and Sanitation, 2009) [https](https://www.nationalplanningcycles.org/sites/default/files/country_docs/Kenya/kenya_national_malaria_strategy_2009-2017.pdf).

In the 1980s and 1990s, malaria prevalence in Kenya was on the rise, with epidemics reported in the western highlands and along the coast (Hay et al., 2002) [https](https://pmc.ncbi.nlm.nih.gov/articles/PMC2738480/). By the year 2000, the average malaria prevalence in Kenya was more than 20% (A. Noor et al., 2016) [https](https://inform.kemri-wellcome.org/wp-content/uploads/2021/06/Kenya-INFORM-Epi-Profile-2016.pdf), and, although we don’t have accurate data from the 1980s and 1990s, the absence of national malaria programs allows us to presume that the prevalence during those decades was similar to what it used to be in 2000, following the natural prevalence of the country.

Since the National Malaria Control Program started to be autonomous in 2000, malaria prevalence has been significantly reduced in most of the regions thanks to the increased financial support. According to national reports, malaria prevalence was 11% in 2010, and about 8% by 2015 (National Malaria Control Programme - NMCP/Kenya et al., 2016) [https](https://dhsprogram.com/pubs/pdf/MIS22/MIS22.pdf).

The situation is, however, still complicated: Kenya had an estimated 8 million malaria cases in 2017, and about 35 million out of the 50 million inhabitants still live in malaria risk areas. The western part of the country, for instance, is a very densely populated region where the malaria prevalence is higher than 30% (WHO, 2018a) [https](https://iris.who.int/handle/10665/275867).

To have a general vision of the malaria development over time, Figure 1 provides an overview of malaria prevalence throughout Kenya in 2000, and in 2015.

**Figure 1:**
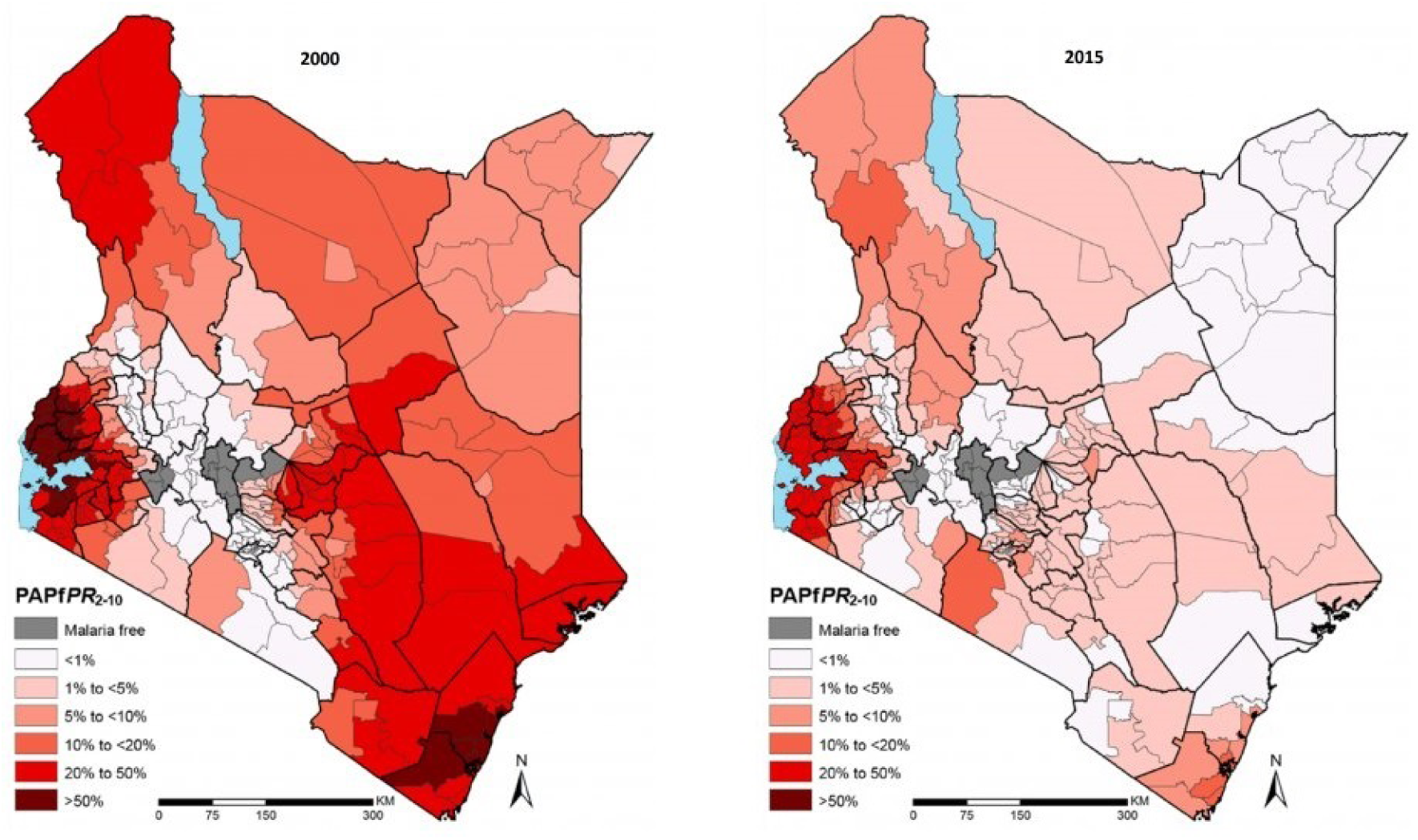
Map of malaria prevalence in Kenya. Years 2000 and 2015 (A. Noor et al., 2016)

**Figure 2:**
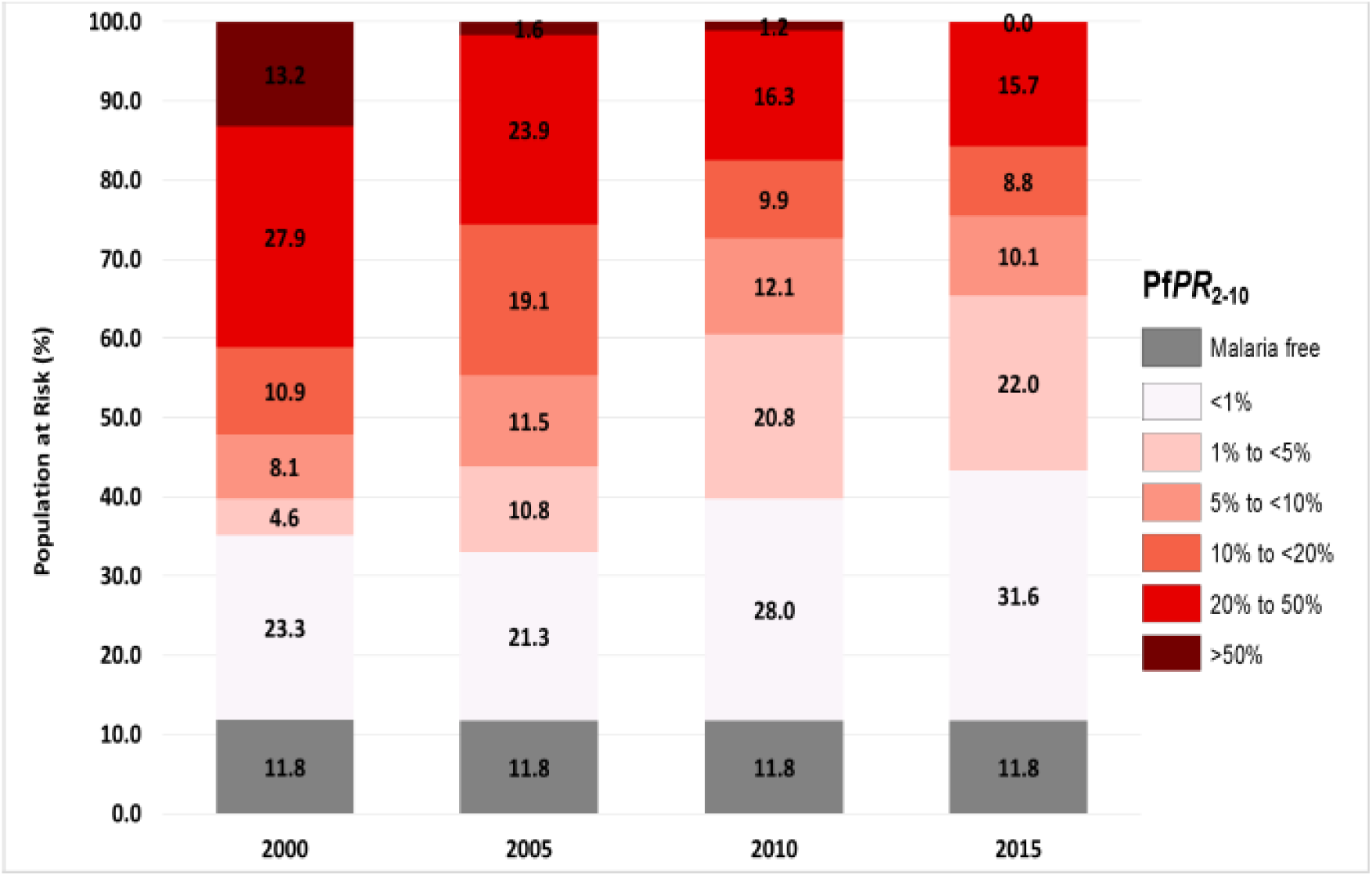
Stratification of malaria prevalence in Kenya by 2000, 2005, 2010 and 2015.

**Figure 3:**
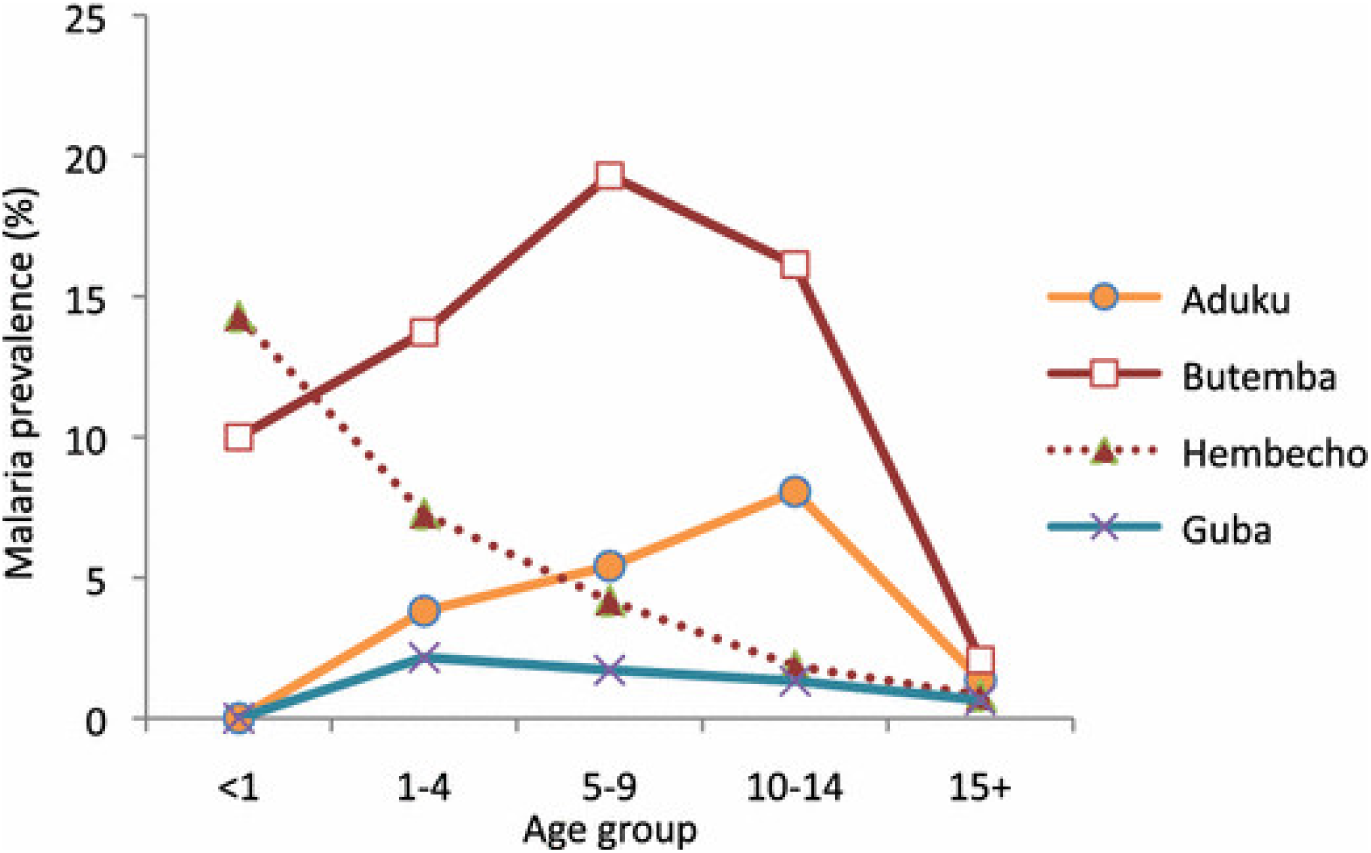
Malaria prevalence among different age groups in 4 sites (Abeku et al., 2015)

**Figure 4:**
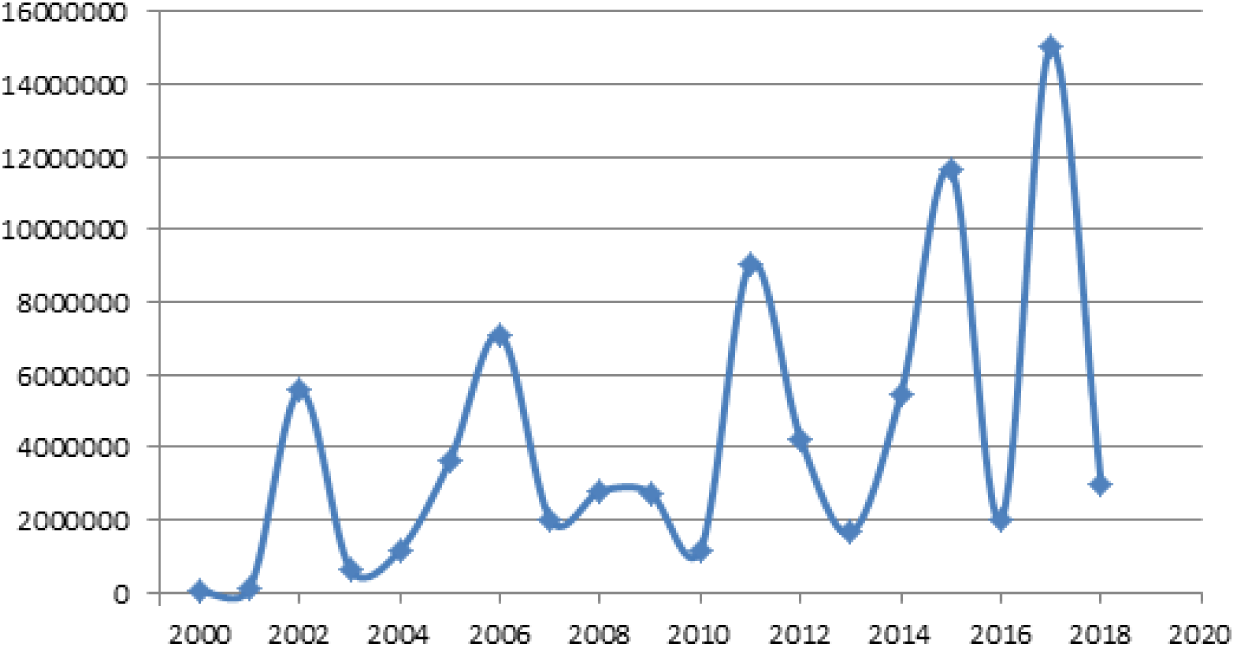
Nets distributed in Kenya between 2000 and 2018 (WHO)

As it appears from the map, Kenya has experienced a significant decrease in malaria prevalence, being lower than 5% nowadays in most of the country. In fact, it is estimated that 60-70% of the Kenyan land has a parasite prevalence of less than 5%, where 78% of the population lives (Kenya Division of Malaria Control & Ministry of Public Health and Sanitation, 2011) [https](https://dhsprogram.com/pubs/pdf/mis7/mis7.pdf). More concretely, the population in areas that are malaria free (or <1%), increased from 35.1% in 2000 to 53.6% in 2015. Malaria-free areas were defined on the basis of temperature limits and, therefore, the proportion of the population in this zone remained constant throughout (A. Noor et al., 2016). (Population grows, but the proportion of population in a given area remains constant over time). In this regard, and according to the WHO Malaria Reports (WMR), the proportion of people living in malaria risk areas has slightly changed over time: It was 76% by 2009 (WMR 2009), and 75% during the period 2010-2014. From WMR 2015 and on, this fraction has remained 70%.

According to the Kenya Malaria Indicator Survey 2010 (KMIS 2010) (Kenya Division of Malaria Control & Ministry of Public Health and Sanitation, 2011), malaria prevalence by 2010 was nearly three times as high in rural areas (12 per cent) as in urban areas (5 per cent). The lake endemic zone had the highest prevalence of malaria overall (38 per cent), while the prevalence in the other zones reached levels below 5 per cent.

The KMIS 2015 indicates that, the prevalence of malaria among children (by microscopy) was 8 percent, a decline from the existing 11 percent at the time of the KMIS 2010. Malaria prevalence was highest among children aged 10-14 years (11 percent), followed closely by children aged 5-9 years (10 percent). Among children aged 6-59 months (0-5 years), who are considered especially vulnerable, the malaria prevalence decreased from 8 percent in 2010 to 5 percent (National Malaria Control Programme - NMCP/Kenya et al., 2016) [https](https://dhsprogram.com/pubs/pdf/MIS22/MIS22.pdf).

In 2015, the prevalence was more than three times higher in rural areas than in urban areas (10 percent and 3 percent, respectively). A similar pattern that was observed in both, the KMIS 2007 and KMIS 2010, and that reflects the fact that urban areas tend to have lower malaria transmission than rural areas (National Malaria Control Programme - NMCP/Kenya et al., 2016).

The prevalence in 2015 continued to be much higher in the lake endemic zone than in other zones, but the rate among children aged 6 months to 14 years was markedly lower in 2015 (27 percent) than in 2010 (38 percent). On the other hand, in the coast endemic area, the malaria rate among children aged 6 months to 14 years increased from 4 percent in 2010 to 8 percent in 2015 (National Malaria Control Programme - NMCP/Kenya et al., 2016).

The next diagram, extracted from the Ministry of Health (2016) (A. Noor et al., 2016), indicates the proportions of the population under different levels of malaria prevalence in Kenya, during the years 2000, 2005, 2010 and 2015:

Based on this diagram, the national average prevalence in 2000 was estimated to be between 18.5% and 27.5%. (In 2005 (11.8% - 18.3%), in 2010 (8.5% - 13%), and in 2015 (7.4% - 11.4%)).

As mentioned before, KMIS 2010 and KMIS 2015 report that the malaria prevalence was 11% and 8%, respectively. These results are actually within the nationally estimated intervals.

It is important to keep in mind that the prevalence presented in the reports is measured among children (PfPR2-10 = Plasmodium falciparum Parasite Rate standardized to the age group 2-10).

In this sense, the prevalence of malaria is generally higher in children, especially under-5s, than in adults (Awosolu, Yahaya, & Farah Haziqah, 2021) [https](https://pmc.ncbi.nlm.nih.gov/articles/PMC8380643/). Young children are also the most susceptible to malaria due to a lack of acquired functional immunity, which older children and adults develop due to multiple exposures (Ranjha et al., 2023) [https](https://www.sciencedirect.com/science/article/pii/S2667009723000519?via%3Dihub). One study shows the malaria prevalence among different age groups in 4 sites. In all of them the malaria prevalence of people over 15 years old is among the lowest (Abeku et al., 2015) [https](https://malariajournal.biomedcentral.com/articles/10.1186/s12936-015-0852-7):

Therefore, despite the uncertainties, it is possible that the actual overall prevalence in Kenya may be slightly lower than the figures presented in the reports.

In summary, when compared with the situation in 2000, endemicity has been significantly reduced across the country. However, some regions with yet high endemicity are also very densely populated, implying many people living at risk of malaria transmission. It is important to note that malaria reduction is resulting from continuous years of malaria interventions. Any decrease in efforts would involve the return of endemic malaria to the regions where the pre-elimination state has been achieved.

### 3.2. Malaria Interventions

As mentioned above, malaria interventions include treatment and prevention. Successful treatments are based on prompt disease recognition and the use of adequate and high-quality therapies for eradication of the parasite causing malaria. Methods for prevention are very diverse (National Malaria Control Programme, 2019) https, and components within the IVM framework include:

- The use of personal protective measures: Bed nets, wearing of protective clothing or repellents which appear in various forms.
- Chemical control: Indoor Residual Spraying (IRS), outdoor spraying.
- Environmental Management (EM): Environmental control measures for vector control, or vector source reduction.
- Biological Control Measures: Larviciding, or larval source management (LSM).
- Sensitization and social mobilization: Advocacy for social behavioral change.

From all the preventive methods, the core vector control strategies implemented in Kenya have been the distribution of long-lasting insecticidal nets (LLINs) to population at risk, indoor residual spraying (IRS) in targeted areas, and larval source management (LSM) (National Malaria Control Programme - NMCP/Kenya et al., 2016).

The targeted population for bed net distribution consists mostly of those living in endemic regions: Before the 2000s, the bed net coverage in Kenya was residual (Kamau et al., 2017) [https](https://pmc.ncbi.nlm.nih.gov/articles/PMC5688631/). Even by 2003, data still shows that only 4 percent of pregnant women slept under an insecticide-treated mosquito net (Central Bureau of Statistics, Ministry of Health, & Macro, 2004) [https](https://www.knbs.or.ke/reports/kdhs-2003/). During 2011, however, more than 10 million bed nets were distributed, and approximately 60% of the population had a bed net. By 2017 estimations are that in accordance with the current programs, almost all the population at risk is already covered under the assumption of one LLIN per every second person, also defined as universal coverage (Kenya Division of Malaria Control & Ministry of Public Health and Sanitation, 2009), (Kibe, Kamau, Gachigi, Habluetzel, & Mbogo, 2019) [https](https://pmc.ncbi.nlm.nih.gov/articles/PMC6616035/).

As a summary of the net delivery in Kenya, apart from the routine distributions, mass distribution campaigns delivered 3.4 million nets in 2006, 10.6 million nets in 2011-2012, 13.1 million nets in 2014-2015 (National Malaria Control Program, 2017) [https](http://www.nmcp.or.ke/index.php/resource-centre/download-centre/category/5-surveillance-monitoring-and-evaluation), and 15 million nets in 2017 (WHO, 2018b). The graph below shows the annual nets distributed in Kenya between 2000 and 2018:

The bed nets distributed were treated with insecticide in order to repel mosquito (such nets are commonly called “Insecticide treated Nets” (ITNs)). Conventional ITNs need to be retreated every 6 months, but since 2007 the GoK is mostly delivering long lasting insecticide nets (LLIN), an improved version of ITN which only requires retreatment or substitution after about 3 years (A. M. Noor, Amin, Akhwale, & Snow, 2007) [https](https://journals.plos.org/plosmedicine/article?id=10.1371/journal.pmed.0040255).

The approximate costs of LLIN per person are 6 US$, where about 5 US$ corresponds to the cost of the net itself, and the rest is transportation/distribution cost (Ntuku et al., 2017) [https](https://malariajournal.biomedcentral.com/articles/10.1186/s12936-016-1671-1).

When referring to insecticide spraying, indoor residual spraying (IRS) has been the most popular type of intervention. IRS interventions are implemented in targeted areas based on two fundamental criteria (Kenya Division of Malaria Control & Ministry of Public Health and Sanitation, 2011):

- Burden reduction
- Response or prevention of epidemics (it’s known when an epidemic could occur)

IRS interventions are made once per year, and the most common compounds used in Kenya are pyrethroids. Also organophosphate is used, but this product is more expensive since it needs to be sprayed twice per year. The approximate cost of IRS interventions per person is 6 US$, where the chemical is 2 US$, and the rest is around 4 US$ for transportation, spraying operations, local labor, etc.

Both LLITNs distribution and IRS need sensitization and social mobilization in order to achieve the desired efficacy (Kenya Division of Malaria Control & Ministry of Public Health and Sanitation, 2011). Similarly, the level of education among the Kenyan population is also a key indicator affecting interventions’ efficacy. Reports indicate that general knowledge in Kenya about malaria transmission is currently at 95%. Nevertheless, still many people do not use or they misuse the nets that have been given. It is estimated that the cost of sensitization for LLITNs interventions is about 1 US$ per person (Temperley et al., 2008) https, which adds to the cost of the net and transport/distribution.

Apart from IVM interventions, malaria control in Kenya also includes case management and measures that incur other expenses. The diagram below displays the proportions of the resources to cover the different objectives of the Kenya malaria strategy (National Malaria Control Programme, 2019).

The Figure 5 shows that malaria prevention accounts for the highest share of the total resource need (67 percent), followed by diagnostics and treatment (14 percent).

**Figure 5:**
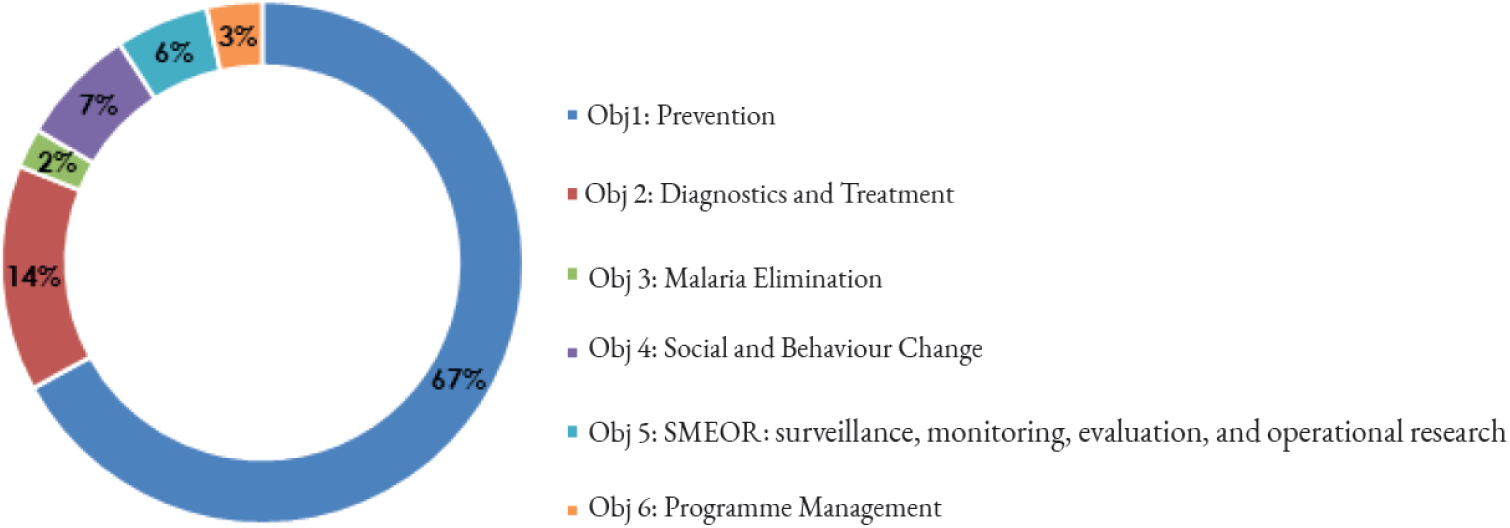
Proportion of resources per KMS objective area (National Malaria Control Programme, 2019)

**Figure 6:**
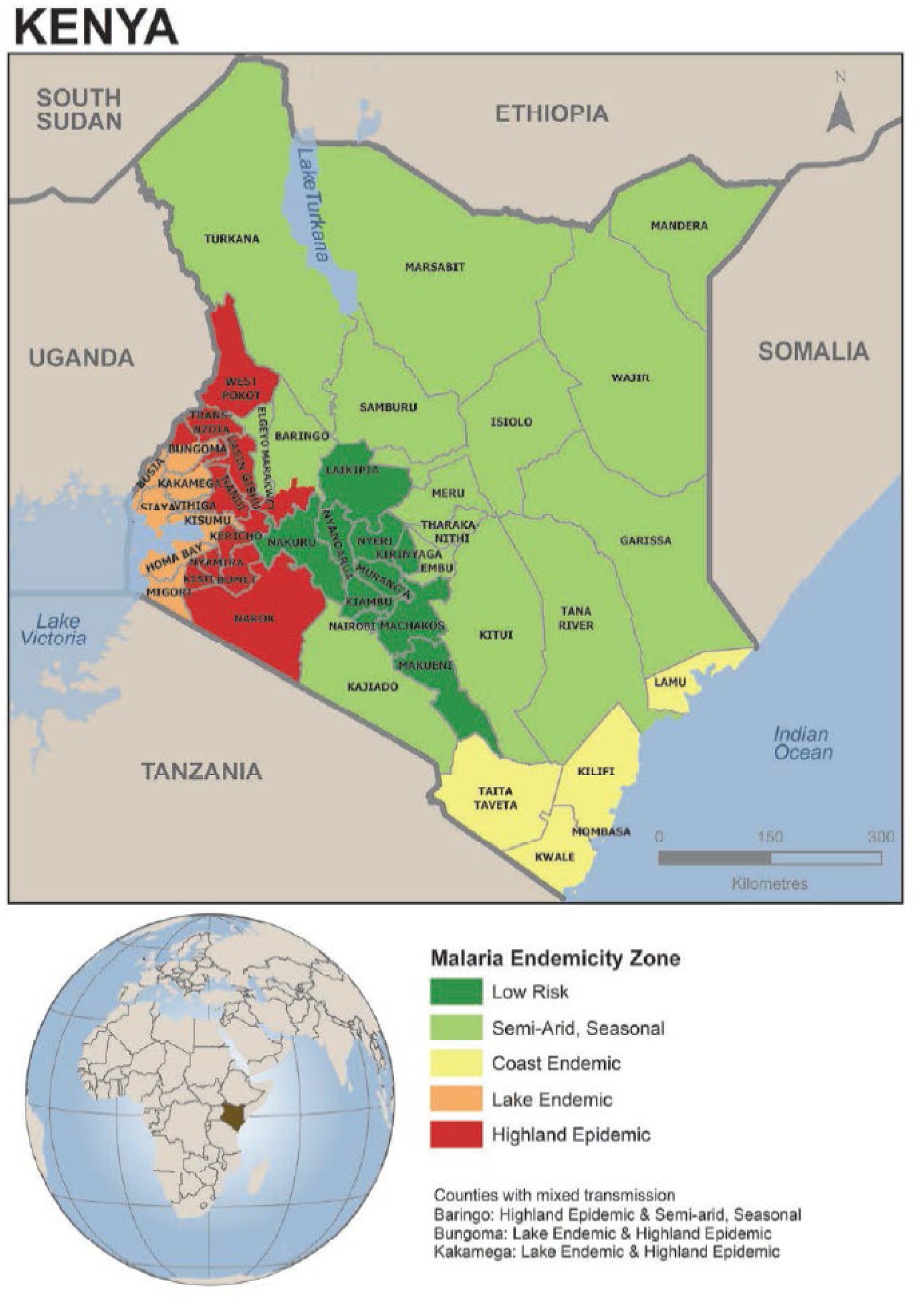
Map of Kenya with the malaria endemic zones (National Malaria Control Programme - NMCP/Kenya et al., 2016)

**Figure 7:**
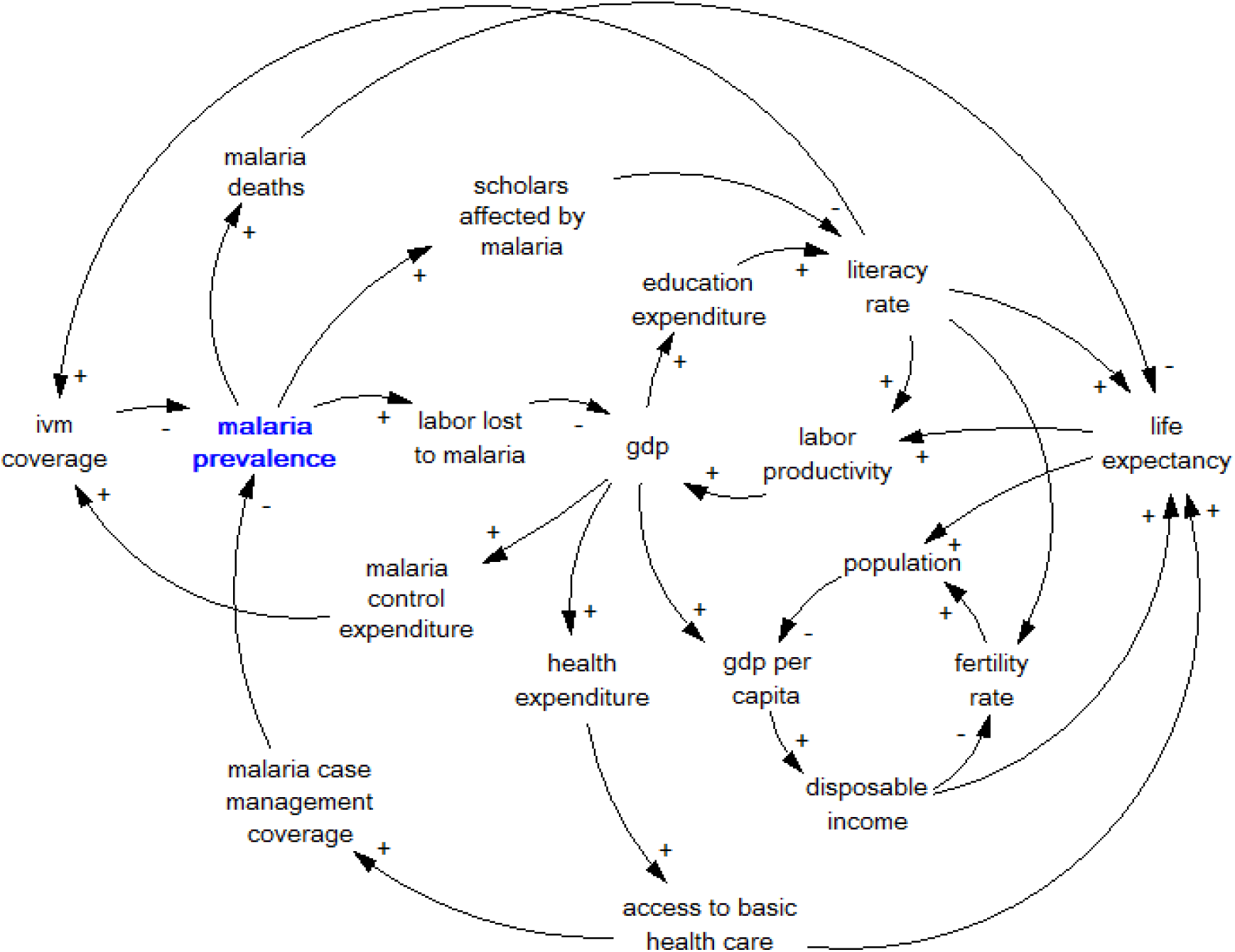
Summarized causal loop diagram of the simulation model.

**Figure 8:**
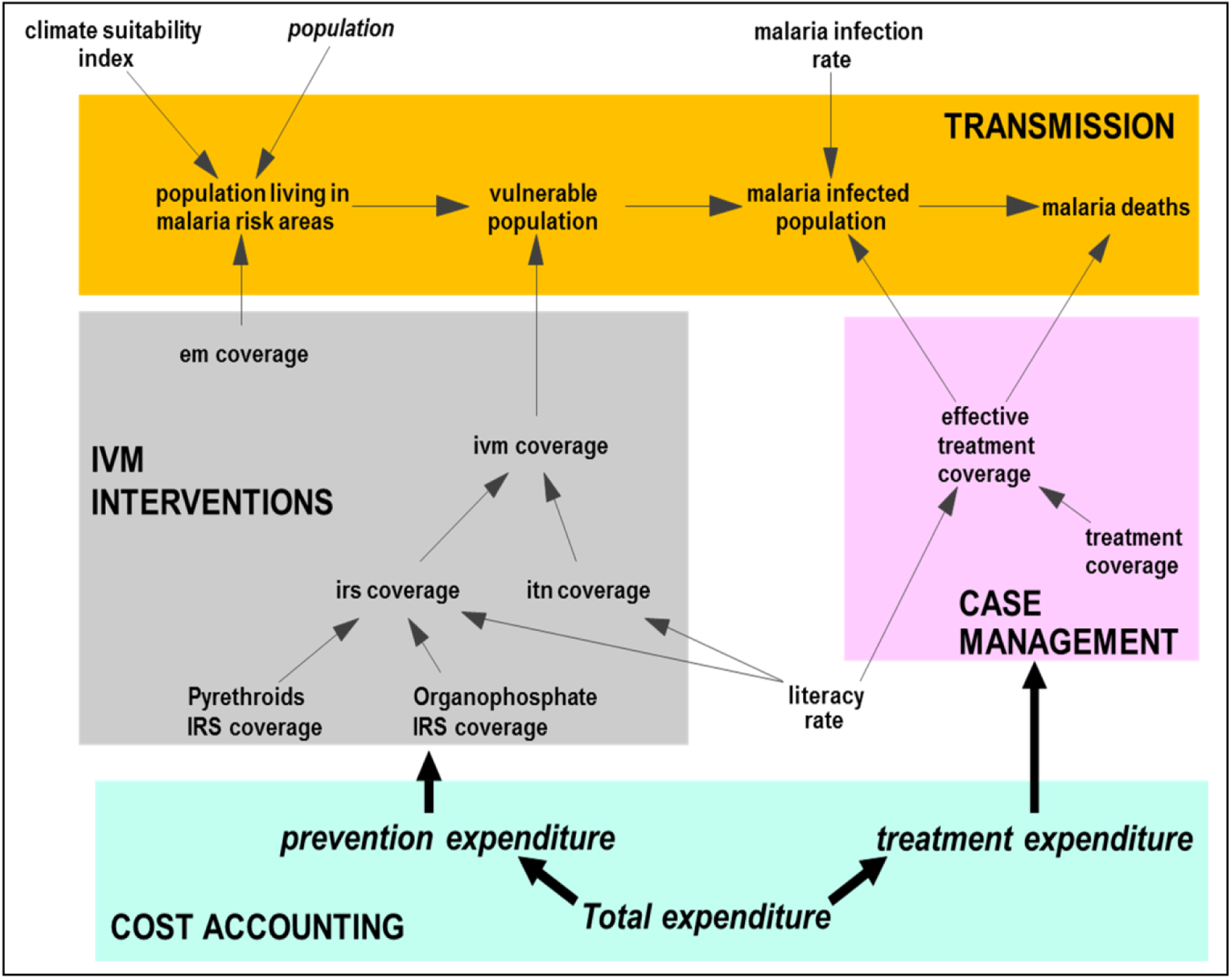
Malaria subsectors in the model.

### 3.3. Geographical Distribution of Interventions

During the implementation of IVM strategies, the decisions are basically made prevalence-based. For instance, in Kenya IRS is not implemented in regions with low prevalence. IRS starts being considered worthwhile when the malaria prevalence is 10% or higher. As for the bed nets, in the provinces of Western, Rift valley, Coast and Nyanza LLINs are delivered to the entire population. In the rest of the country LLINs are only delivered to people in the highest risk categories; pregnant women and children of less than one year old. The delivery policy is such that in regions of malaria prevalence the net distribution aims at placing one LLIN per two persons.

In order to have a more comprehensive view of the interventions carried out in the different regions, Kenya can be divided into four malaria epidemiological zones (Kenya Division of Malaria Control & Ministry of Public Health and Sanitation, 2010) [http](https://www.c-hubonline.org/baad0977-6068-4330-bd3d-39e9fd3b20b3/eyJhbGciOiJBMTI4S1ciLCJlbmMiOiJBMT):

#### -A: Endemic areas (lake and coast)

These are areas of stable malaria transmission (with altitudes ranging from 0 to 1,300 m) around Lake Victoria in western Kenya and in the coastal regions. Rainfall, temperature and humidity are the determinants of the perennial transmission of malaria. The vector life cycle is usually short with high survival rate due to the suitable climatic conditions. Transmission is intense throughout the year, with annual Entomological Inoculation Rates (EIR)^2^ between 30 and 100.

#### -B: Malaria epidemic prone areas of western highlands of Kenya

Malaria transmission in the western highlands of Kenya is seasonal, with considerable year-to-year variation. The epidemic phenomenon is experienced when climatic conditions favor sustainability of minimum temperatures around 18°C. This increase in minimum temperatures during the long rain period favors and sustains vector breeding resulting in increased intensity of malaria transmission. The whole population is vulnerable and case fatality rates during an epidemic can be up to ten times greater than what is experienced in regions where malaria occurs regularly.

#### -C: Semi-arid, seasonal malaria transmission

This epidemiological zone in arid and semi-arid areas of northern and south-eastern parts of the country experiences short periods of intense malaria transmission during the rainfall seasons. Temperatures are usually high and water pools created during the rainy season provide the malaria vectors breeding sites. Extreme climatic conditions like El Niño Southern Oscillation lead to flooding in these areas, causing epidemic outbreaks with high morbidity rates due to the low immune status of the population.

#### -D: Low risk malaria areas

This zone covers the central highlands of Kenya including Nairobi. The temperatures are usually too low to allow completion of the sporogonic cycle (production of sporozoites) of the malaria parasite in the vector. However, increasing temperatures and changes in the hydrological cycle associated with climate change are likely to increase the areas suitable for malaria vector breeding with the introduction of malaria transmission in areas where it previously never existed.

The following map includes the four malaria epidemiological zones in Kenya. Although the endemic zones do not represent a large area in the malaria endemicity map, these regions contain almost one third of the total population in Kenya.

In the diagram below, we can observe the population distribution for the four malaria epidemiological zones. Population numbers are from 2009, but the proportions of the population have been kept constant in the given zones:

Table 2^3^ represents the policies for malaria interventions depending on the malaria epidemiological zone:

**Table 1:** Population distribution by malaria epidemiology (Kenya Division of Malaria Control & Ministry of Public Health and Sanitation, 2010)

| Epidemiologic strata | Total projected population 2009 | % of total projected population | Cumulative % of total projected population |
| --- | --- | --- | --- |
| Lake stable endemic & Coast seasonal stable Endemic | 11,452,028 | 29.0% | 29.0% |
| Highland epidemic Prone | 8,007,718 | 20.3% | 49.3% |
| Seasonal low transmission including arid and Semi arid | 8,029,683 | 20.4% | 69.7% |
| Low Risk | 11,933,834 | 30.3% | 100.0% |
| **Total** | **39,423,263** | **100%** |  |

**Table 2:** Stratification of districts by malaria risk and appropriate intervention in 2010 (Kenya Division of Malaria Control & Ministry of Public Health and Sanitation, 2010)

| Epidemiology | Case Management | LLIN | IRS | Health Education/BCC | IPTp | EPR | Surveillance |
| --- | --- | --- | --- | --- | --- | --- | --- |
| Lake stable endemic & Coast seasonal stable Endemic | X | X | X | X | X |  | X |
| Highland epidemic prone | X | X | X | X |  | X | X |
| Seasonal low transmission including arid and Semi arid | X |  |  | X |  | X | X |
| Low risk | X |  |  | X |  |  | X |

From table 2, we observe that in 2010 LLINs were aimed to be delivered in the two zones with higher risk. These two areas comprise roughly half of the population in Kenya, and 70% (0.5/0.7) of all the population at risk. Therefore, when everybody in these areas has access to LLINs, Kenya would attain 70% universal coverage in the country, which is defined as one LLIN for every two persons at risk of malaria. But, as mentioned before, 2017 estimations indicate that the universal coverage has already been reached for everybody living in malaria risk areas.

IRS has only been implemented in endemic areas for disease burden reduction, and also IRS campaigns for malaria prevention where epidemics can occur.

Case management and surveillance are implemented in all the zones: Case Management is the combination of diagnosis and treatment, and surveillance basically consists of the monitoring of malaria incidence: “Surveillance is the continual and systematic collection, analysis and interpretation of malaria data essential to the planning, implementation and evaluation of interventions. It is also a tool for measuring the health status of a population” (World Health Organization. Malaria, 2005).

Health education includes sensitization activities that promote public awareness of malaria transmission. Communication programs embrace basic strategies to increase demand for and acceptance of malaria interventions and services, including information, education and communication (IEC) and behavior change communication (BCC) methodologies (Roll Back Malaria Partnership, 2008).

Beyond LLINs and IRS interventions, little EM and biological control has been implemented in Kenya. However, some environmental modifications introduced through infrastructures or agricultural projects provided EM indirectly. In this regard, the coastal region is an unique area in Kenya; IRS has rarely been used, but thanks to the implementation of LLINs, and EM with larviciding, the region has experienced prevalence reductions from 60% in the late nineties (Mbogo et al., 2003) to 4% by 2010 (Kenya Division of Malaria Control & Ministry of Public Health and Sanitation, 2011). However, it seems that the prevalence in coastal Kenya is on the rise again, as in 2015 it was reported 8% prevalence (National Malaria Control Programme - NMCP/Kenya et al., 2016) (Snow et al., 2015).

## 4. Materials and Methods

This section introduces the reader to the simulation model that has been used as the cornerstone for the study. The model was created using the System Dynamics methodology (Sterman, 2000), and at its core, it consists of a structure that reproduces over time the historical behavior of Kenya’s most relevant socio-economic development indicators since 1980.

Attached to this structure, the model includes the malaria sectors, which are dynamically linked two-way with the rest of the model: In one direction, the malaria sectors use various inputs generated from the rest of the model, such as population, literacy rate or access to basic healthcare. In the other direction, malaria prevalence and deaths are used to assess their impact on education, productivity, or life expectancy, among others, generating the feedback loops that drive the malaria dynamics in Kenya and their impact on society and economy over time.

As mentioned before, the malaria sector builds on the Malaria Management Model (MMM) (Pedercini et al., 2011), although with some important structural differences. The causal loop diagram (CLD) below illustrates how some of the most important variables in the system influence each other through cause-and-effect relationships. The CLD also maps out feedback loops, which are essential to our understanding how the system behaves over time:

As observed from the diagram, malaria prevalence is interconnected with the rest of the model in areas of health, education, and GDP production. Later, in the section on results, we will elaborate on some of the loops that can be extracted from this CLD, which are crucial to understand the dynamics of malaria and its impact on the socio-economic development in Kenya.

To provide a better understanding of the key factors and mechanisms that drive malaria in the model, the next paragraphs will provide a short description of the specific malaria subsectors.

Just as the rest of the model, the malaria sector is developed by means of the System Dynamics methodology. The malaria sector can be divided into four subsectors:

1. *IVM Interventions*
2. *Case Management*
3. *Malaria Transmission*
4. *Malaria Costs Accounting*

All the sectors indicated above interact dynamically, generating endogenously the major trends of development for malaria and highlighting the impact of malaria interventions. The diagram below provides a simplified representation of the four malaria subsectors with the main indicators and some of the links connecting each other:

Further details on the structure of the malaria sub-sectors and their function are provided below.

### 4.1. IVM Interventions

This sector represents the implementation mechanisms and related costs of selected representative IVM interventions. These interventions are basically the combination of the most relevant protective measures: Bed-net or LLIN distribution, spraying operations (IRS), and environmental management (EM). Possible future achievements regarding vaccination are not considered, as the time required for developing such vaccination, its potential effectiveness, and the resources involved remain highly uncertain.

### 4.2. Case Management

This sector keeps track of the diagnosis and treatment coverage, and expenditures for the malaria infected population. Access to basic healthcare is used, together with the contribution of malaria treatment expenditure, to determine the theoretical treatment coverage.

The actual coverage is based on the theoretical coverage, but also the percentage of people who attend formal health services, and the average efficacy of the malaria treatments in terms of drug resistance of the parasite that causes malaria. Such treatment coverage strongly affects malaria mortality, malaria case fatality rate, and the duration of the infectiveness.

### 4.3. Malaria Transmission

This sector provides a representation of the mechanisms underlying long-term dynamics of malaria infections and deaths.

Climatic conditions play an important role in malaria transmission: the density and level of activity of the vector, depend on climatic conditions such as temperature, rainfalls, and humidity. Consequently, more suitable climate conditions may turn low risk areas into high risk areas. Vulnerable population is determined based on the estimated proportion of population living in risk areas, and on the actual coverage of the malaria preventive methods (IVM). Actual coverage of IVM depends on the intensity of IVM interventions, as well as on the education level and per capita income of local population (disposable income also affects literacy rate).

Malaria infections are determined based on the interaction between vulnerable population and malaria infectious population through the mosquito population. Considering the short life-span of a mosquito (two or three weeks) when compared to the long-term time horizon of the model, the mosquito life-cycle is not explicitly represented in this model. On the contrary, the ability of the mosquito population to function as vector for the malaria parasite is implicitly represented through the malaria infectivity, which is the probability of infection after being bitten by an infectious mosquito, and the transmission rate of the malaria parasite, which is a proxy of the natural Entomological Inoculation Rate (natural EIR).

As mentioned before, the EIR is a measure of malaria transmission intensity, and, in this case, the natural EIR represents the number of infective mosquito bites a person receives per unit of time, typically per year, in the absence of any malaria intervention.

The deaths caused by malaria are calculated based on the malaria infected population and the malaria case fatality rate, which depends on the efficacy and coverage of malaria treatments, as described above.

### 4.4. Malaria Cost Accounting

This sector summarizes all economic costs of the epidemic and of implemented interventions. From this sector, we derive to other sectors the long-term impacts of malaria programs on human life expectancy, literacy rate, and productivity, among others.

The indicators calculated in this sector will allow for the extraction of a broad assessment of the desirability of alternative strategies to fight malaria.

## 5. Base Run Results from Key Indicators

As a mean of model validation, the model was subjected to a number of validity tests (Barlas, 1996), including both structural and behavioral tests. Regarding the former type of tests, the model underwent reiterative cycles of revision by local malaria experts, who reviewed the model structure and parameters (IVM workshops at ICIPE, Nairobi 2011, 2012 and 2015).

Regarding behavioral validation, a base run simulation was generated for the period 1980-2017, with the aim of assessing the model’s ability to replicate the historical trends for key malaria-related indicators. Due to the incompleteness and, at times, inconsistency of the available data, such a validation has been especially challenging with regard to some indicators.

More precisely, we observed an important disparity between simulated malaria cases and malaria cases from data collections before 2010. According to WHO Malaria Reports (WMR) (WHO, 2018a), and national reports such as the Kenya Malaria Indicator Surveys (KMIS) (Kenya Division of Malaria Control, Ministry of Public Health and Sanitation, Kenya National Bureau of Statistics, & National Coordinating Agency for Population and Development, 2009) (Kenya Division of Malaria Control & Ministry of Public Health and Sanitation, 2011) (National Malaria Control Programme - NMCP/Kenya et al., 2016), the number of malaria cases has been increasing during the period 2000-2010. Data collection from different sources showed values of around 3 million cases in 2002, and an almost uninterrupted growth over time until reaching approximately 11 million cases in 2011. Figure 9 displays the trend in the data collected from WMR.

**Figure 9:**
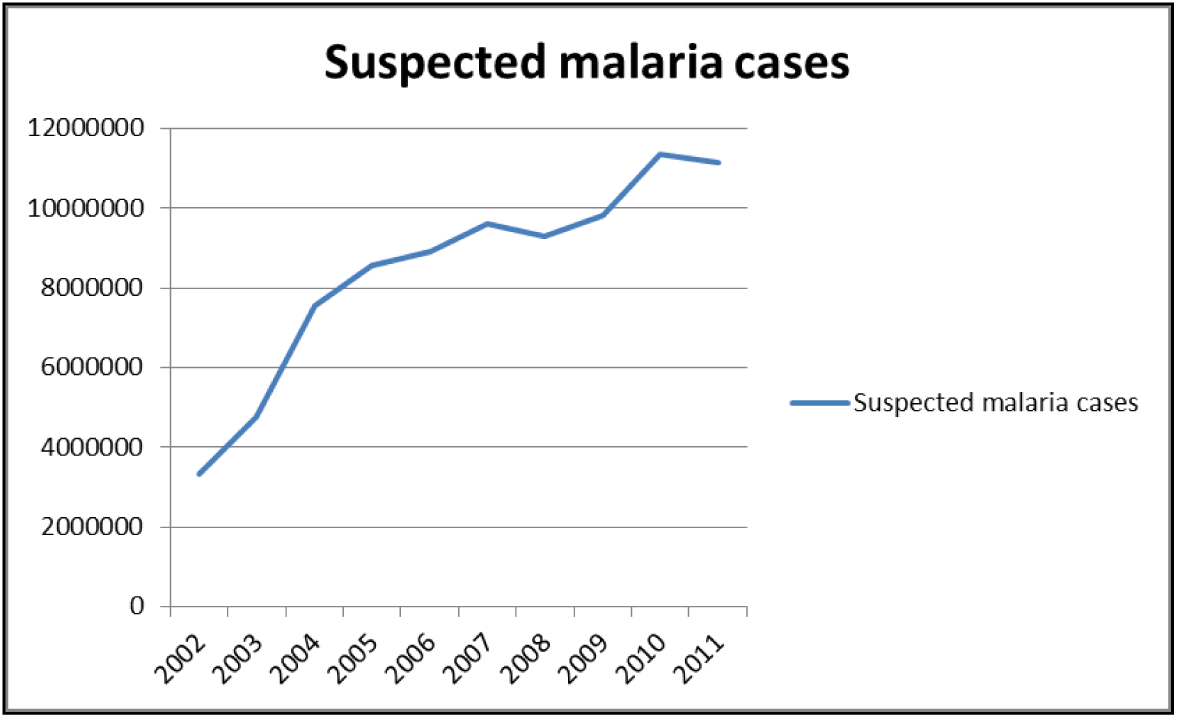
Suspected malaria cases 2002-2011 (source: WMR)

This growth indicates that the number of cases in 2011 was almost three times as high as in 2002. Even considering the significant population growth rate of Kenya, the suspected cases in 2011 per thousand people is double compared to 2002.

In addition, the KMIS 2010 indicates that the prevalence in children below five years increased from 4 per cent in 2007 to 8 per cent in 2010 (Kenya Division of Malaria Control & Ministry of Public Health and Sanitation, 2011) (The prevalence from 2007 is based on the KMIS 2007 (Kenya Division of Malaria Control et al., 2009)), suggesting that the endemicity increased during the period 2007 to 2010

This increase in malaria incidence that seem to emerge from collected data is not consistent with the growing intensity of anti-malarial interventions in the country. In fact, the efforts made in Kenya over the period 2000 to 2010 led to a situation where the population had more access to antimalarial resources than ever before.

Figure 10 (WMR) illustrates the evolution of coverage of ITN and IRS from 2000 to 2010: The increase in the percentage of households with access to ITNs during those years was substantial.

**Figure 10:**
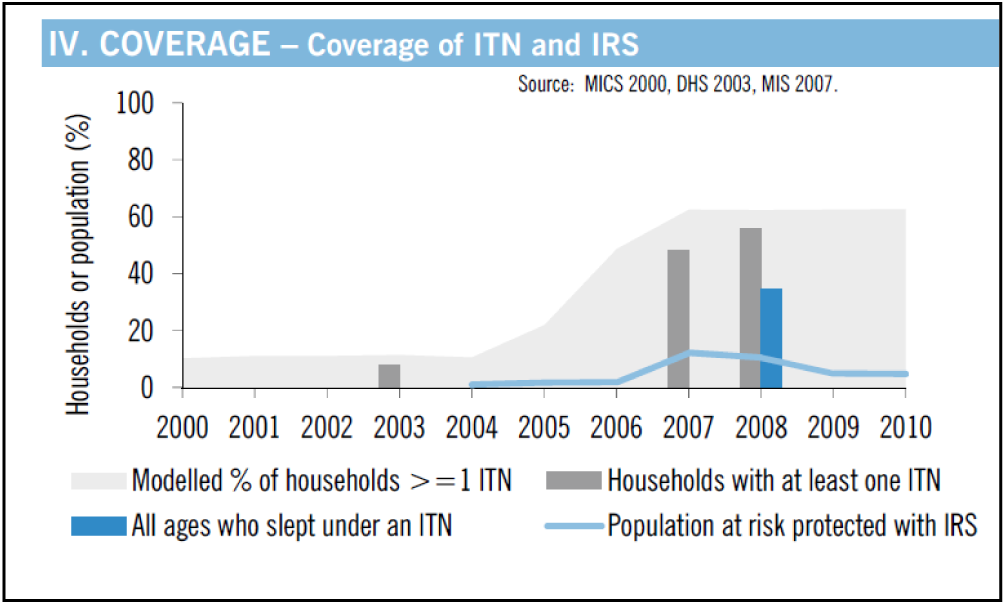
Coverage of ITNs and IRS (source: WMR)

Such increase in coverage of preventive interventions lead us to consider the possibility that the apparent increase in malaria cases was the result of a systemic underreporting of malaria incidence, possibly due to some problem at the survey level.

In this case, it was essential to interpret the data correctly in order to understand the system; otherwise we would conclude that the increase of the anti-malaria coverage over the period 2000-2010 only led to an increasing number of malaria cases over time.

Some other factors were analyzed with the purpose of finding what could be the reason to this apparent increase in the number of cases:

Since malaria is strongly related to temperature, rainfall and humidity (Small, Goetz, & Hay, 2003) https, one of the factors considered was that the increase in incidence could have been caused by a climatic change. But that does not seem to be the case based on the analysis of key main climatic indicators: Actually the average temperature in Kenya has been relatively constant during that period, and, if we take into consideration the rainfall, its behavior does not provide any pattern that could explain the increase of malaria cases. The Figure 11 shows trends for historical temperature and rainfall.

**Figure 11:**
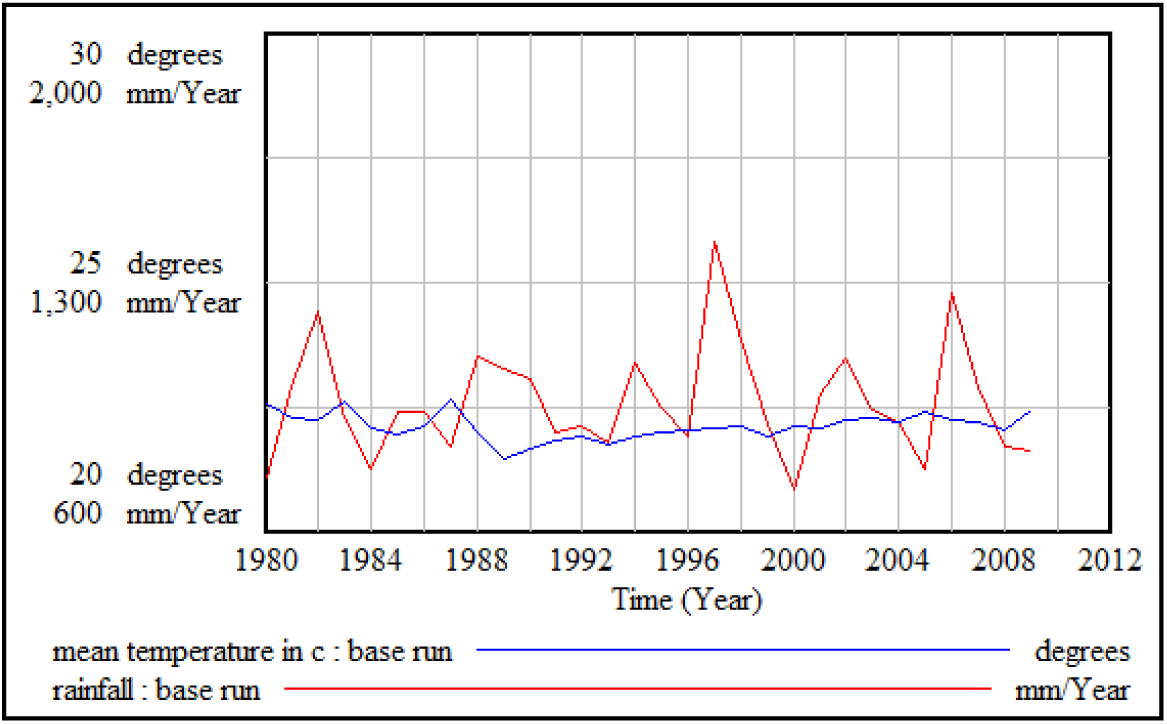
Average temperature (°C) and annual rainfall (mm) in Kenya (1980-2009)

Other socio-economic factors, such as literacy rate, per capita income, or malnutrition were also considered, but without finding any possible such driver for the given increase in the number of malaria cases. Rather, the increase in the number of cases might have been caused by an improvement of the malaria reporting system, which has expanded access to health services and to the population, increasing also the survey attendance. Also, the collection of data for malaria cases has certainly improved, achieving better monitoring and diagnosis of malaria cases, and increasing the quality of the data collections.

Therefore, we presume that only from 2010 the surveillance system has been able to provide numbers of reported cases which are reasonably close to the reality. Based on these premises, the development of the reporting system is assumed to have caused an artificial increase in the number of cases over time, while as the real number of malaria cases has in fact decreased.

The highlights of the Kenya profile in WMR 2008 and WMR 2009 seem to contradict each other with regard to the number of estimated malaria cases^4^. But at least both numbers are more in line with our simulation results. These reports, by the way, also question the validity of an increasing number in suspected cases over the years 2001 to 2007:

Figure 12 shows a comparison between the model results from the base run simulation (blue line) and the suspected malaria cases from the data collection (red line). Particularly for 2006, our model estimates 13.5 million cases, a figure that is between the estimation presented in WMR 2008 and in WMR 2009.

**Figure 12:**
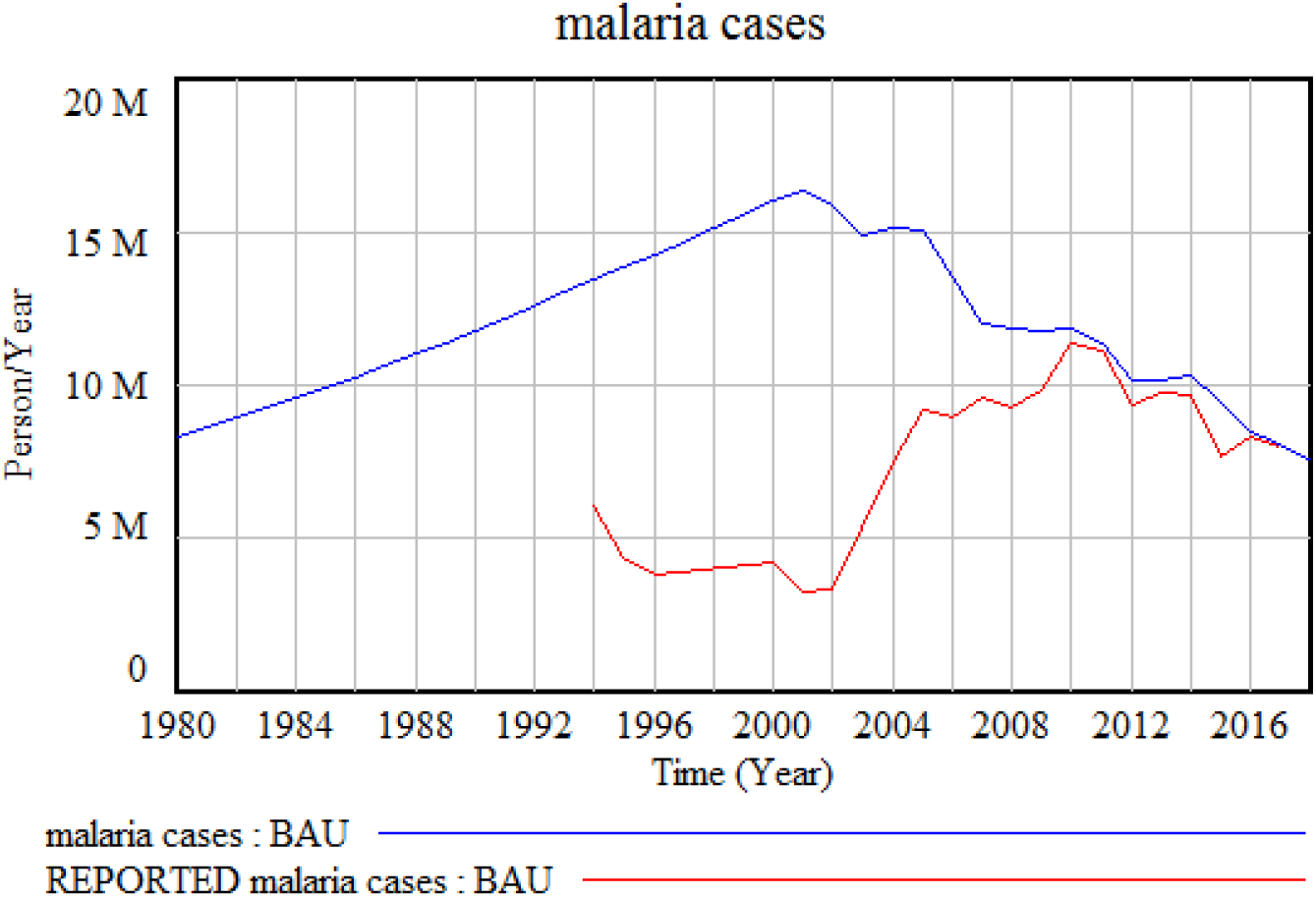
Model simulation vs. historical data for suspected malaria cases.

As observed in the graph, the simulation (blue line) displays a very different behavior of the total number of malaria cases compared to the data collected on suspected malaria cases (red line) for most of the analysis period. Only since 2010, when the surveillance and reporting system presumably became reliable enough to provide a realistic picture of the malaria situation in the country, the number of cases simulated by the model aligned with the suspected malaria cases from the reports.

The model simulation thus provides a more rational picture of the actual development of malaria cases over the 30 years before 2010. In fact, given the increasing implementation of IVM in Kenya since 2001, the expected trend in malaria cases over time is a decreasing curve. Prior to that, the number of cases was approximately coupled with the population growth in the country; without widespread malaria control programs in the 80s and 90s, we believe that Kenya kept a natural malaria prevalence, which, as mentioned before, it is estimated to be more than 20% at national level .

Therefore, the simulation is based on two premises: First, that the proportion of population living in malaria risk areas (or vulnerable population) has been relatively constant and around 70%-75% of the total population (WHO, 2011), and second, the number of cases is affected by the IVM coverage and the access to treatments. As a final note, the results of the model simulation are intended to be inclusive also for the cases that were not reported, including asymptomatic cases and those treated at home.

After showing simulated malaria cases, the next indicator is the malaria prevalence. Figure 13 shows the malaria prevalence in Kenya, i.e. the population fraction having malaria at any given point in time. The blue line represents the simulation, and the red line is the estimated average of the data on malaria prevalence described before:

**Figure 13:**
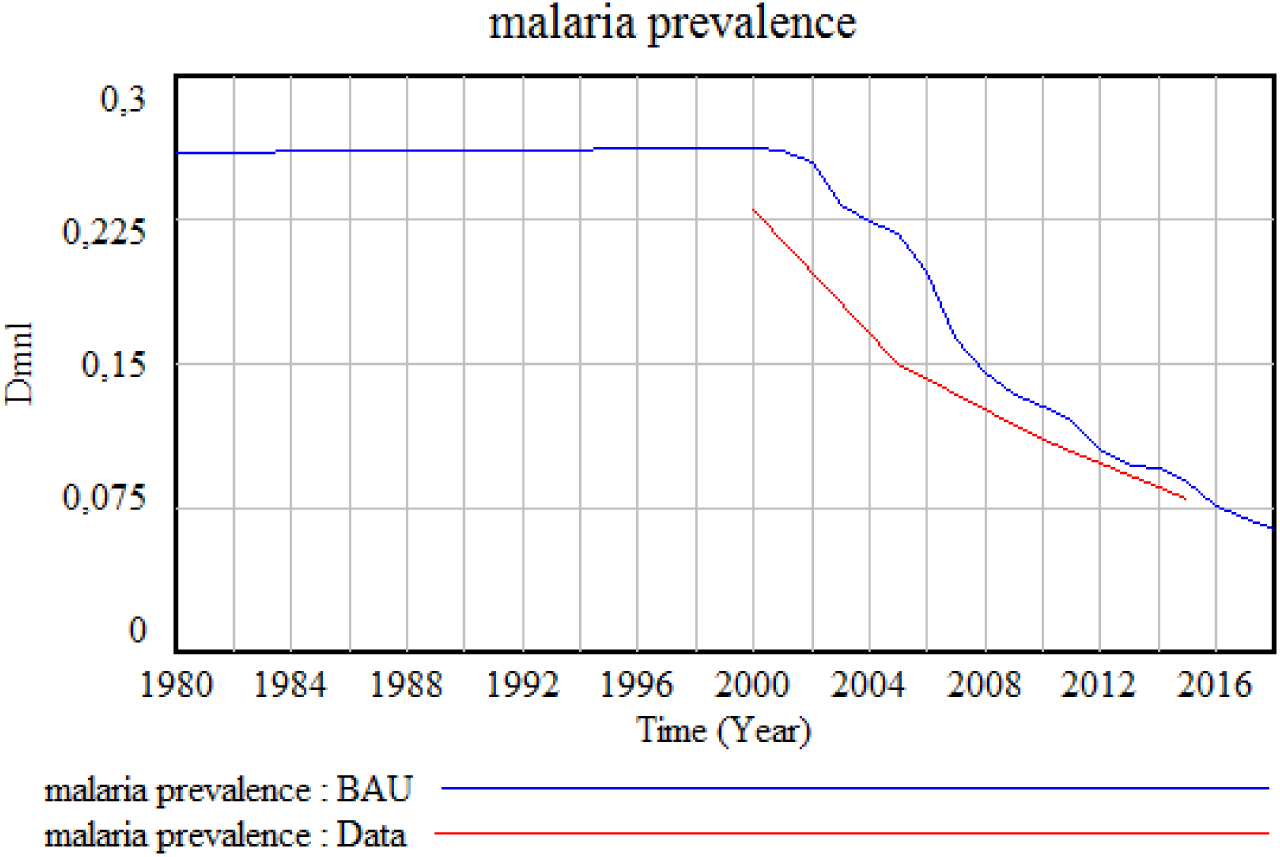
Model simulation vs. historical data for malaria prevalence in Kenya.

As mentioned before, we have assumed a relatively constant prevalence during the 80s and 90s, with a decrease from 2001 as a result of the malaria control programs. But as with the malaria cases, there is a significant level of uncertainty in the data, especially for the years before 2010, which is when we believe that malaria was underreported.

Something similar happens in the case of the number of malaria deaths; the historical data differs significantly from the simulation before 2007, and the most plausible explanation follows the same arguments as those discussed above for malaria cases. Figure 14 shows the simulated number of deaths versus the reported malaria deaths from the World Malaria Reports (2000-2006) and the KNBS Economic Surveys (2010-2017) (Kenya National Bureau of Statistics, 2018) [https](https://www.knbs.or.ke/?s=Economic+Survey&post_type=us_portfolio).

**Figure 14:**
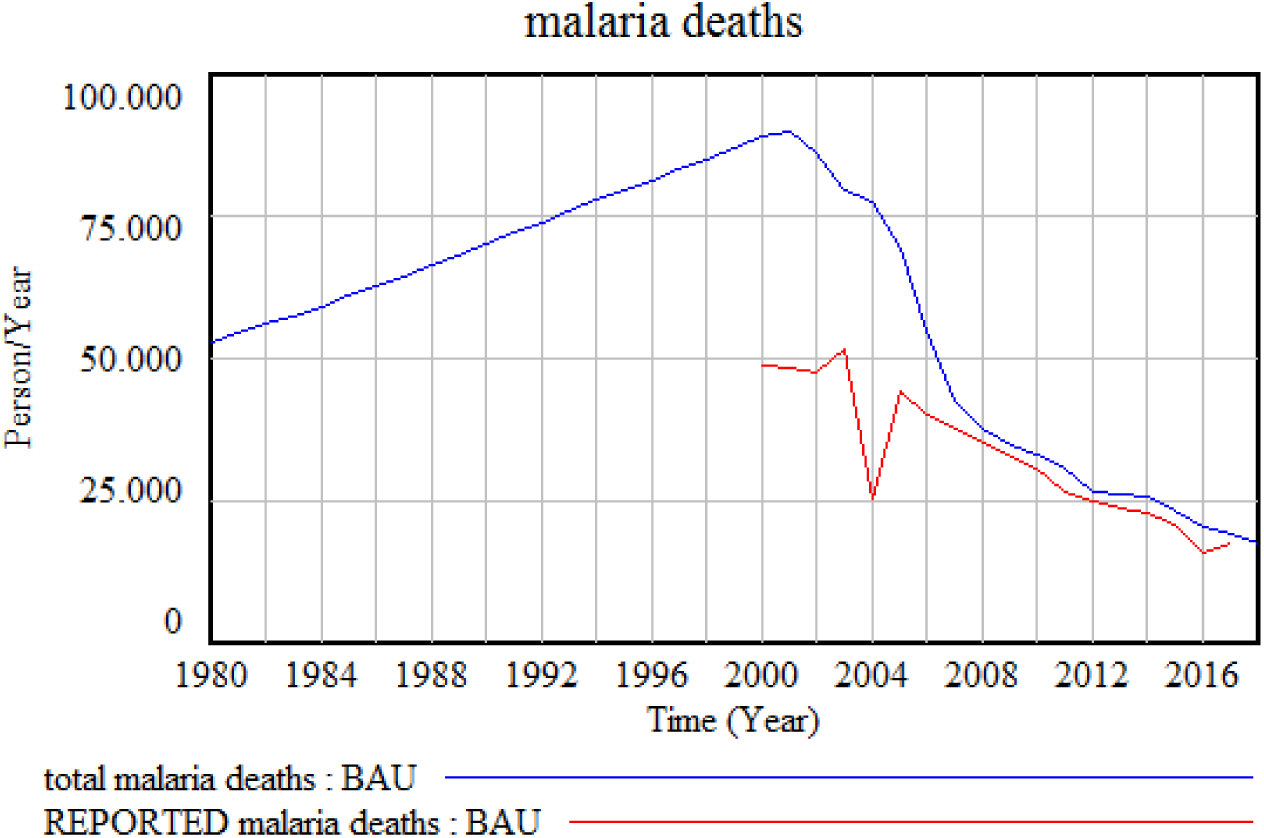
Model simulation vs. historical data for malaria deaths.

Although the number of deaths is generally better reported, there is still a small gap between our simulation and the data collection. We believe that this discrepancy originates from the proportion of the deaths of unknown cases that are attributable to malaria. In fact, without a proper diagnosis it is difficult to determine whether malaria is the real cause of the death or not. For example, according to the KNBS Economic Survey, in 2011 malaria accounted for 21.0 per cent on the average, for all causes of deaths registered (Kenya National Bureau of Statistics, 2011) [https](https://www.knbs.or.ke/reports/2011-economic-survey/). But if we consider 21% of all the estimated deaths in Kenya during the same year, the number of malaria deaths would be around three times higher than what has been reported.

There is also a large discrepancy in the numbers reported depending on the source: In the WMRs, the number of reported deaths from 2011 are around 12000 or less, while the KNBS Economic Surveys provide a more detailed breakdown of deaths by county, and the number of deaths seem to follow a more logical development according to the malaria prevalence in the country. The KMIS reports do not indicate information about deaths by malaria.

A final challenge related to the historical data has been to match the funding for malaria interventions with the cost of the interventions implemented:

The cost and implementation of antimalarial campaigns has increased significantly over the last decade. Since 2001, the GoK in collaboration with partners has deployed large amounts of LLINs and IRS interventions, together with an increased access to treatments, especially Artemisinin-based Combination Therapies (ACT). There is, however, also certain disparity between the calculated budgets to deploy such interventions, and the reported funding.

Figure 15 displays the reported funding in Kenya (red line) and the budget necessary to cover the reported interventions (blue line), which in this case, is the sum of preventive measures (ITNs, IRS, EM and Sensitization), and case management.

**Figure 15:**
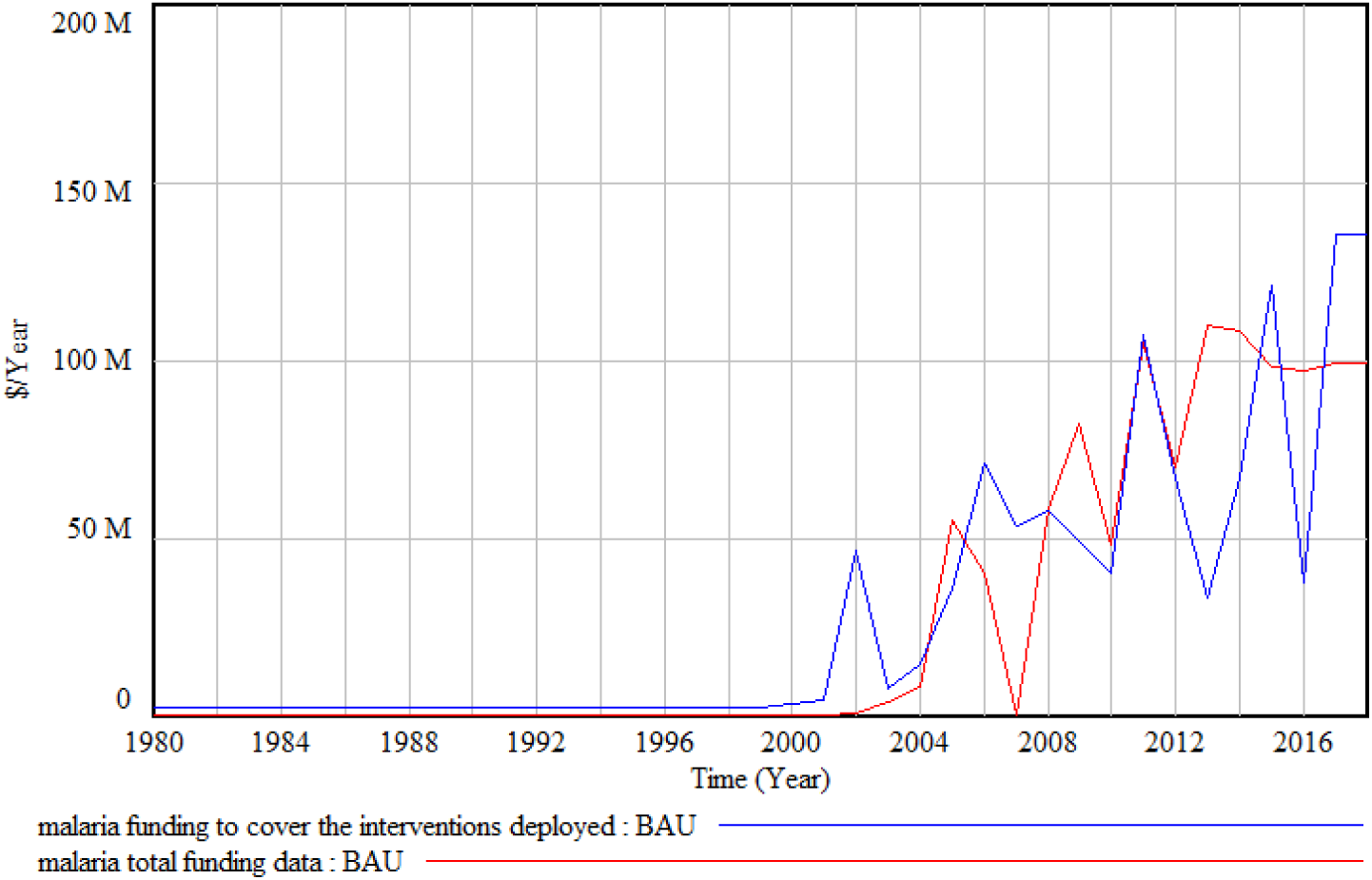
Funding reported vs. funding needed for implemented interventions.

**Figure 16:**
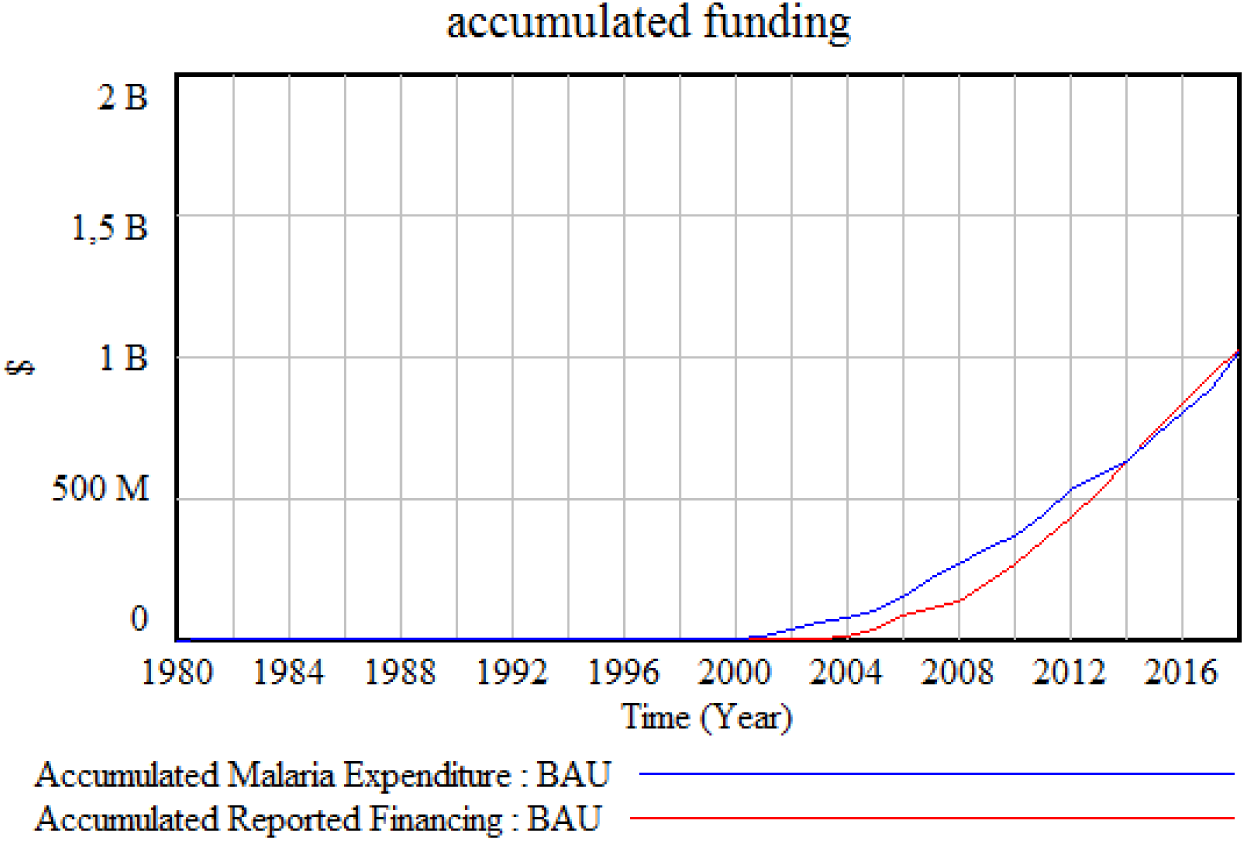
Accumulated Funding vs. Accumulated cost of interventions

**Figure 17:**
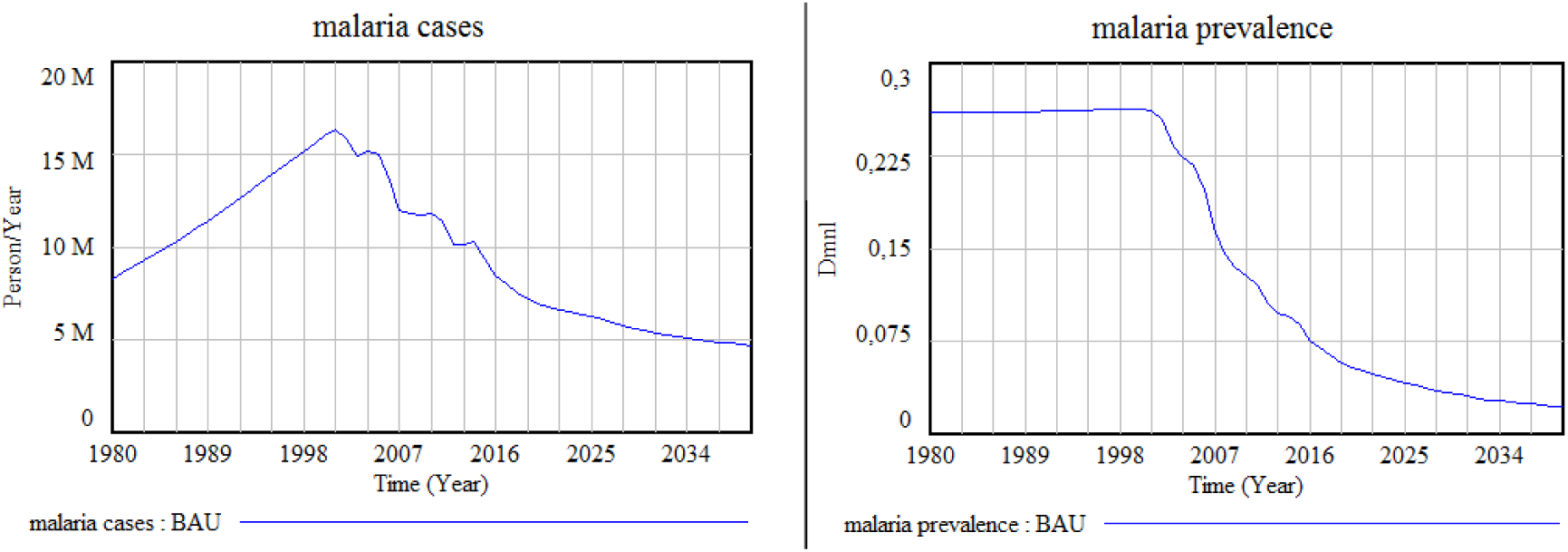
Scenario 1: Constant expenditure at 2 US$/person/year, maintaining the allocation of IVM interventions.

**Figure 18:**
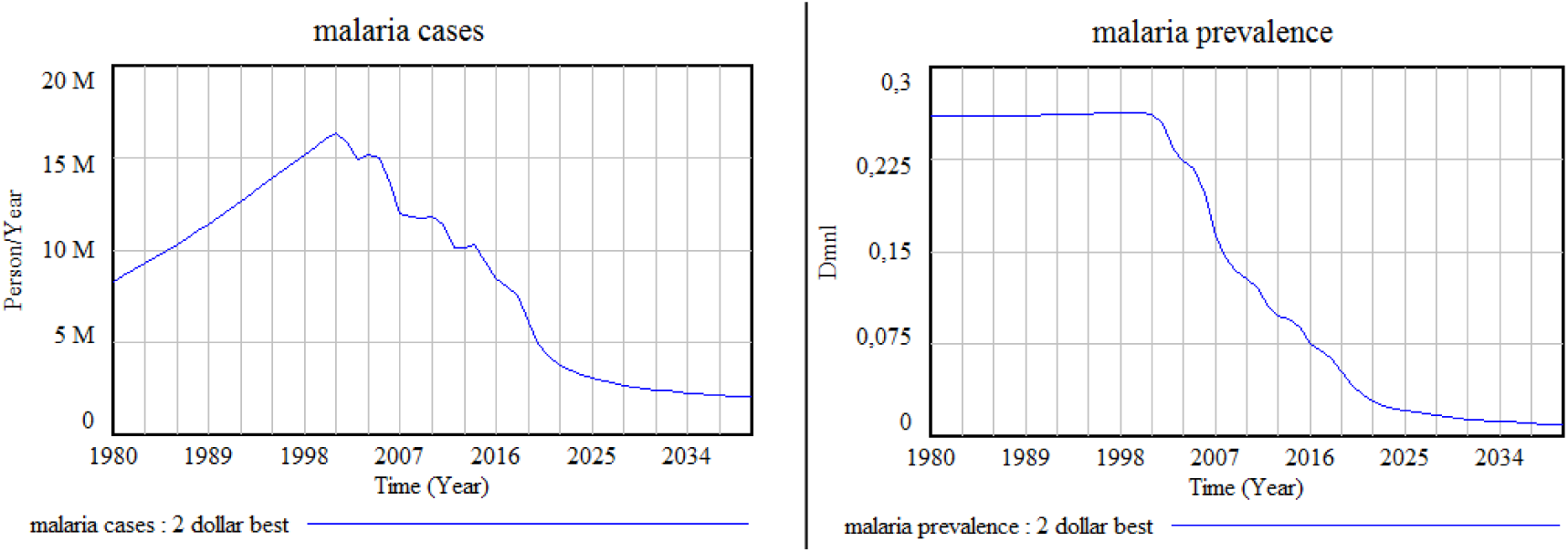
Scenario 2: Constant expenditure at 2 US$/person/year, with an optimal reallocation of the budget for IVM interventions.

There is a certain mismatch between the graphs, although both lines exhibit qualitatively similar increasing paths.

In this case, we believe that underreporting is not the reason why the two graphs do not square, and upon deep examination, the costs of the interventions are in line with our calculations.

In fact, when we sum all the reported funding through the years, and all the costs up to 2018, both numbers match. The next figure represents the accumulated funding, and the accumulated cost of malaria interventions in Kenya:

The graph suggests that either the costs were covered up-front, or donor funding was released earlier than indicated by the reports. In either case, both curves coincide in 2018, indicating that the funding and costs of the interventions are the same.

In summary, the model validation has focused mostly on structural validity tests, especially on direct confirmation of model’s structure and parameters by local malaria experts. In terms of behavioral validation, the results from the model cannot be directly compared with the historical data available for validation purposes. Considering all available information, however, the simulation results provide a more realistic representation of malaria trends than what official data alone would provide. While clearly the availability of more complete and reliable data would improve model accuracy, we believe that the approximation provided by the current model can facilitate the design of effective IVM strategies in Kenya.

Based on the current analysis, the resulting model can also be customized to other geographical areas in order to help decision-makers provide location-specific malaria control policies.

## 6. Cost of Malaria Elimination: Policy Analysis

In this chapter we will use the Kenya Malaria Model (KMM) as the framework to evaluate the long-term impact of different IVM strategies on malaria transmission.

The policies start in 2018, and the goal is to find an optimal solution that could lead Kenya to a state of elimination, or pre-elimination of malaria by 2030. The time horizon of the simulations is 2040, an end-point chosen to investigate whether the implemented policies are sustainable over time.

First, we will portray to the business as usual scenario (BAU), i.e. the current state of malaria interventions in Kenya:

The expenditure to fight malaria in Kenya differs from year to year. During the period 2011-2018, the annual budget has been around 100 Million (constant 2010 US$). When divided by a total population of around 50 million (2018), the approximate per capita expenditure in real terms is on average about 2 US$ per person per year.

The WMR 2017 indicates that the annual per capita expenditure to fight malaria in Kenya was around 1.6 US$ per person during 2014-2016. By adding costs associated with healthcare staff and out-of-pocket expenditure, 2 US$ per person per year is an acceptable estimate, in line with the total budget across the latest years.

However, the intensity of malaria control is not the same across the entire country. The anti-malaria interventions in Kenya are made at the regional level and they depend on the regional epidemiology (See Table 2). Presumably, other conditions in the targeted area are also taken into consideration, such as malaria prevalence, environmental conditions for malaria transmission, or existing coverage prior intervention. In consequence, the interventions are logically tailored to the various endemic regions of the country depending on the malaria risk.

Based on the malaria risk map and the eco-epidemiology of malaria in Kenya, districts have been stratified into 4 regions as follows (Kenya Division of Malaria Control & Ministry of Public Health and Sanitation, 2010):

A. Lake stable endemic and Coast seasonal stable endemic: Risk > 20%
B. Highland epidemic-prone districts: Risk 5% - 20%
C. Seasonal low transmission including arid and semiarid districts: Risk < 5%
D. Low risk districts: Risk < 0.1%

The malaria prevalence is actually different depending on the district. For example, the 2007 Kenya Malaria Indicator Survey (page 30) (Kenya Division of Malaria Control et al., 2009) https indicates that there are variations in malaria parasite prevalence across the epidemiological zones of the country. The following prevalence rates correspond to children with less than 5 years of age:

- 16.4% in endemic areas
- 1% in epidemic prone areas
- 1.4% in areas of seasonal malaria transmission (arid and semi-arid lowlands)
- 0.4% in low risk transmission areas

The above values are not accurate, but serve as a guide for the prevalence corresponding to under-5 children (the weakest population group together with pregnant woman).

In this sense, an intelligent distribution of resources would presumably involve allocating the budget proportionally according to the endemicity of the area. Considering also that the policy of GoK is to deliver LLINs and IRS only in endemic areas, we also estimate that with the current situation, more than 85% of the total budget for both IVM and case management goes to endemic zones.

Current policies also assume that the rest of the regions are engaged in maintaining, and, if possible, reducing prevalence in order to avoid any resurgence of malaria, but at a cost corresponding to a small share of the total budget. For regions with low prevalence, surveillance and case management are considered sufficient when combined with EPR, sensitization, and LLINs for the weakest groups of the population.

Although the model includes the tools to allow for it, in our policy scenarios we have not implemented any future variations in rainfall patterns or temperature due to a possible climate change, which could lead to variations of malaria suitability in some regions. Also, the mosquito resistance to IRS and the parasite resistance to current treatments are considered constant in our simulations, and they correspond to the current values elicited from the literature.

Based on such assumptions, the model simulations portray different projections under three policy scenarios, with the objective of assessing how different levels of expenditure and different resource allocations can impact on projected malaria cases, prevalence, and deaths.

### 6.1. Scenario 1: Business as Usual

In the first projection, the annual per capita expenditure to fight malaria is fixed at the current value of 2 US$. We assume that the budget prioritizes treatment coverage for case management, and the remainder is invested in preventive methods, maintaining the same resource allocation share as in 2017.

In this sense, the preventive measures are distributed as follows: Bed nets or LLINs (75%), spraying or IRS (5%), Environmental Management (8%), and sensitization (12%). Projections under these assumptions indicate a slow decrease of the malaria cases over time, yet maintaining a numbers of over 5 million cases annually by 2030 unless additional interventions are implemented. Malaria prevalence would go down to 3% by 2030, and 2% by 2040.

The graphs below display the results for malaria cases and prevalence in the BAU scenario until 2040:

The estimated IVM coverage is kept roughly constant with a value around 60%, and the simulation also indicates a reduction in malaria mortality by 2030, with approximately 10000 deaths annually in a projected population of 64 million inhabitants.

### 6.2. Scenario 2: Constant Budged and reallocation of IVM interventions

In our second scenario, we also assume a constant per capita expenditure of 2 US$ per person per year, but with an optimal reallocation of the budget for prevention to minimize malaria prevalence by 2030.

Estimations indicate that bed net universal coverage has already been reached in 2017 for everybody living in malaria risk areas. Therefore, the policy should be such that the net delivery only aims at maintaining the coverage with a substitution policy, and taking into account that the lifespan of a net is around 3 years. Given that the net use is still below 70%, the policy should also encourage sensitization campaigns aimed at increasing the efficacy of bed net ownership.

As mentioned before, environmental management (EM) is particularly cost-effective in densely populated areas. Given the current costs and the low coverage of EM by 2017, it seems more profitable to invest in EM than in spraying operations (IRS).

According to the model, the optimal IVM policy in this scenario would consist of increasing investments on EM while keeping the LLIN coverage, and enhance sensitization campaigns. The preventive measures would be distributed as follows: LLINs (44%), IRS (0%), EM (12%), and sensitization (44%).

The results from this policy indicate a more rapid decrease in malaria prevalence with exactly the same costs as the BAU scenario. More precisely, the number of malaria cases by 2030 would be less than 2.5 million, which is half the number reported in the BAU scenario. The malaria prevalence would become 2% in 2030, and less than 1% in 2040.

The graphs below display the results for malaria cases and prevalence in scenario 2:

In this scenario, the estimated IVM coverage in 2030 would be more than 80%, and the annual deaths by malaria would be reduced to around 4000 by 2030, and 3000 by 2040.

In addition, the malaria reduction resulting from this policy would imply an increase in GDP, which, by 2030, would amount to around 1800 million US$ more than the GDP under the BAU scenario. (In the next chapter, we will explore how malaria affects the economy of a country such as Kenya).

### 6.3. Scenario 3: Budged increase and optimal reallocation of IVM interventions

In our third scenario, we assume an increase in the per capita expenditure up to 5 US$ per person per year and the following reallocation of the budget for prevention: LLINs (17%), IRS (46%), EM (20%), and sensitization (17%).

Such shift of resources from LLINs to IRS with respect to our second scenario reflects the fact that 17% of the budget is sufficient to maintain virtually universal coverage of LLINs, and thus more resources can be used for IRS. The expenditure for EM is also increased, but merely up to 20% of the total budget because increases in the EM expenditure are characterized by diminishing returns; it is assumed that EM interventions are implemented first and foremost in very densely populated areas, where interventions affect a larger number of people and are more cost-effective. Beyond 20%, the unit efficacy of EM expenditure would fall below current levels of IRS profitability in terms of malaria reduction.

As illustrated in the Figure 19, in this scenario malaria is pre-eliminated by 2030, with sustainable results through 2040. However, as indicated earlier on, this scenario does not consider the possible increase in resistance to current IRS products, which could indeed reduce IRS effectiveness. For that reason, a larger share of investment towards EM would be a more effective and safe choice.

**Figure 19:**
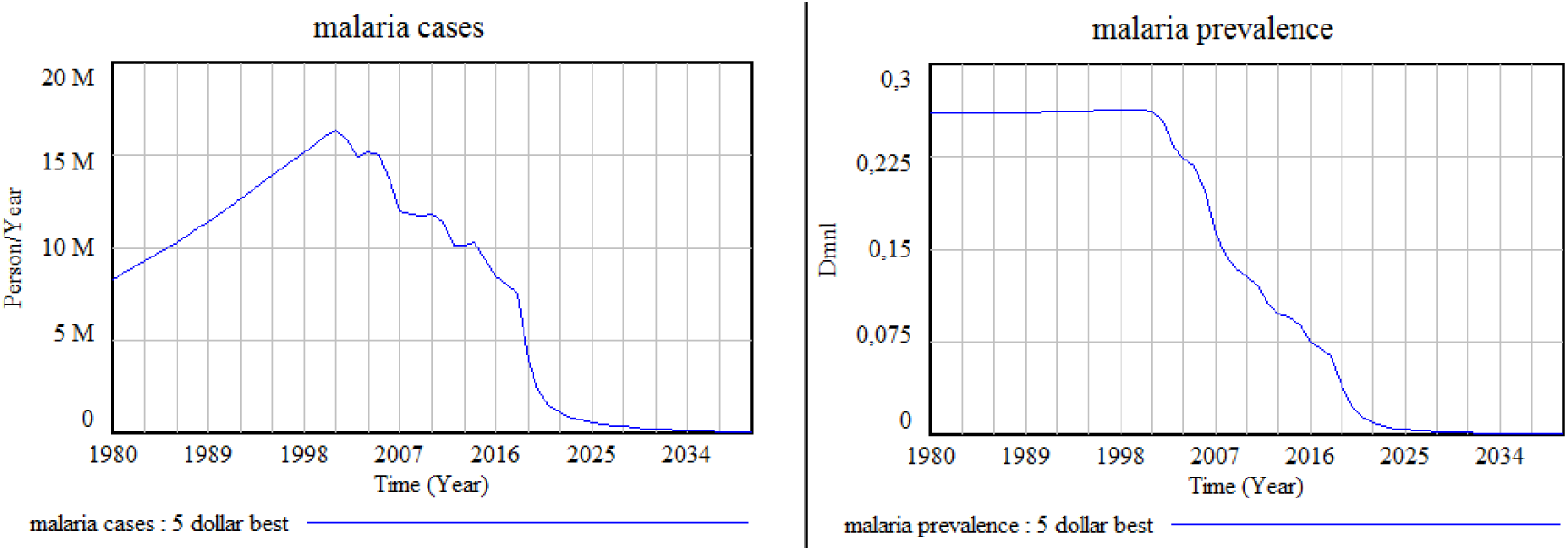
Scenario 3: Scaling up to 5 US$/person/year, with an optimal reallocation of the budget for IVM interventions.

Under this scenario the estimated IVM coverage in 2030 for people living in malaria risk areas would be nearly 100%, and, as observed in the graphs below, the number of cases and prevalence would become marginal.

Figure 20 provides a direct visual comparison of the impact of the three scenarios discussed, on malaria cases and prevalence. While the BAU scenario results in a stabilization at 5 million malaria cases in the long term, in scenario 2 the number of cases is less than half of that, and in scenario 3 the number of cases by 2030 is close to zero.

**Figure 20:**
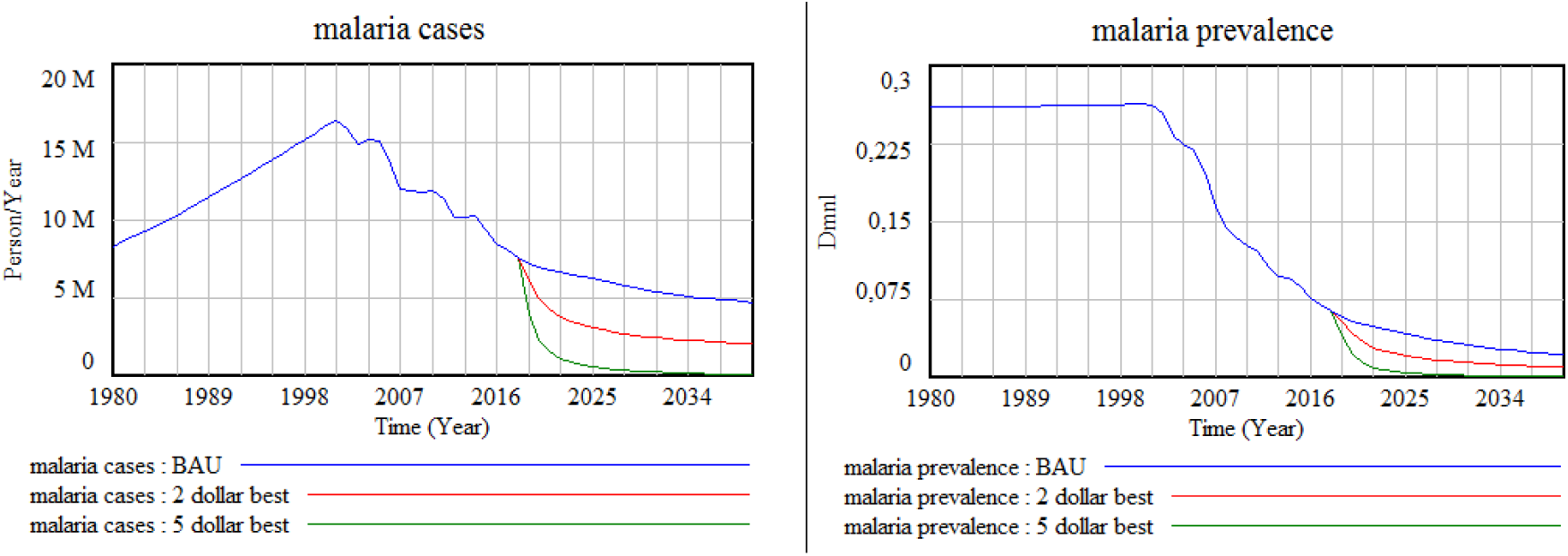
Comparison of scenario results for malaria cases and prevalence.

The same applies to the malaria prevalence and the number of deaths by malaria, which in scenario 3 have virtually been reduced to zero. These results indicate that both an overall budget increase and a reallocation of budget across interventions is needed to achieve malaria elimination over the coming decade.

Figure 21, below, indicates the proportion of the population at risk efficiently covered with IVM under the different scenarios:

**Figure 21:**
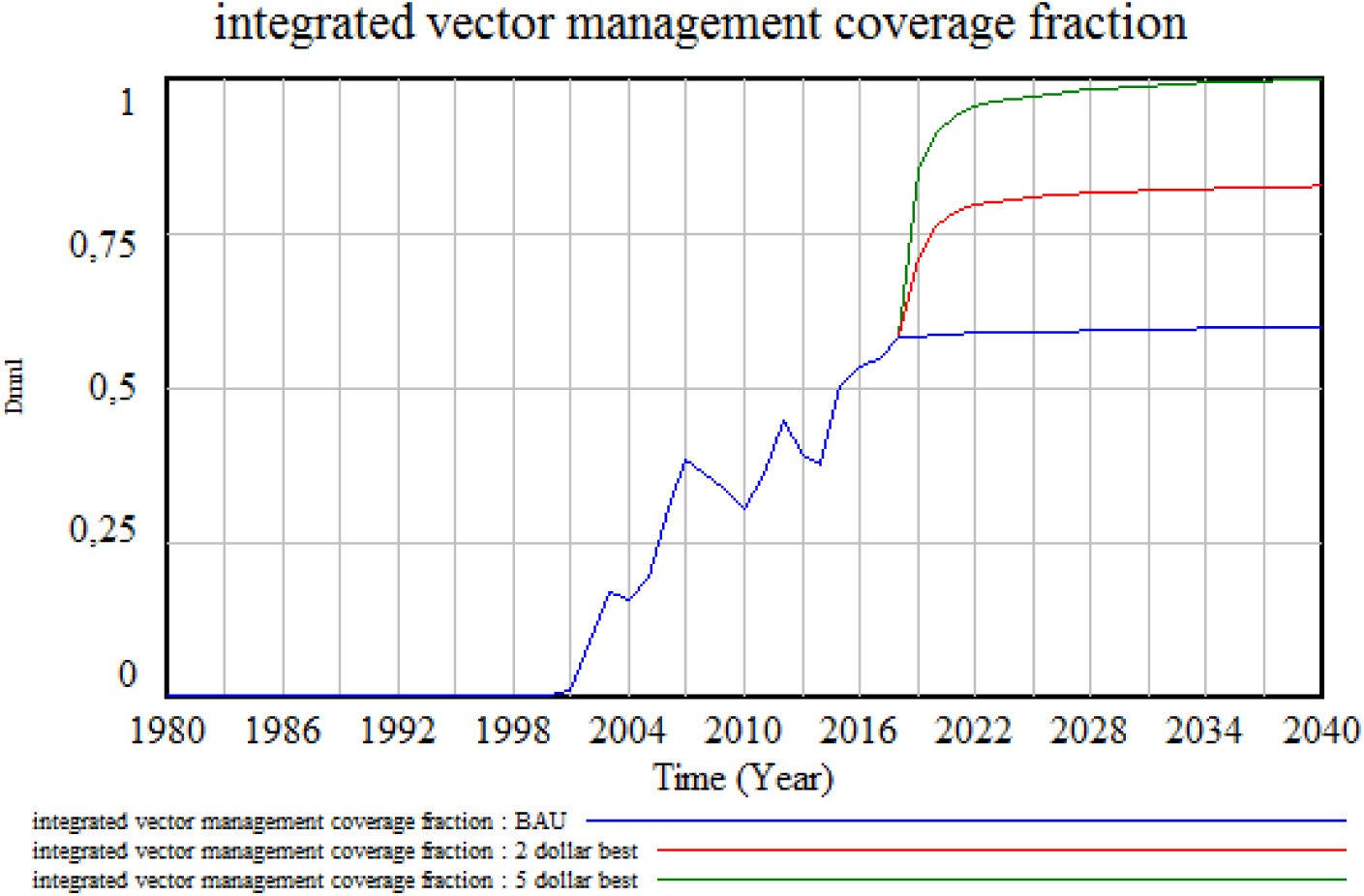
Comparison of scenario results for IVM coverage.

Figure 22, below, indicates the number of children under-5 who would die annually under the different scenarios:

**Figure 22:**
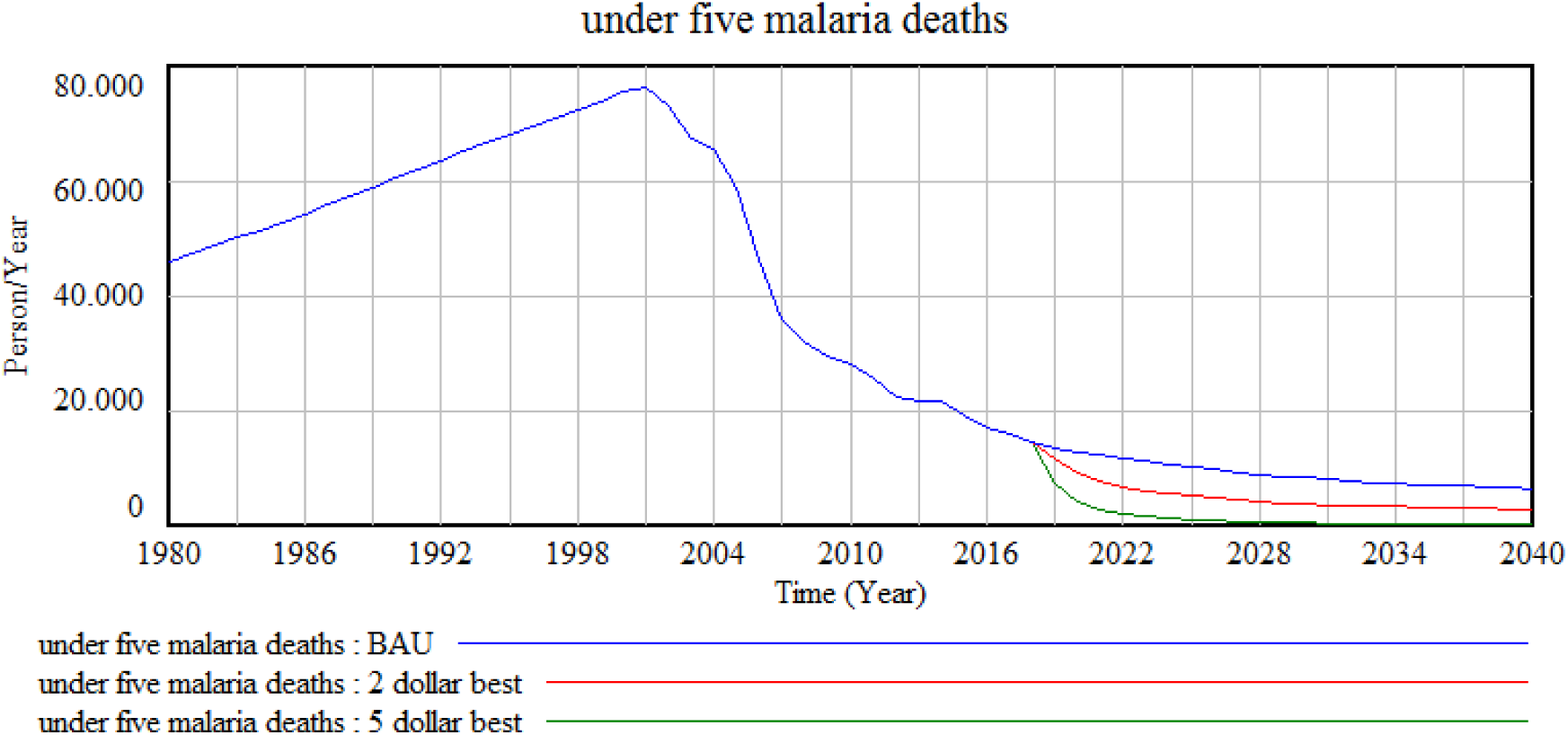
Comparison of scenario results for under-5 malaria deaths.

**Figure 23:**
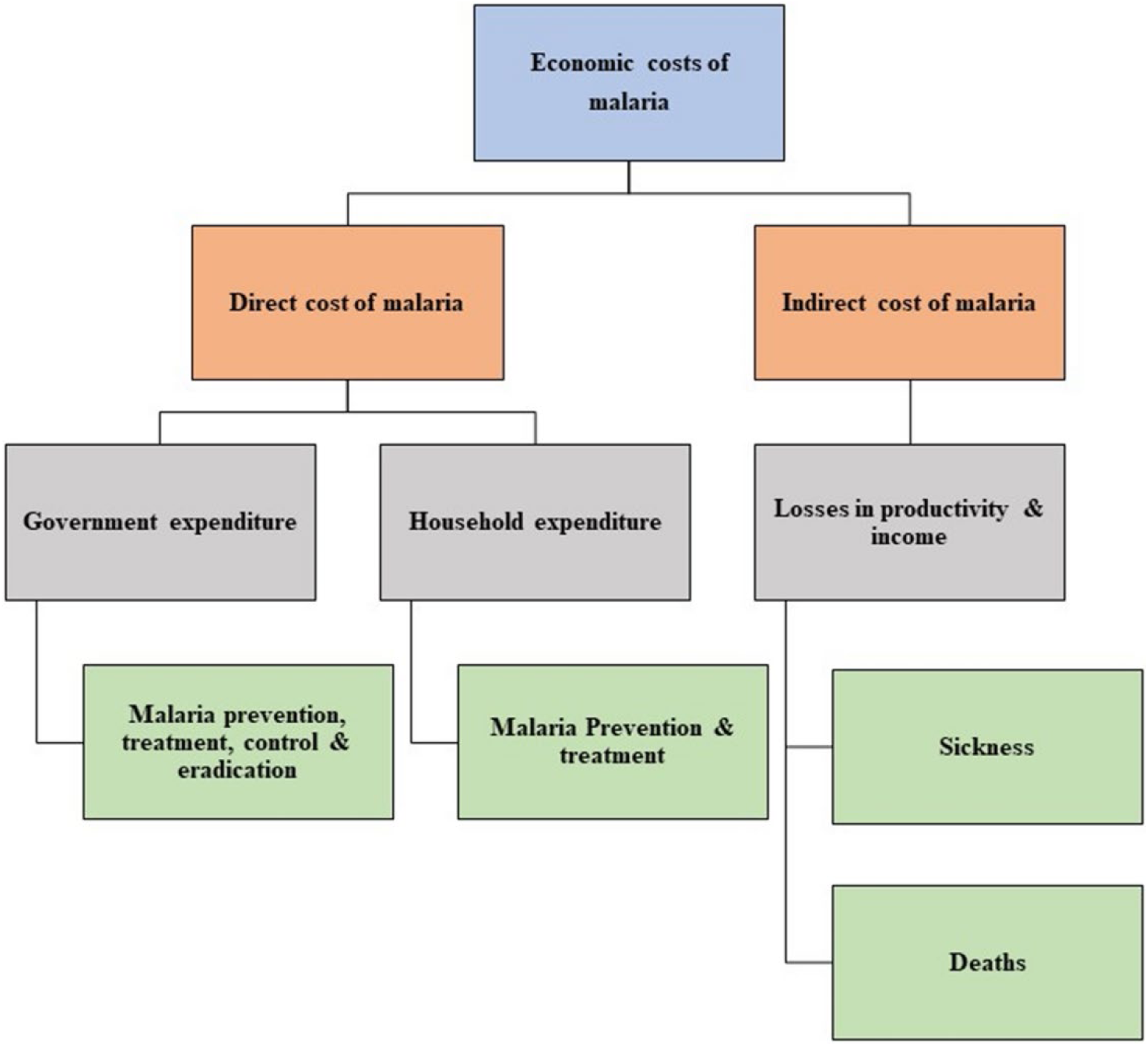
Economic costs of malaria

**Figure 24:**
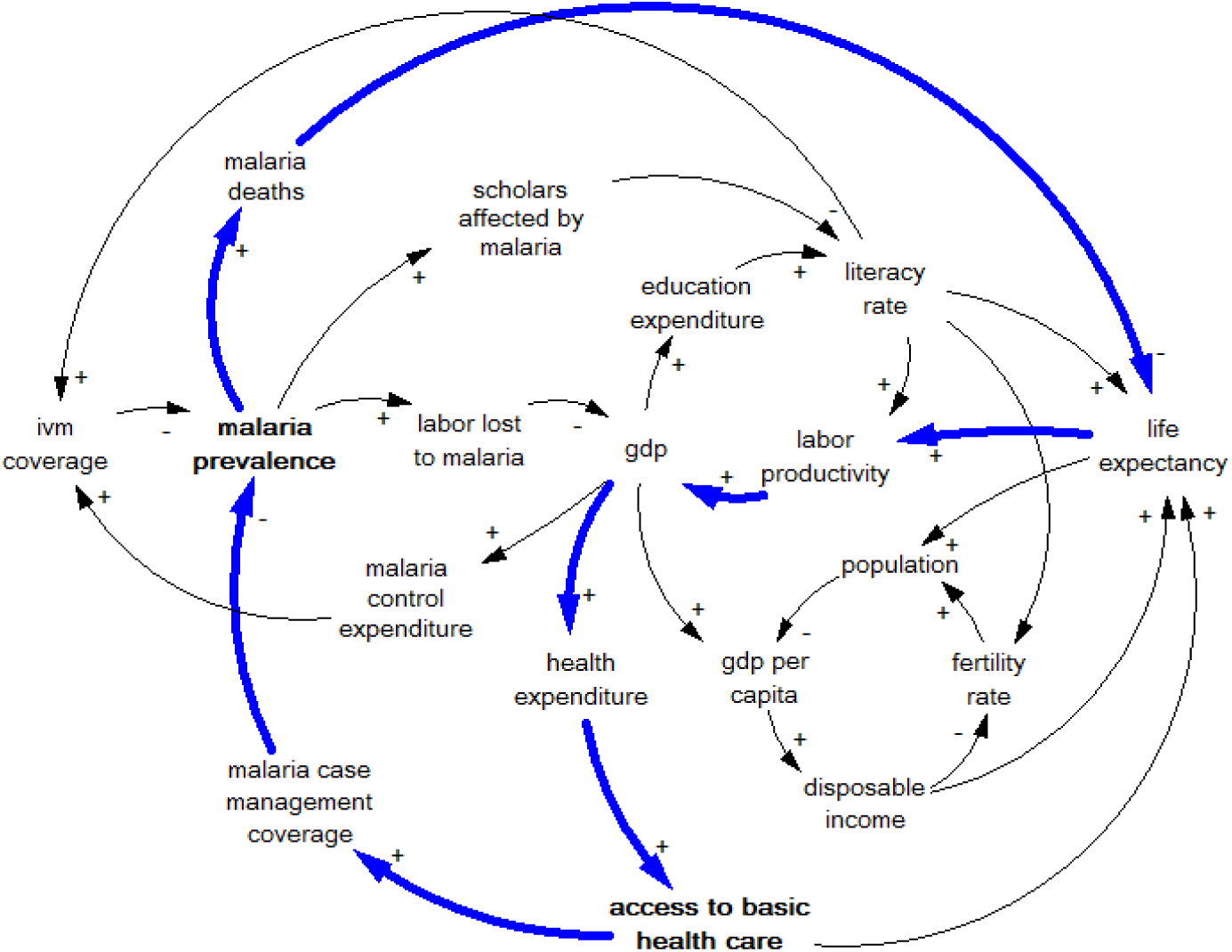
Feedback loop of malaria affecting Healthcare.

**Figure 25:**
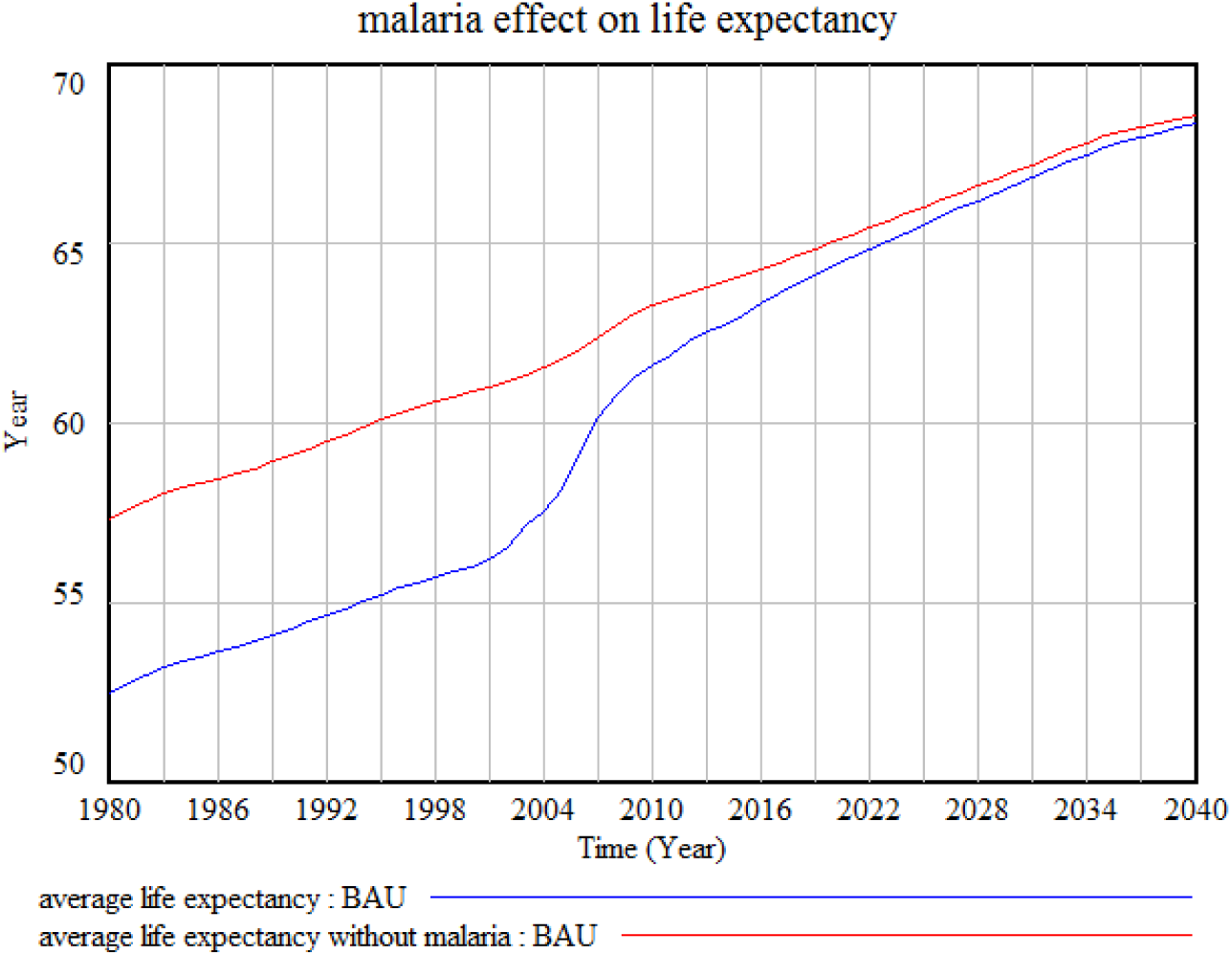
Average life expectancy in Kenya with and without malaria.

**Figure 26:**
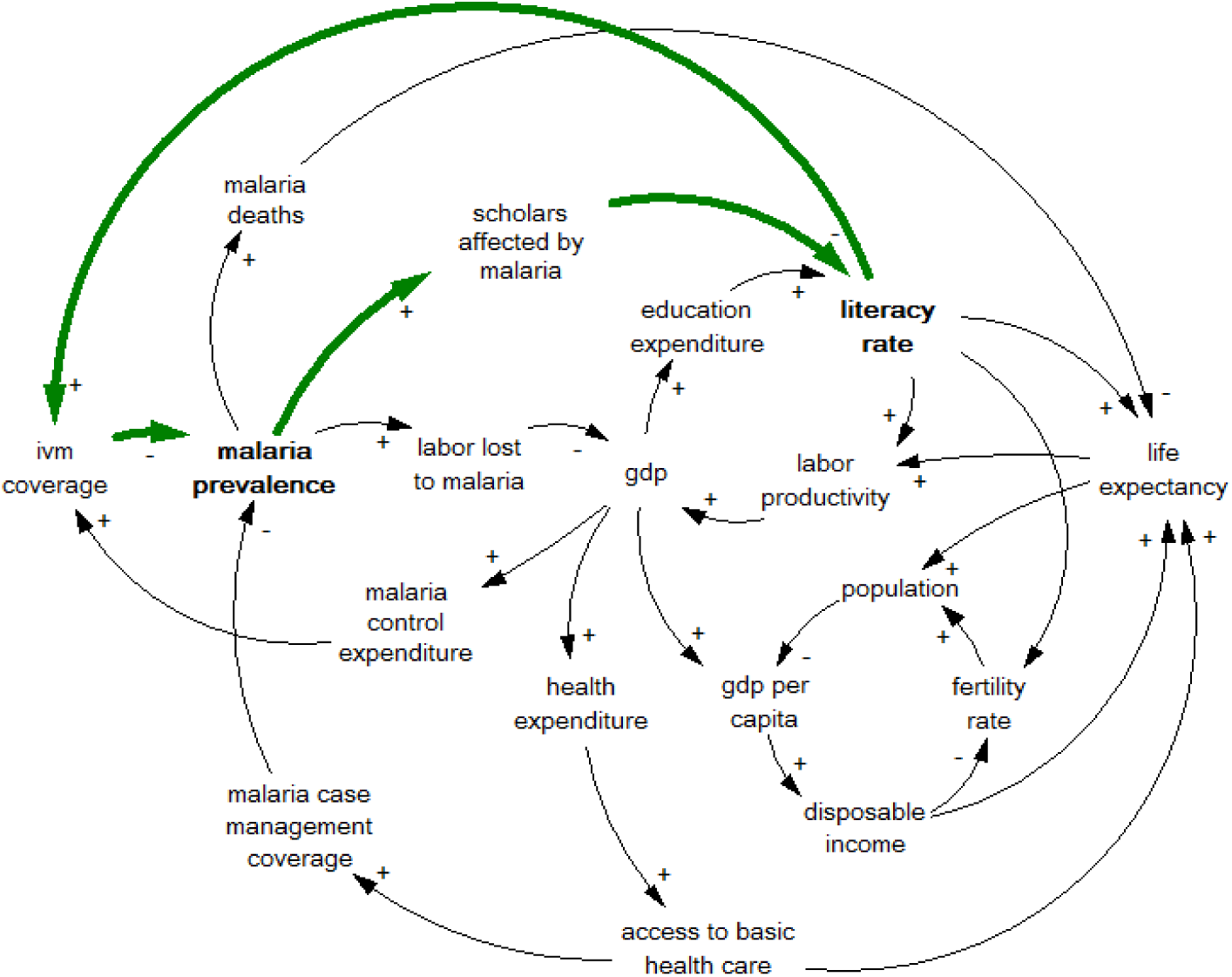
Feedback loop of malaria affecting literacy rate.

**Figure 27:**
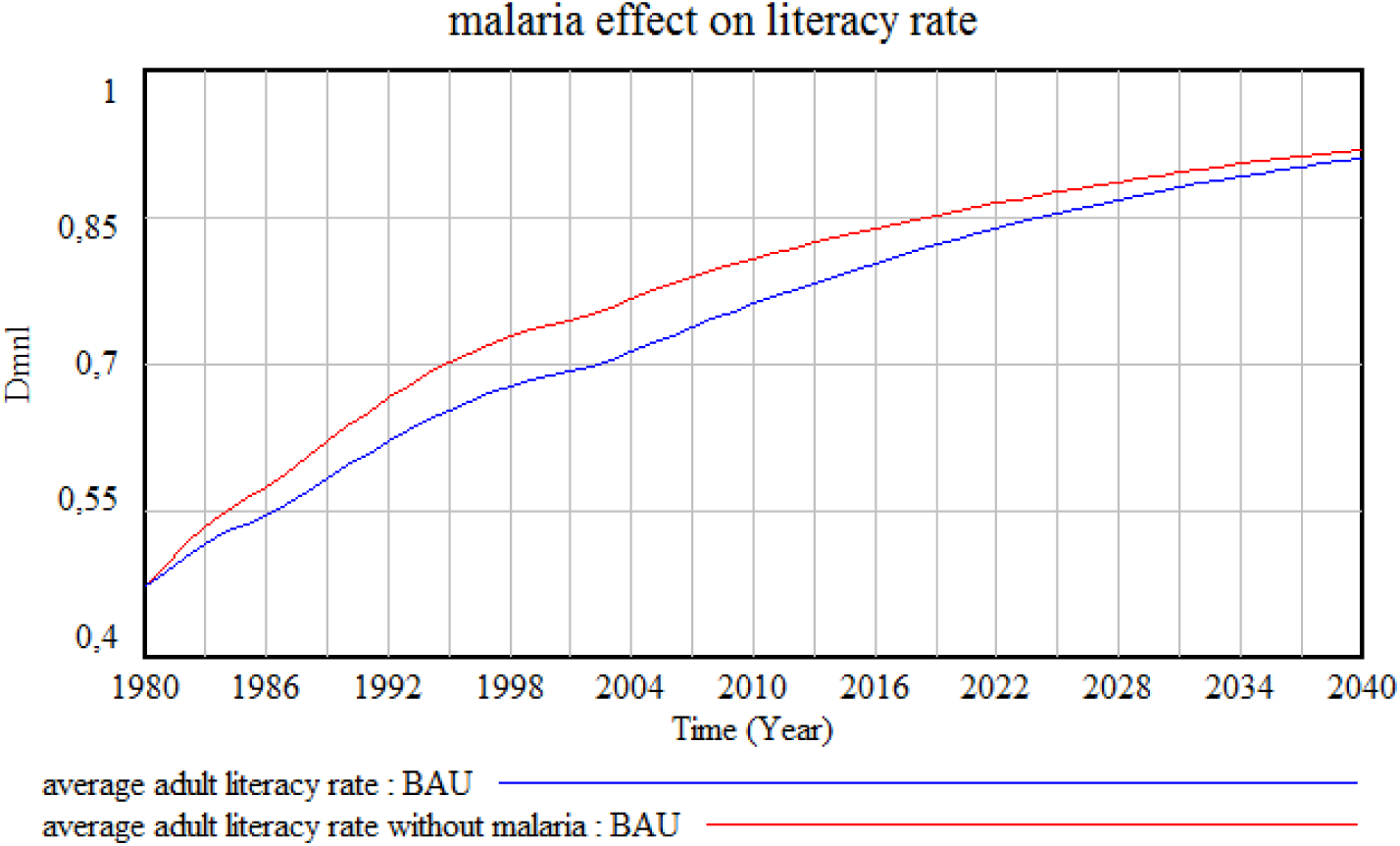
Literacy rate in Kenya with and without malaria.

**Figure 28:**
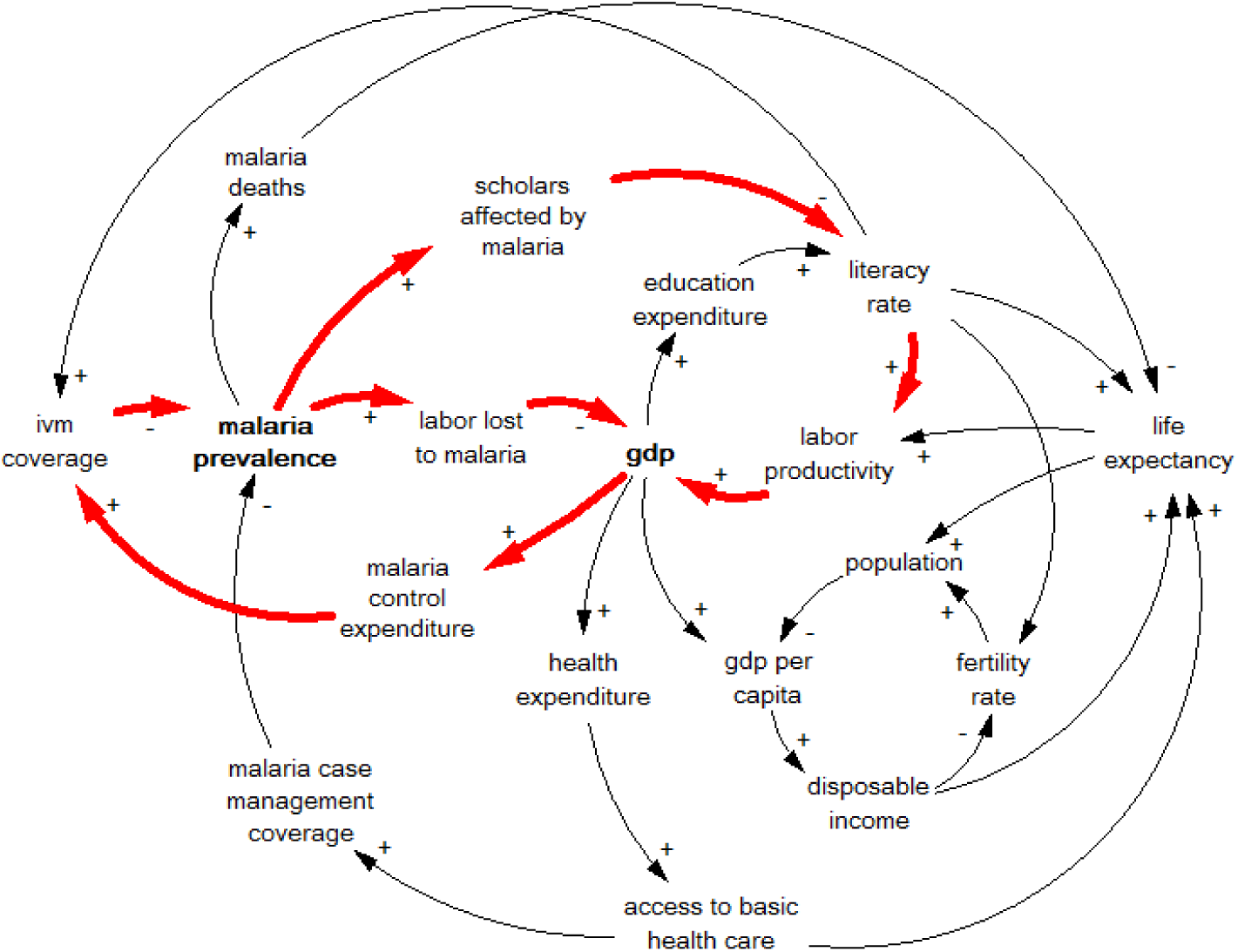
Feedback loop of malaria affecting GDP.

**Figure 29:**
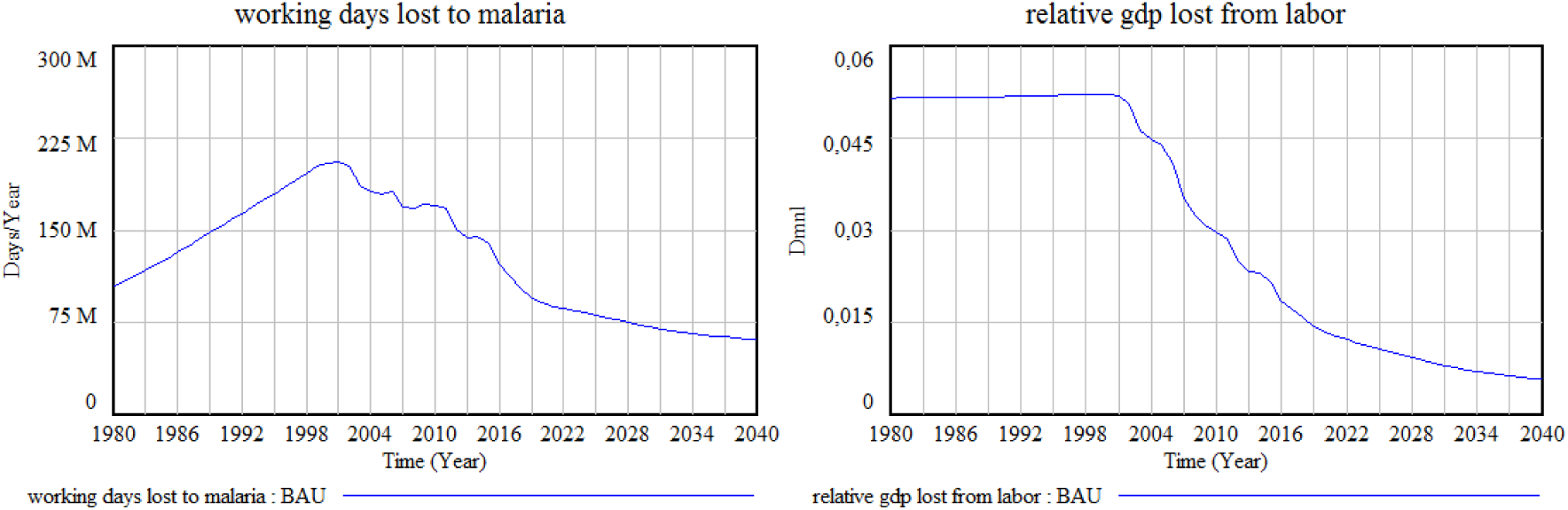
Working days lost to malaria, and GDP fraction lost from malaria afflicted labor.

In summary, the model simulations indicate that for net protection, optimal results are achieved when the LLIN distribution provides universal coverage, i.e. one bed net per two persons. Additional expenditure in LLINs would not provide substantial additional protection, and thus budget resources should be directed to increase the net use or other types of interventions. In this sense, EM is a highly cost-effective strategy in densely populated areas, while its profitability tends to decrease in less populated areas. IRS interventions do not exhibit such diminishing returns, although their effectiveness is strongly linked to the possible emergence of mosquito resistance to IRS products. Therefore, the exact mix of interventions must be determined based on close monitoring of the emergence of resistance, and considering location-specific factors that can render.

## 7. Benefits from malaria elimination

To evaluate the total malaria cost accounting, we must, in addition to considering the expenditures for prevention and case management, balance the bill with the indirect costs of malaria and the possible benefits that would arise from malaria elimination.

The picture below illustrates a breakdown of the economic costs of malaria, which can be classified into direct and indirect costs (Elnour, Grethe, Siddig, & Munga, 2023) https.

Understanding the real costs of malaria in Kenya requires an analysis of its impact on various aspects of economy and society, including education, healthcare, productivity, and overall development. Malaria significantly burdens households through direct medical expenses and indirect costs such as lost workdays and reduced productivity. These costs impede economic growth, exacerbate poverty, and strain public resources (Mezieobi et al., 2025) https.

All these impacts deteriorate the country’s prospects, causing a lack of progress in malaria control capacity, and reinforcing the burden of the disease. To visualize these negative implications, we will explore some of the most relevant feedback loops portrayed in the CLD in Figure 7. That will help us understand the dynamics of malaria in socioeconomic terms, and aid our evaluation of the benefits of a malaria free state in Kenya.

The diagram below highlights (with thick, blue arrows) one of the major feedback loops in the CLD, portraying one of the impacts of malaria prevalence on healthcare:

Taking malaria prevalence as the starting point of the loop, an increase in malaria prevalence would cause an increase of malaria deaths, which, in return reduces life expectancy. A decrease in life expectancy would reduce labor productivity, having a negative impact on GDP production and the corresponding health expenditure as a fraction of the GDP. Cutting health expenditure would reduce the access to basic healthcare and the budget to fight malaria, which ultimately would increase even more the malaria prevalence, closing the loop.

The nature of this loop is reinforcing, meaning that an increase in malaria prevalence leads to even further increases in endemicity.

Indeed, malaria is the cause of many deaths in Kenya. We calculate those deaths by using case fatality rates. Taking as a reference a study done in Ghana (Bawah & Binka, 2008) https, we have analyzed the malaria mortalities, and, by using decrement life-tables, we have estimated the total number of person-years that would have been saved had malaria been eliminated from the population. Using this result we are able to determine the impact of malaria on life expectancy.

The graph below illustrates this impact: The blue curve shows the historical life expectancy in Kenya, and the simulated projection of the future life expectancy under the business as usual (BAU) scenario. (As seen before, BAU refers to the continuation of the current trends and policies in Kenya without any significant changes of interventions, including malaria control policies). The red curve indicates how life expectancy would have developed without malaria.

Before the 2000s, when Kenya had a malaria prevalence of more than 20%, the model indicates that more than 15% of the total deaths were attributable to malaria, and that life expectancy at birth would have likely increased by around 5 years if malaria had been eliminated as a cause of death.

Another important loop that we can distill from the main CLD portrays one of the effects of malaria prevalence on education. The loop is highlighted by thick green arrows:

In this case, malaria prevalence prevents many children in school age from attending school. That compromises the education of the children affected by the disease, and in the long run, the average adult literacy rate. Since the literacy rate is intimately related to the efficacy of malaria interventions, a decrease or stagnation of the literacy rate would ultimately reduce the malaria control coverage, causing even higher levels of malaria prevalence. The nature of this feedback loop is also reinforcing.

We have quantified from the model that, in the year 2000, almost one million of children-years were lost in the schools as a result of malaria. In the graph below, the blue curve shows the historical evolution of the adult literacy rate in Kenya, and the projection of the literacy rate over the future under the BAU scenario, Next to it, the red curve shows how adult literacy rate would have developed without malaria.

Despite of malaria, Kenya is one of the countries with the highest literacy rates in Sub-Saharan Africa. According to Statista https, it was 81.5% in 2018 for individuals aged 15 and older. Had Kenya been a free malaria country after 1980, however, the adult literacy rate would be close to 85% by 2018.

Last but not least, we will elaborate on a feedback loop where malaria is involved in determining the economic growth in Kenya. In the picture below, we observe the loop highlighted with thick red arrows, which in reality is a double loop, where malaria affects GDP through both the literacy rate and the labor force lost to malaria.

On the one hand, and as indicated previously, malaria reduces the school attendance of children, slowing down the development of literacy rate. Such an impact has long-term implications for labor productivity, which affects directly the GDP. On the other hand, malaria also reduces the labor force available for work, compromising GDP as well.

These two negative effects of malaria on GDP impact the funds available to the government in its fight against malaria. A reduction of the budget for malaria control would eventually decrease the IVM coverage, increasing malaria prevalence. In other words, malaria decreases the national economic growth that otherwise would allow a country to adopt more expenditure in health and education services, reducing malaria transmission. This mechanism forms a positive, i.e. reinforcing feedback loop.

As in the previous loops, in this case the literature also includes numbers that can be used to validate the quantitative results from the model:

It is a well-known fact, for instance, that farmers suffering from malaria cultivate 60% less land than their healthy counterparts. Malaria-afflicted farmers lose up to 22 days of work through the illness and harvest only 40.0% of their crops compared to the healthy farmers who could harvest almost 100.0% of their yield [https](https://malariajournal.biomedcentral.com/articles/10.1186/1475-2875-9-91) (Oladepo, Tona, Oshiname, & Titiloye, 2010) [https](https://pmc.ncbi.nlm.nih.gov/articles/PMC2856588/). This affects not only GDP, but also food security, causing a level of undernourishment in the population and, thus, reducing the overall ability to perform agricultural tasks. In this regard, we assume that the 60% reduction in farming productivity integrates the undernourishment as a consequence of the malaria symptoms.

A previous study indicated that countries with an intensive malaria prevalence, grew 1.3% less per person per year, and that a 10% reduction in malaria was associated with 0.3% higher economic growth (Gallup & Sachs, 2000) [https](https://www.ncbi.nlm.nih.gov/books/NBK2624/). Also from the literature, a 10% increase in malaria prevalence reduced the monthly individual wage income by 3.3% to 3.8%. Moreover, the total economic malaria costs constitutes (on average) 1% of total household income. And in Kenya, it was estimated that 170 million working days were lost due to the disease each year (MOH 2001) (Elnour et al., 2023) [https](https://www.ncbi.nlm.nih.gov/books/NBK2624/).

Considering approximately 230 working days per person per year, the 170 million working days would correspond to the equivalent of 740000 workers lost to malaria every year. In our model, the simulated working age population in 2001 is around 16.5 million. Assuming a 67% labor participation rate in Kenya https, the total labor supply would be about 11 million people. Then, 740000 workers would constitute nearly 7% of the entire workforce, which is in line with the fraction of labor lost to malaria obtained in the model.

Under these conditions, malaria caused an economic loss due to the reduction of work days of more than 5% of the GDP, which is almost 12 times the budget destined to fight malaria nowadays^5^.

The figure below shows both the simulated working days lost to malaria, and the fraction of the GDP lost from the resulting reduction of workable days, under the BAU scenario:

This calculation takes into account only the working days lost. If we consider additional effects in GDP from other factors such as life expectancy or literacy rate, the impact is even greater. However, we also need to consider that malaria does not affect all income groups in the same manner; in this sense, the estimate above is based on the assumption that all individuals have the same probability of getting malaria, but in reality, malaria is strongly associated with poverty, meaning that malaria infected population normally corresponds to lower income cohorts.

In summary, the direct and indirect economic costs of malaria strain healthcare systems, increase household expenditures, and deepen the financial instability among the affected populations, thereby perpetuating vicious cycles of poverty (Mezieobi et al., 2025). On the contrary, beyond direct health benefits, malaria elimination in Kenya would imply a significant increase in the average labor productivity, not to mention the positive impacts on important social variables, such as life expectancy and children attendance at school, which should be taken into account when it comes to evaluate IVM strategies.

## 8. Conclusions

Much improvement has been accomplished in Kenya with regard to malaria control programs, but more investigation is required to assess whether the implemented policy frameworks will cause the desired malaria reduction over the coming years. Based on the preliminary analysis, carried out using the Kenya Malaria Model (KMM), it appears that both, a substantial increase in malaria budget and a reallocation of such budget across interventions, is necessary in order to achieve malaria elimination in 2030.

By maintaining the current malaria programs, our model estimates that malaria prevalence would decrease to 3% by 2030, and tend to stabilize at about 2% by 2040. But when it comes to elimination, more funding would be needed: Our simulations indicate that elimination could be achieved over the next decade by increasing the current budget to 5 US$/person/year.

In terms of budget allocation across interventions, the study considered fundamentally the implementation of the four most common types of protective interventions and their possible combination by means of Integrated Vector Management (IVM). These interventions are; long-lasting insecticide-treated nets (LLINs), indoor residual spraying (IRS), environmental management (EM), and malaria sensitization or social behavior change.

Regarding the first type of operation, since 2016 nearly everybody living in malaria risk areas has access to a bed net, and estimations indicate that the goal of universal coverage, defined as one LLIN for every two persons at risk of malaria, has been achieved. With universal coverage established, the question remains whether it is necessary to increase coverage to one LLIN per person, or efforts should move towards increasing and improving people’s bed net usage i.e. appropriate use to increase the protective effectiveness, and towards a more intensive use of other types of malaria control interventions.

Since malaria remains a leading cause of illness and death in Kenya, it is also a source of cycles that hinder the country’s socioeconomic development prospects. By reducing malaria prevalence, the country benefits from increases in indicators such as GDP, literacy rate, and life expectancy, among others. Moreover, malaria elimination in Kenya would generate sufficient economic benefits to cover the costs to fight the disease.

Further steps in the current research would elaborate on scenarios with mass distribution vaccines, or the effect of a possible climate change on malaria transmission.

## Supporting information

Supporting information: Model, Equations and Data

## Data Availability

All data produced in the present study are available in the links provided in the manuscript, in the manuscript itself, or upon reasonable request to the authors.

## Footnotes

1 Malaria sensitization refers to raising awareness and educating communities about malaria, its prevention, and treatment. This involves sharing information on symptoms, prevention methods, and the importance of seeking prompt medical care. Sensitization campaigns aim to empower individuals and communities to take ownership of malaria control and prevention efforts (WHO). Social behavior change (SBC) or social and behavior change communication (SBCC) is a strategic approach focused on understanding and influencing the factors that drive behaviors related to malaria, promoting positive behaviors related to prevention, testing, and treatment (WHO). For simplification purposes, we merge sensitization and SBC as the same type of intervention.

2 The entomological inoculation rate is the average number of inoculations with malaria parasites received by a person over a period of time (usually annually). It is used to measure malaria transmission intensity and is dependent on the frequency with which people living in an area are bitten by anopheline mosquitoes carrying sporozoites (WHO 2015).

3 Note that the table includes IPTp and EPR, which are not discussed in the paper. IPTp consist of intermittent preventive treatment of malaria for pregnant women, only delivered in endemic areas. Epidemic Preparedness and Response (EPR) is an approach intended to improve epidemic preparedness and response by establishment of malaria early warning systems and carrying out preventive measures such as IRS campaigns in order to avoid epidemics. EPR is useless in endemic areas since malaria transmission is present during the whole year. BCC stands for Behavior Change Communication.

4 WMR 2008: “Kenya had an estimated 11.3 million malaria cases in 2006. The majority of cases are caused by P. falciparum but most are unconfirmed. The number of reported cases increased in 4 out of 5 years between 2001 and 2006; it is not known whether this represents improved reporting or an increase in incidence.” WMR 2009: “Kenya had an estimated 15 million malaria cases in 2006. The majority are due to P. falciparum. Almost all the reported 9 million suspected malaria cases in 2007 were unconfirmed. The number of reported cases increased between 2001 and 2007; it is not known whether this represents improved reporting or an increase in incidence.”

5 The GDP 2001 in Kenya was = 23400 million (constant 2010 US$), and the malaria expenditure as per 2018 was around 100 million (constant 2010 US$). According to the model, the economic loss due to the reduction of workable days is (relative GDP lost from labor) = 5.2% of GDP 2001 = 1170 million. This value is 11.7 times bigger than the total malaria expenditure.

## References

1. Abeku, T., Helinski, M., Kirby, M., Kefyalew, T., Awano, T., Batisso, E., … Meek, S. (2015). Monitoring changes in malaria epidemiology and effectiveness of interventions in Ethiopia and Uganda: Beyond Garki Project baseline survey. Malaria Journal, 14, 337. doi:10.1186/s12936-015-0852-7

2. Awosolu, O. B., Yahaya, Z. S., & Farah Haziqah, M. T. (2021). Prevalence, Parasite Density and Determinants of Falciparum Malaria Among Febrile Children in Some Peri-Urban Communities in Southwestern Nigeria: A Cross-Sectional Study. Infect Drug Resist, 14, 3219–3232. doi:10.2147/idr.S312519

3. Barlas, Y. (1996). Formal aspects of model validity and validation in system dynamics. System Dynamics Review, 12(3), 183–210. doi:10.1002/(sici)1099-1727(199623)12:3<183::Aid-sdr103>3.0.Co;2-4

4. Bawah, A., & Binka, F. (2008). How Many Years of Life Could Be Saved If Malaria Were Eliminated from a Hyperendemic Area of Northern Ghana? The American journal of tropical medicine and hygiene, 77, 145–152. doi:10.4269/ajtmh.2007.77.145

5. Beier, J. C., Keating, J., Githure, J. I., Macdonald, M. B., Impoinvil, D. E., & Novak, R. J. (2008). Integrated vector management for malaria control. Malar J, 7 *Suppl 1*, S4. doi:10.1186/1475-2875-7-S1-S4

6. Central Bureau of Statistics, C. B. S. K., Ministry of Health, M. O. H. K., & Macro, O. R. C. (2004). Kenya Demographic and Health Survey 2003. Retrieved from Calverton, Maryland, USA: http://dhsprogram.com/pubs/pdf/FR151/FR151.pdf

7. Elnour, Z., Grethe, H., Siddig, K., & Munga, S. (2023). Malaria control and elimination in Kenya: economy-wide benefits and regional disparities. Malar J, 22(1), 117. doi:10.1186/s12936-023-04505-6

8. Gallup, J., & Sachs, J. (2000). The Economic Burden of Malaria. The American journal of tropical medicine and hygiene, 64, 85–96. doi:10.4269/ajtmh.2001.64.85

9. Hay, S. I., Simba, M., Busolo, M., Noor, A. M., Guyatt, H. L., Ochola, S. A., & Snow, R. W. (2002). Defining and detecting malaria epidemics in the highlands of western Kenya. Emerg Infect Dis, 8(6), 555–562. doi:10.3201/eid0806.010310

10. Kamau, A., Nyaga, V., Bauni, E., Tsofa, B., Noor, A. M., Bejon, P., … Hammitt, L. L. (2017). Trends in bednet ownership and usage, and the effect of bednets on malaria hospitalization in the Kilifi Health and Demographic Surveillance System (KHDSS): 2008-2015. BMC Infect Dis, 17(1), 720. doi:10.1186/s12879-017-2822-x

11. Kenya Division of Malaria Control, & Ministry of Public Health and Sanitation. (2009). National Malaria Strategy 2009-2017. Retrieved from https://www.nationalplanningcycles.org/sites/default/files/country_docs/Kenya/kenya_national_malaria_strategy_2009-2017.pdf

12. Kenya Division of Malaria Control, & Ministry of Public Health and Sanitation. (2010). National Malaria Policy 2010. Retrieved from https://nmcp.or.ke/

13. Kenya Division of Malaria Control, & Ministry of Public Health and Sanitation. (2011). 2010 Kenya Malaria Indicator Survey. Retrieved from https://dhsprogram.com/pubs/pdf/mis7/mis7.pdf

14. Kenya Division of Malaria Control, Ministry of Public Health and Sanitation, Kenya National Bureau of Statistics, & National Coordinating Agency for Population and Development. (2009). 2007 Kenya Malaria Indicator Survey. Retrieved from https://www.malariasurveys.org/documents/KMIS_2007_Consolidated_Apr09-1.pdf

15. Kenya National Bureau of Statistics. (2011). Economic Survey 2011.

16. Kenya National Bureau of Statistics. (2018). Economic Survey 2018.

17. Kibe, L. W., Kamau, A. W., Gachigi, J. K., Habluetzel, A., & Mbogo, C. M. (2019). A formative study of disposal and re-use of old mosquito nets by communities in Malindi, Kenya. MalariaWorld journal, 6, 9–9. Retrieved from https://pubmed.ncbi.nlm.nih.gov/31293898/

18. Kibe, L. W., Mbogo, C. M., Keating, J., Molyneux, S., Githure, J. I., & Beier, J. C. (2006). Community based vector control in Malindi, Kenya. African health sciences, 6(4), 240–246. doi:10.5555/afhs.2006.6.4.240

19. Mbogo, C. M., Mwangangi, J. M., Nzovu, J., Gu, W., Yan, G., Gunter, J. T., … Beier, J. C. (2003). Spatial and temporal heterogeneity of Anopheles mosquitoes and Plasmodium falciparum transmission along the Kenyan coast. Am J Trop Med Hyg, 68(6), 734–742.

20. Mezieobi, K. C., Alum, E. U., Ugwu, O. P., Uti, D. E., Alum, B. N., Egba, S. I., & Ewah, C. M. (2025). Economic burden of malaria on developing countries: A mini review. Parasite Epidemiol Control, 30, e00435. doi:10.1016/j.parepi.2025.e00435

21. National Malaria Control Program, M. o. H. (2017). Post Mass Net Distribution Long Lasting Insecticidal Net Survey 2017. Retrieved from Nairobi, Kenya: http://www.nmcp.or.ke/index.php/resource-centre/download-centre/category/5-surveillance-monitoring-and-evaluation

22. National Malaria Control Programme - NMCP/Kenya, Kenya National Bureau of Statistics - KNBS, & ICF International. (2016). Kenya Malaria Indicator Survey 2015. Retrieved from Nairobi, Kenya: http://dhsprogram.com/pubs/pdf/MIS22/MIS22.pdf

23. National Malaria Control Programme, M. o. H., Kenya. (2019). *Kenya Malaria Strategy 2019-*2023. Retrieved from https://fountainafrica.org/wp-content/uploads/2020/01/Kenya-Malaria-Strategy-2019-2023

24. Noor, A., Macharia, P., Ouma, P., Oloo, S., Maina, J., Gogo, E., … Ejersa, W. (2016). The epidemiology and control profile of malaria in Kenya: reviewing the evidence to guide the future vector control.

25. Noor, A. M., Amin, A. A., Akhwale, W. S., & Snow, R. W. (2007). Increasing Coverage and Decreasing Inequity in Insecticide-Treated Bed Net Use among Rural Kenyan Children. PLOS Medicine, 4(8), e255. doi:10.1371/journal.pmed.0040255

26. Ntuku, H. M., Ruckstuhl, L., Julo-Reminiac, J. E., Umesumbu, S. E., Bokota, A., Tshefu, A. K., & Lengeler, C. (2017). Long-lasting insecticidal net (LLIN) ownership, use and cost of implementation after a mass distribution campaign in Kasai Occidental Province, Democratic Republic of Congo. Malar J, 16(1), 22. doi:10.1186/s12936-016-1671-1

27. Oladepo, O., Tona, G. O., Oshiname, F. O., & Titiloye, M. A. (2010). Malaria knowledge and agricultural practices that promote mosquito breeding in two rural farming communities in Oyo State, Nigeria. Malar J, 9, 91. doi:10.1186/1475-2875-9-91

28. Pedercini, M., Movilla Blanco, S., & Kopainsky, B. (2011). Application of the malaria management model to the analysis of costs and benefits of DDT versus non-DDT malaria control. PLoS One, 6(11), e27771. doi:10.1371/journal.pone.0027771

29. Ranjha, R., Singh, K., Baharia, R. K., Mohan, M., Anvikar, A. R., & Bharti, P. K. (2023). Age-specific malaria vulnerability and transmission reservoir among children. Global Pediatrics, 6, 100085. 10.1016/j.gpeds.2023.100085

30. Roll Back Malaria Partnership. (2008). The Global Malaria Action Plan for a Malaria-free World. Retrieved from Geneva: The Roll Back Malaria Partnership.: https://www.afro.who.int/sites/default/files/2017-06/Gmapfull.pdf

31. Small, J., Goetz, S. J., & Hay, S. I. (2003). Climatic suitability for malaria transmission in Africa, 1911-1995. Proc Natl Acad Sci U S A, 100(26), 15341–15345. doi:10.1073/pnas.2236969100

32. Snow, R. W., Kibuchi, E., Karuri, S. W., Sang, G., Gitonga, C. W., Mwandawiro, C., … Noor, A. M. (2015). Changing Malaria Prevalence on the Kenyan Coast since 1974: Climate, Drugs and Vector Control. PLoS One, 10(6), e0128792. doi:10.1371/journal.pone.0128792

33. Sterman, J. D. (2000). Business dynamics. Systems thinking and modeling for a complex world. Boston et. al.: Irwin McGraw-Hill.

34. Temperley, M., Mueller, D. H., Njagi, J. K., Akhwale, W., Clarke, S. E., Jukes, M. C. H., … Brooker, S. (2008). Costs and cost-effectiveness of delivering intermittent preventive treatment through schools in western Kenya. Malaria Journal, 7(1), 196. doi:10.1186/1475-2875-7-196

35. WHO. (2011). World Malaria Report 2011. Retrieved from https://www.who.int/publications/i/item/9789241564403

36. WHO. (2018a). World Malaria Report 2018. Geneva: World Health Organization.

37. WHO. (2018b). World Malaria Report 2018 (9789241565653). Retrieved from https://apps.who.int/iris/bitstream/handle/10665/275867/9789241565653-eng.pdf

38. World Health Organization. Malaria, U. (2005). Malaria control in complex emergencies : an inter-agency field handbook / World Health Organization … et al.. In. Geneva: World Health Organization.

