## Supporting information: Model, Equations and Data for "Costs and Benefits of Malaria Elimination in Kenya by Means of IVM Implementation": Complete list of mathematical model equations.pdf

### APPENDIX

#### Complete list of mathematical model equations:

Below we list all the equations used in the Kenya Malaria Model. Initial values and parameter values are those from the baseline simulation.

********************************
arrays
********************************

age: UNDER FIVE,FIVE AND OVER

region: CENTRAL,COAST,EASTERN,NAIROBI,NORTH EASTERN,NYANZA,RIFT VALLEY,WESTERN

sex: FEMALE, MALE

********************************
Simulation control parameters
********************************

FINAL TIME = 2040
Units: Year

INITIAL TIME = 1980
Units: Year

TIME STEP = 0.0625
Units: Year

SAVEPER = 1
Units: Year

********************************
1 Population
********************************

AVERAGE life expectancy:INTERPOLATE:
Units: Year

average life expectancy 0=
average life expectancy without malaria*effect of proportion of deaths due to malaria on life expectancy
-effect of exposure to malaria on lifespan
Units: Year

average life expectancy without malaria=
EFFECT OF WELLBEING INDEX ON AVERAGE LIFE EXPECTANCY(wellbeing index)
Units: Year

becoming elderly=
Working Age Population/WORKING AGE DURATION
Units: Person/Year

becoming school age=
Infant Population/INFANT STAGE DURATION
Units: Person/Year

becoming working age=
School Age Population/SCHOOL AGE DURATION
Units: Person/Year

births=
childbearing female population*total fertility rate/FERTILE PERIOD
Units: Person/Year

childbearing female population=
Working Age Population*PROPORTION OF CHILDBEARING FEMALE IN WORKING AGE POPULATION
Units: Person

contraceptive prevalence=
INITIAL CONTRACEPTIVE PREVALENCE*(relative average adult literacy rate^ELASTICITY OF CONTRACEPTIVE PREVALENCE TO LITERACY RATE)
Units: Dmnl

desired fertility rate=
EFFECT OF INCOME ON DESIRED FERTILITY RATE(relative perceived pc real disposable income)
Units: Dmnl

EFFECT OF CONTRACEPTIVE PREVALENCE ON TOTAL FERTILITY RATE(
[(0.1,0)-(1,3)],(0.1,2.2),(0.25,1.8),(0.4,1.5),(0.6,1.26),(0.8,1.1),(1,1))
Units: Dmnl

effect of exposure to malaria on lifespan= WITH LOOKUP (
malaria prevalence,
([(0,0)-(1,1)],(0,0),(1,1) ))
Units: Year

EFFECT OF INCOME ON DESIRED FERTILITY RATE(
[(0.4,2)-(3,5)],(0.4,5),(0.8,4),(1.2,3.4),(1.6,3.1),(2,2.9),(3,2.8))
Units: Dmnl

effect of proportion of deaths due to malaria on life expectancy= WITH LOOKUP (

Proportion Of Deaths Due To Malaria,
([(0,0.5)-
(0.9,1)],(0,1),(0.007,0.997),(0.011,0.995),(0.014,0.994),(0.069,0.97),(0.172,0.923),(0.184,0.918),(0.207,0.908),(0.31,0.859),(0.345,0.842),(0.368,0.83),(0.483,0.77),(0.828,0.516) ))
Units: Dmnl

EFFECT OF WELLBEING INDEX ON AVERAGE LIFE EXPECTANCY(
[(0,0)-
(1,100)],(0,32),(0.04,34.5),(0.08,42.5),(0.12,49),(0.16,55),(0.24,63),(0.32,68),(0.4,71),(0.48,73),(0.56,74),(0.64,74.5),(1,75))
Units: Year

ELASTICITY OF CONTRACEPTIVE PREVALENCE TO LITERACY RATE=
2.4
Units: Dmnl

elderly death rate= WITH LOOKUP (
average life expectancy 0,
([(0,0)-
(80,1)],(0,1),(20,0.410881),(22.5,0.389394),(25,0.370157),(27.5,0.352756),(30,0.336902),(32.5,0.322367),(35,0.308965),(37.5,0.296544),(40,0.284989),(42.5,0.274432),(45,0.265855),(47.5,0.257306),(50,0.248818),(52.5,0.240404),(55,0.232093),(57.5,0.223906),(60,0.215861),(62.5,0.208031),(65,0.199094),(67.5,0.188776),(70,0.1778),(72.5,0.166073),(75,0.15349),(77.5,0.139955),(80,0.125409) ))
Units: Dmnl/Year

ELDERLY DEATH RATE ADJUSTMENT FACTOR=
0
Units: Dmnl/Year

elderly deaths=
Elderly Population*(elderly death rate+ELDERLY DEATH RATE ADJUSTMENT FACTOR)
Units: Person/Year

elderly net migration=
Elderly Population*NET migration rate
Units: Person/Year

Elderly Population= INTEG (
becoming elderly+elderly net migration-elderly deaths,
ELDERLY population data)
Units: Person

ELDERLY population data
Units: Person

FERTILE PERIOD=
30
Units: Year

HEALTH CARE WEIGHT=
0.15
Units: Dmnl [0,1]

INCOME WEIGHT=
0.7
Units: Dmnl [0.5,1]

infant death rate= WITH LOOKUP (
average life expectancy 0,
([(0,0)-
(80,1)],(0,1),(20,0.12662),(22.5,0.113627),(25,0.103322),(27.5,0.0954962),(30,0.086664),(32.5,0.078377),(35,0.0722173),(37.5,0.0664472),(40,0.0593487),(42.5,0.0541115),(45,0.0490303),(47.5,0.0442555),(50,0.0397517),(52.5,0.0354835),(55,0.0314233),(57.5,0.0275458),(60,0.0238273),(62.5,0.0219045),(65,0.0186952),(67.5,0.0157407),(70,0.0129822),(72.5,0.0104157),(75,0.008036),(77.5,0.005832),(80,0.00378833) ))
Units: Dmnl/Year

INFANT DEATH RATE ADJUSTMENT FACTOR=
0
Units: Dmnl/Year

infant deaths=
Infant Population*(infant death rate+INFANT DEATH RATE ADJUSTMENT FACTOR)
Units: Person/Year

infant net migration=
Infant Population*NET migration rate
Units: Person/Year

Infant Population= INTEG (
births+infant net migration-becoming school age-infant deaths,
INFANT population data)
Units: Person

INFANT population data
Units: Person

INFANT STAGE DURATION=
6
Units: Year

INITIAL AVERAGE ADULT LITERACY RATE= INITIAL(
average adult literacy rate)
Units: Dmnl

INITIAL CONTRACEPTIVE PREVALENCE=
0.16

Units: Dmnl

INITIAL PC REAL DISPOSABLE INCOME= INITIAL(
PC real disposable income)
Units: Ksh05/(Year*Person)

INITIAL PERCEIVED PC REAL DISPOSABLE INCOME= INITIAL(
Perceived Pc Real Disposable Income)
Units: Ksh05/(Person*Year)

INITIAL PROPORTION OF DEATHS DUE TO MALARIA=
0.18
Units: Dmnl [0.1,0.3]

NATURAL fertility rate:INTERPOLATE:
Units: Dmnl

NET migration rate
Units: Dmnl/Year

PC real disposable income:INTERPOLATE:
Units: Ksh05/(Person*Year)

Perceived Pc Real Disposable Income=
SMOOTH N(pc real disposable income 0,
TIME FOR INCOME TO AFFECT FERTILITY AND MORTALITY,
INITIAL PC REAL DISPOSABLE INCOME, 1)
Units: Ksh05/(Person*Year)

PROPORTION OF CHILDBEARING FEMALE IN WORKING AGE POPULATION=
0.47
Units: Dmnl

Proportion Of Deaths Due To Malaria=
SMOOTH N(total malaria deaths/deaths, TIME STEP, INITIAL PROPORTION OF DEATHS DUE TO MALARIA, 1)
Units: Dmnl

REFERENCE SATURATION INCOME=
250000
Units: Ksh05/(Person*Year)

relative average adult literacy rate=
average adult literacy rate/INITIAL AVERAGE ADULT LITERACY RATE
Units: Dmnl

relative perceived pc real disposable income=
Perceived Pc Real Disposable Income/INITIAL PERCEIVED PC REAL DISPOSABLE INCOME
Units: Dmnl

school age death rate= WITH LOOKUP (
average life expectancy 0,
([(0,0)-
(80,1)],(0,1),(20,0.012801),(22.5,0.011582),(25,0.0104877),(27.5,0.00949433),(30,0.008588),(32.5,0.00775667),(35,0.00698867),(37.5,0.006277),(40,0.005614),(42.5,0.004957),(45,0.004342),(47.5,0.003779),(50,0.003262),(52.5,0.00278433),(55,0.00234067),(57.5,0.001931),(60,0.001546),(62.5,0.001189),(65,0.000933667),(67.5,0.000703333),(70,0.000508333),(72.5,0.00035),(75,0.000226667),(77.5,0.000136),(80,7.26667e-005) ))
Units: Dmnl/Year

SCHOOL AGE DEATH RATE ADJUSTMENT FACTOR=
0
Units: Dmnl/Year

school age deaths=
School Age Population*(school age death rate+SCHOOL AGE DEATH RATE ADJUSTMENT FACTOR)
Units: Person/Year

SCHOOL AGE DURATION=
9
Units: Year

school age net migration=
School Age Population*NET migration rate
Units: Person/Year

School Age Population= INTEG (
becoming school age+school age net migration-becoming working age-school age deaths,
SCHOOL age population data)
Units: Person

SCHOOL age population data
Units: Person

TIME FOR INCOME TO AFFECT FERTILITY AND MORTALITY=
10
Units: Year

total fertility rate=
desired fertility rate*EFFECT OF CONTRACEPTIVE PREVALENCE ON TOTAL FERTILITY RATE(contraceptive prevalence)
Units: Dmnl

wellbeing index=
MIN(1, Perceived Pc Real Disposable Income/REFERENCE SATURATION INCOME)*INCOME WEIGHT
+(average access to basic health care*HEALTH CARE WEIGHT)

+(average adult literacy rate*(1-(INCOME WEIGHT+HEALTH CARE WEIGHT)))
Units: Dmnl

working age death rate= WITH LOOKUP (
average life expectancy 0,
([(0,0)-
(80,1)],(0,1),(20,0.013531),(22.5,0.012284),(25,0.011165),(27.5,0.01015),(30,0.009223),(32
.5,0.008369),(35,0.007584),(37.5,0.006853),(40,0.006174),(42.5,0.005598),(45,0.004943),(4
7.5,0.004339),(50,0.003783),(52.5,0.003269),(55,0.002794),(57.5,0.002351),(60,0.001937),(
62.5,0.001549),(65,0.001173),(67.5,0.000896),(70,0.000659),(72.5,0.000462),(75,0.000306)
,(77.5,0.000186),(80,0.000102) ))
Units: Dmnl/Year

WORKING AGE DEATH RATE ADJUSTMENT FACTOR=
0
Units: Dmnl/Year [-0.02,0.02]

working age deaths=
Working Age Population*(working age death rate+WORKING AGE DEATH RATE ADJUSTMENT FACTOR)
Units: Person/Year

WORKING AGE DURATION=
50
Units: Year

working age net migration=
Working Age Population*NET migration rate
Units: Person/Year

Working Age Population= INTEG (
becoming working age+working age net migration-becoming elderly-working age deaths,
WORKING age population data)
Units: Person

WORKING age population data
Units: Person

*******************************
2 Education
*******************************

ADJUSTMENT INITIAL LITERATE ELDERLY POPULATION=
0
Units: Person

ADJUSTMENT INITIAL LITERATE WORKING AGE POPULATION=
0
Units: Person

ADULT literacy rate
Units: Dmnl

average adult literacy rate=
MIN(1,(Literate Working Age Population+Literate Elderly Population)/(Elderly Population+Working Age Population))
Units: Dmnl

AVERAGE COST PER STUDENT=
8900
Units: Ksh05/(Year*Person)

becoming literate elderly=
Literate Working Age Population/WORKING AGE DURATION
Units: Person/Year

becoming literate working age=
(School Age Population To Complete Primary Education-scholars affected by malaria symptoms)/SCHOOL AGE DURATION
Units: Person/Year

dropout=
School Age Population To Complete Primary Education*dropout fraction
Units: Person/Year

dropout fraction= WITH LOOKUP (
Time,
([(1999,0)-(2016,0.04)],(1999,0.032),(2016,0.01) ))
Units: Dmnl/Year

EDUCATION expenditure as a fraction of gdp
Units: Dmnl

initial literate elderly population=
ADULT literacy rate*Elderly Population+ADJUSTMENT INITIAL LITERATE ELDERLY POPULATION
Units: Person

INITIAL LITERATE SCHOOL AGE POPULATION=
3.5e+006
Units: Person

initial literate working age population=
Working Age Population*ADULT literacy rate+ADJUSTMENT INITIAL LITERATE WORKING AGE POPULATION
Units: Person

INITIAL PUBLIC SPENDING ON EDUCATION PER CAPITA= INITIAL(
public spending on education per capita)

Units: Ksh05/(Year*Person)

literate elderly deaths=
Literate Elderly Population*elderly death rate
Units: Person/Year

Literate Elderly Population= INTEG (
becoming literate elderly-literate elderly deaths,
initial literate elderly population)
Units: Person

literate school age deaths=
School Age Population To Complete Primary Education*school age death rate
Units: Person/Year

literate working age deaths=
Literate Working Age Population*working age death rate
Units: Person/Year

Literate Working Age Population= INTEG (
becoming literate working age-becoming literate elderly-literate working age deaths,
initial literate working age population)
Units: Person

percentage of school age population already in school=
School Age Population To Complete Primary Education/School Age Population
Units: Dmnl

percentage of school age population covered=
MIN(1,public spending on education/total expenditure to cover primary school)
Units: Dmnl

percentage of school age population going to school=
MIN(percentage of school age population covered,proportion of school age population willing to go to school)
Units: Dmnl

proportion of school age population willing to go to school= WITH LOOKUP (
pc real disposable income 0,
([(10000,0.4)-(35000,1)],(10000,0.5),(15000,0.7),(20000,0.85),(25000,0.95),(30000,1),(35000,1) ))
Units: Dmnl

public spending on education=
EDUCATION expenditure as a fraction of gdp*real gdp 0
Units: Ksh05/Year

public spending on education per capita=
public spending on education/School Age Population
Units: Ksh05/(Year*Person)

relative public spending on education per capita=
public spending on education per capita/INITIAL PUBLIC SPENDING ON EDUCATION PER CAPITA
Units: Dmnl

scholars affected by malaria symptoms=
population fraction affected by malaria symptoms*School Age Population To Complete Primary Education
Units: Person

School Age Population= INTEG (
becoming school age+school age net migration-becoming working age-school age deaths,
SCHOOL age population data)
Units: Person

SCHOOL age population data
Units: Person

School Age Population To Complete Primary Education= INTEG (
school entrance rate-becoming literate working age-dropout-literate school age deaths,
INITIAL LITERATE SCHOOL AGE POPULATION)
Units: Person

school enrollment gap=
percentage of school age population going to school-percentage of school age population already in school
Units: Dmnl

school entrance rate=
School Age Population*school enrollment gap/TIME TO ENROLL STUDENTS
+becoming literate working age
+dropout
+literate school age deaths
Units: Person/Year

TIME TO ENROLL STUDENTS=
1
Units: Year

total expenditure to cover primary school=
AVERAGE COST PER STUDENT*School Age Population
Units: Ksh05/Year

*******************************
3 Healthcare
*******************************

annual basic health expenditure=
share of health expenditure for basic healthcare*real health expenditure
Units: Ksh05/Year

average access to basic health care=
number of doctors per capita*RELATIONSHIP BETWEEN ACCESS TO BASIC HEALTH CARE AND DOCTORS PER CAPITA
Units: Dmnl

AVERAGE NUMBER OF DOCTORS PER HEALTH FACILITY=
1
Units: Doctor/Hospital

Basic Health Capital= INTEG (
annual basic health expenditure-basic health capital depreciation,
INITIAL BASIC HEALTH CAPITAL)
Units: Ksh05

basic health capital depreciation=
Basic Health Capital/DEPRECIATION TIME
Units: Ksh05/Year

DEPRECIATION TIME=
20
Units: Year

ELASTICITY OF NUMBER OF DOCTORS TO CAPITAL=
0.1
Units: Dmnl

HEALTH expenditure as fraction of gdp
Units: Dmnl

INITIAL BASIC HEALTH CAPITAL=
6.1e+010
Units: Ksh05

INITIAL NUMBER OF DOCTORS PC= INITIAL(
number of doctors per capita)
Units: Doctor/Person

INITIAL RESIDUAL BASIC HEALTH CAPITAL= INITIAL(
residual basic health capital)
Units: Ksh05

INVESTMENT COST PER HEALTH FACILITY=
8.9e+006
Units: Ksh05/Hospital

number of doctors=

total number of health facilities*AVERAGE NUMBER OF DOCTORS PER HEALTH FACILITY*(relative residual basic health capital^ELASTICITY OF NUMBER OF DOCTORS TO CAPITAL)
Units: Doctor

number of doctors per capita=
number of doctors/total population
Units: Doctor/Person

pc health expenditure=
real health expenditure/total population
Units: Ksh05/(Year*Person)

real health expenditure=
real gdp 0*HEALTH expenditure as fraction of gdp
Units: Ksh05/Year

RELATIONSHIP BETWEEN ACCESS TO BASIC HEALTH CARE AND DOCTORS PER CAPITA=
1000
Units: Dmnl/(Doctor/Person)

relative number of doctors pc=
number of doctors per capita/INITIAL NUMBER OF DOCTORS PC
Units: Dmnl

relative residual basic health capital=
residual basic health capital/INITIAL RESIDUAL BASIC HEALTH CAPITAL
Units: Dmnl

residual basic health capital=
Basic Health Capital*(1-share of basic health capital in health facilities)
Units: Ksh05

share of basic health capital in health facilities= WITH LOOKUP (
Time,
([(1980,0.1)-(2010,0.3)],(1980,0.285),(1985,0.234),(1990.15,0.2),(1995,0.18),(2000,0.165),(2005,0.16),(2010,0.16) ))
Units: Dmnl

share of health expenditure for basic healthcare= WITH LOOKUP (
Time,
([(1980,0.2)-(2015,0.8)],(1980,0.4),(1985,0.4),(1990,0.4),(1995,0.4),(2000,0.4),(2005,0.4),(2010,0.4),(2015,0.4) ))
Units: Dmnl

total number of health facilities=

Basic Health Capital*share of basic health capital in health facilities/INVESTMENT COST PER HEALTH FACILITY
Units: Hospital

********************************
4 Production
********************************

AVERAGE DEPRECIATION TIME=
20
Units: Year

Capital= INTEG (
real investment 0-depreciation,
INITIAL CAPITAL)
Units: Ksh05

capital share= WITH LOOKUP (
Time,
([(1980,0)-(2020,0.5)],(1980,0.35),(1990.03,0.35),(2000,0.35),(2010,0.35),(2019.76,0.35) ))
Units: Dmnl

depreciation=
Capital/AVERAGE DEPRECIATION TIME
Units: Ksh05/Year

DOMESTIC revenue
Units: Ksh05/Year

domestic revenue projected=
real gdp 0*TAX RATE PROJECTED
Units: Ksh05/Year

domestic revenue simulated=
IF THEN ELSE( Time<2010 , DOMESTIC revenue , domestic revenue projected )
Units: Ksh05/Year

effect of adult literacy rate on productivity=
relative adult literacy rate^ELASTICITY OF TFP TO ADULT LITERACY RATE
Units: Dmnl

effect of life expectancy on productivity=
relative life expectancy^ELASTICITY OF TFP TO LIFE EXPECTANCY
Units: Dmnl

ELASTICITY OF PROPENSITY TO SAVE TO INCOME=
0.6
Units: Dmnl

ELASTICITY OF TFP TO ADULT LITERACY RATE=
0.4
Units: Dmnl

ELASTICITY OF TFP TO LIFE EXPECTANCY=
0.7
Units: Dmnl

GDP growth
Units: Dmnl/Year

households revenue=
real gdp 0
+subsidies and transfers simulated
+interest on domestic debt simulated
+NET current transfers from abroad
Units: Ksh05/Year

INITIAL ADULT LITERACY RATE= INITIAL(
average adult literacy rate)
Units: Dmnl

INITIAL AVERAGE LIFE EXPECTANCY= INITIAL(
average life expectancy 0)
Units: Year

INITIAL CAPITAL=
3e+012
Units: Ksh05

INITIAL PC REAL DISPOSABLE INCOME 0= INITIAL(
pc real disposable income 0)
Units: Ksh05/(Person*Year)

INITIAL PRODUCTION=
6e+011
Units: Ksh05/Year

INITIAL PROPENSITY TO SAVE=
0.2
Units: Dmnl

INITIAL REAL GDP GROWTH RATE=
0.036
Units: Dmnl/Year

INITIAL WORKING FORCE= INITIAL(
working age population without malaria symptoms)
Units: Person

INTEREST on domestic debt:INTERPOLATE:
Units: Ksh05/Year

INTEREST ON DOMESTIC DEBT AS SHARE OF SAVING=
0.08
Units: Dmnl

Interest On Domestic Debt Projected=
DELAY N(saving*INTEREST ON DOMESTIC DEBT AS SHARE OF SAVING ,
LENDING TO GOVERNMENT DISBURSMENT TIME , INTEREST on domestic debt , 1)
Units: Ksh05/Year

interest on domestic debt simulated=
IF THEN ELSE( Time<2010 , INTEREST on domestic debt , Interest On Domestic Debt Projected )
Units: Ksh05/Year

labor productivity=
effect of life expectancy on productivity*effect of adult literacy rate on productivity
Units: Dmnl

LENDING TO GOVERNMENT DISBURSMENT TIME=
1
Units: Year

NET current transfers from abroad
Units: Ksh05/Year

pc real disposable income 0=
real disposable income/total population
Units: Ksh05/(Person*Year)

PRIVATE saving:INTERPOLATE:
Units: Ksh05/Year

propensity to save=
MIN(1, INITIAL PROPENSITY TO SAVE*relative pc real disposable income^ELASTICITY OF PROPENSITY TO SAVE TO INCOME)
Units: Dmnl

real disposable income=
households revenue-domestic revenue simulated
Units: Ksh05/Year

REAL gdp:INTERPOLATE:
Units: Ksh05/Year

real gdp 0=
INITIAL PRODUCTION*relative production

Units: Ksh05/Year

real gdp growth rate=
TREND(real gdp 0, TIME HORIZON TO MEASURE GROWTH RATE, INITIAL REAL GDP GROWTH RATE)
Units: Dmnl/Year

REAL gross capital formation
Units: Usd05/Year

REAL investment:INTERPOLATE:
Units: Ksh05/Year

real investment 0=
saving
Units: Ksh05/Year

real pc gdp=
real gdp 0/total population
Units: Ksh05/(Person*Year)

relative adult literacy rate=
average adult literacy rate/INITIAL ADULT LITERACY RATE
Units: Dmnl

relative capital=
Capital/INITIAL CAPITAL
Units: Dmnl

relative life expectancy=
average life expectancy 0/INITIAL AVERAGE LIFE EXPECTANCY
Units: Dmnl

relative pc real disposable income=
pc real disposable income 0/INITIAL PC REAL DISPOSABLE INCOME 0
Units: Dmnl

relative production=
(relative capital^capital share)*(relative working force^(1-capital share))*labor productivity
Units: Dmnl

relative working force=
working age population without malaria symptoms/INITIAL WORKING FORCE
Units: Dmnl

saving=
real disposable income*propensity to save
Units: Ksh05/Year

SUBSIDIES and transfers
Units: Ksh05/Year

SUBSIDIES AND TRANSFERS AS SHARE OF GDP=
0.01
Units: Dmnl

subsidies and transfers projected=
real gdp 0*SUBSIDIES AND TRANSFERS AS SHARE OF GDP
Units: Ksh05/Year

subsidies and transfers simulated=
IF THEN ELSE( Time<2004 , SUBSIDIES and transfers , subsidies and transfers projected )
Units: Ksh05/Year

TAX RATE PROJECTED=
0.22
Units: Dmnl

TIME HORIZON TO MEASURE GROWTH RATE=
1
Units: Year

working age population without malaria symptoms=
Working Age Population*(1-labor fraction lost to malaria)
Units: Person

*******************************
5 Malaria transmission
*******************************

area fraction covered by environmental management=
MIN(1,total area covered by environmental management/area of population living in malaria risk areas)
Units: Dmnl

area of population living in malaria risk areas=
IF THEN ELSE( Time<2018, TOTAL LAND AREA OF KENYA*CLIMATE SUITABILITY INDEX(Time),
TOTAL LAND AREA OF KENYA*PROJECTED CLIMATE SUITABILITY INDEX)
Units: Square kilometer

average duration of immunity and infectiveness=
REFERENCE AVERAGE DURATION OF IMMUNITY AND INFECTIVENESS*effect of case management coverage on duration fraction of infectiousness
Units: Year

CLIMATE SUITABILITY INDEX(
[(1980,0.7)-(2015,0.8)],(1980,0.75),(1990,0.75),(2000,0.75),(2010,0.75),(2015,0.75))
Units: Dmnl

deaths due to other causes=
Malaria Infected And Partially Immune Population/average life expectancy 0
Units: Person/Year

em coverage=
POPULATION DENSITY TABLE(area fraction covered by environmental management)*PROPORTIONAL REDUCTION IN RISK FOR EM COVERED POPULATION
Units: Dmnl

indicated ivm coverage fraction=
em coverage+protective measures effective coverage+irs effective coverage
Units: Dmnl

infected recovered=
Malaria Infected And Partially Immune Population/average duration of immunity and infectiveness
Units: Person/Year

INITIAL MALARIA PREVALENCE=
0.26
Units: Dmnl

integrated vector management coverage fraction=
MIN(1,
indicated ivm coverage fraction+
(MAX(em coverage,MAX(protective measures effective coverage,irs effective coverage))-indicated ivm coverage fraction)
*INTERVENTIONS OVERLAPPING FACTOR)
Units: Dmnl

INTERVENTIONS OVERLAPPING FACTOR=
0.5
Units: Dmnl

malaria case fatality rate[age]=
NATURAL MALARIA CASE FATALITY RATE[age]*effect of case management coverage on duration fraction of malaria mortality
Units: Dmnl

malaria cases=
Malaria Infected And Partially Immune Population
*MALARIA CONTACT RATE*MALARIA INFECTIVITY*(non infected vulnerable population/vulnerable population)
Units: Person/Year

malaria cases old model= WITH LOOKUP (
Time,
([(1980,0)-
(2030,3e+007)],(1980,1.22382e+007),(1981,1.26341e+007),(1982,1.30791e+007),(1983,1.3541e+007),(1984,1.40142e+007),(1985,1.44951e+007),(1986,1.49873e+007),(1987,1.54943e+007),(1988,1.60169e+007),(1989,1.65557e+007),(1990,1.71184e+007),(1991,1.77009e+007),(1992,1.82922e+007),(1993,1.88864e+007),(1994,1.94805e+007),(1995,2.00738e+007),(1996,2.06683e+007),(1997,2.12716e+007),(1998,2.18849e+007),(1999,2.25089e+007),(2000,2.31437e+007),(2001,2.36985e+007),(2002,2.28185e+007),(2003,2.1965e+007),(2004,2.26436e+007),(2005,2.22162e+007),(2006,1.94397e+007),(2007,1.61244e+007),(2008,1.48272e+007),(2009,1.3866e+007),(2010,1.30115e+007),(2011,1.16931e+007),(2012,1.0016e+007),(2013,9.73755e+006),(2014,9.78901e+006),(2015,9.15157e+006),(2016,7.89278e+006),(2017,7.50722e+006),(2018,6.89609e+006),(2019,6.4049e+006),(2020,6.0582e+006),(2021,5.80961e+006),(2022,5.63034e+006),(2023,5.49952e+006),(2024,5.41458e+006),(2025,5.38033e+006),(2026,5.38182e+006),(2027,5.40834e+006),(2028,5.45292e+006),(2029,5.51083e+006),(2030,5.57875e+006) ))
Units: Person/Year

MALARIA CONTACT RATE=
3
Units: Dmnl/Year

malaria deaths[UNDER FIVE]=
malaria cases*(Infant Population/total population)*malaria case fatality rate[UNDER FIVE]
malaria deaths[FIVE AND OVER]=
malaria cases*((total population-Infant Population)/total population)*malaria case fatality rate[FIVE AND OVER]
Units: Person/Year

malaria incidence=
malaria cases
Units: Person/Year

Malaria Infected And Partially Immune Population= INTEG (
malaria incidence-deaths due to other causes-infected recovered-total malaria deaths,
total population*INITIAL MALARIA PREVALENCE)
Units: Person

MALARIA INFECTIVITY=
1
Units: Dmnl

malaria mortality=
total malaria deaths/total population
Units: Dmnl/Year

malaria prevalence=
MAX(0,Malaria Infected And Partially Immune Population/total population)
Units: Dmnl

NATURAL MALARIA CASE FATALITY RATE[UNDER FIVE]=
0.065
NATURAL MALARIA CASE FATALITY RATE[FIVE AND OVER]=
0.003
Units: Dmnl

non infected vulnerable population=
MAX(0,vulnerable population-Malaria Infected And Partially Immune Population)
Units: Person

POPULATION at risk
Units: Person

POPULATION DENSITY TABLE(
[(0,0)-(1,1)],(0,0),(1.19e-006,0.000588),(3.26e-006,0.00153),(4.58e-006,0.00212),(7.12e-006,0.00314),(8.44e-006,0.00366),(1.07e-005,0.00454),(1.24e-005,0.00516),(1.48e-005,0.00601),(3.95e-005,0.0147),(4.83e-005,0.0177),(5.76e-005,0.0209),(6.73e-005,0.0242),(7.14e-005,0.0255),(8.51e-005,0.0298),(8.75e-005,0.0305),(9.11e-005,0.0315),(9.34e-005,0.0321),(9.44e-005,0.0324),(9.79e-005,0.0333),(0.000103,0.0346),(0.000105,0.0351),(0.000112,0.0368),(0.000115,0.0377),(0.000119,0.0386),(0.000126,0.0402),(0.000134,0.0421),(0.000137,0.0428),(0.000179,0.0526),(0.000184,0.0535),(0.000204,0.0582),(0.000207,0.0588),(0.000216,0.0604),(0.000223,0.0619),(0.000233,0.0637),(0.00024,0.0652),(0.000241,0.0652),(0.000261,0.0687),(0.000267,0.0699),(0.000277,0.0714),(0.00029,0.0736),(0.000305,0.076),(0.000308,0.0765),(0.000311,0.0769),(0.000319,0.0782),(0.000323,0.0789),(0.000327,0.0794),(0.000361,0.0847),(0.000365,0.0854),(0.000383,0.088),(0.000387,0.0885),(0.000396,0.0898),(0.000403,0.0907),(0.000406,0.0911),(0.000409,0.0915),(0.000412,0.0918),(0.000426,0.0935),(0.000431,0.0941),(0.00046,0.0974),(0.000466,0.098),(0.000482,0.0996),(0.000487,0.1),(0.000526,0.104),(0.000561,0.107),(0.000567,0.108),(0.000573,0.108),(0.000598,0.111),(0.000619,0.112),(0.000627,0.113),(0.000734,0.121),(0.000735,0.121),(0.000738,0.121),(0.000748,0.121),(0.000751,0.122),(0.000761,0.122),(0.000779,0.123),(0.000828,0.126),(0.000839,0.126),(0.00086,0.127),(0.00087,0.127),(0.001,0.133),(0.002,0.163),(0.003,0.183),(0.004,0.199),(0.005,0.213),(0.006,0.225),(0.007,0.235),(0.008,0.244),(0.009,0.253),(0.01,0.261),(0.02,0.319),(0.03,0.359),(0.04,0.391),(0.05,0.417),(0.1,0.511),(0.15,0.575),(0.2,0.625),(0.25,0.667),(0.3,0.704),(0.35,0.736),(0.4,0.765),(0.45,0.792),(0.5,0.817),(0.55,0.84),(0.6,0.861),(0.65,0.882),(0.7,0.901),(0.75,0.919),(0.8,0.937),(0.85,0.954),(0.9,0.97),(0.95,0.985),(1,1))
Units: Dmnl

population living in malaria risk areas=
proportion of population living in malaria risk areas*total population
Units: Person

PROJECTED CLIMATE SUITABILITY INDEX=
0.75
Units: Dmnl

proportion of malaria cases reported=
IF THEN ELSE(Time<1995, 0 , MIN(1, REPORTED malaria cases/malaria cases) )
Units: Dmnl

proportion of population living in malaria risk areas=
area of population living in malaria risk areas/TOTAL LAND AREA OF KENYA
Units: Dmnl

PROPORTIONAL REDUCTION IN RISK FOR EM COVERED POPULATION=
0.35
Units: Dmnl

REFERENCE AVERAGE DURATION OF IMMUNITY AND INFECTIVENESS=
0.531
Units: Year

REPORTED malaria cases
Units: Person/Year

REPORTED malaria deaths
Units: Person/Year

TOTAL LAND AREA OF KENYA=
581677
Units: Square kilometer

total malaria deaths=
SUM(malaria deaths[age!])
Units: Person/Year

total population=
Elderly Population+Infant Population+School Age Population+Working Age Population
Units: Person

under five malaria deaths=
malaria deaths[UNDER FIVE]
Units: Person/Year

under five malaria deaths as share of total deaths=
IF THEN ELSE(total malaria deaths<1, 0, under five malaria deaths/total malaria deaths)
Units: Dmnl

under six malaria cases=
malaria cases*Infant Population/total population
Units: Person/Year

vulnerable population=
MAX(1,population living in malaria risk areas*(1-integrated vector management coverage fraction))
Units: Person

******************************
6 IVM interventions
******************************

Area Covered By Larviciding=
DELAY N(larviciding expenditure/LARVICIDING UNIT COST, TIME FOR LARVICIDING IMPLEMENTATION,0,3)
Units: Square kilometer

Area Covered By Mosquito Source Reduction= INTEG (
Source Reduction-source regeneration,
INITIAL AREA COVERED BY SOURCE REDUCTION)
Units: Square kilometer

AVERAGE BEDNET EFFECTIVE DURATION=
3.3
Units: Year

AVERAGE DURATION OF SOURCE REDUCTION INTERVENTIONS=
8
Units: Year

bednet covered population=
Bednets In Households*COVERAGE PER BEDNET
Units: Person

BEDNET UNIT PRICE=
6
Units: $/Bednet

bednets delivered=
budget for bednets/BEDNET UNIT PRICE
Units: Bednet/Year

Bednets In Households= INTEG (
bednets delivered-discarded bednets,
INITIAL BEDNETS IN HOUSEHOLDS)
Units: Bednet

budget for bednets=
bednet expenditure*FRACTION OF PROTECTIVE MEASURES EXPENDITURE FOR BEDNETS
Units: $/Year

budget for non bednets protective measures=
bednet expenditure*(1-FRACTION OF PROTECTIVE MEASURES EXPENDITURE FOR BEDNETS)
Units: $/Year

COVERAGE PER BEDNET=

2
Units: Person/Bednet

ddt coverage=
Population Covered By Ddt Application
Units: Person

ddt expenditure=
irs expenditure*IF THEN ELSE(Time < 2017, SHARE OF IRS BUDGET FOR DDT(Time), FUTURE SHARE OF IRS BUDGET FOR DDT)
Units: $/Year

DDT MOSQUITO RESISTANCE=
0.4
Units: Dmnl

DDT PRICE PER PERSON PER YEAR=
4
Units: $/(Person*Year)

discarded bednets=
Bednets In Households/AVERAGE BEDNET EFFECTIVE DURATION
Units: Bednet/Year

effect of household sensitizing on protective measures=
MIN(1,efficacy of protective measures without sensitization+(MAXIMUM SENSITIZATION EFFECTIVENESS-efficacy of protective measures without sensitization)*(proportion of households in malaria risk areas sensitized/SENSITIZATION COVERAGE NECESSARY TO ACHIEVE MAXIMUM EFFECTIVENESS))
Units: Dmnl

EFFECT OF LITERACY RATE ON PROTECTIVE MEASURES EFFICACY(
[(0,0)-(1,1)],(0,0.15),(0.2,0.35),(0.4,0.48),(0.6,0.55),(0.8,0.59),(1,0.6))
Units: Dmnl

efficacy of protective measures without sensitization=
EFFECT OF LITERACY RATE ON PROTECTIVE MEASURES EFFICACY(average adult literacy rate)
Units: Dmnl

em expenditure=
integrated vector management interventions budget*normalized share of ivm budget for em
Units: $/Year

em expenditure for infrastructure and household modifications=
em expenditure*SHARE OF EM EXPENDITURE FOR INFRASTRUCTURE AND HOUSEHOLD MODIFICATIONS TIME SERIES(Time)
Units: $/Year

em unit cost=
IF THEN ELSE(Time<2017, ENVIRONMENTAL MANAGEMENT UNIT COST(Time), FUTURE EM UNIT COST)
Units: $/Square kilometer

ENVIRONMENTAL MANAGEMENT UNIT COST(
[(1980,2000)-(2017,5000)],(1980,3000),(2017,3000))
Units: $/Square kilometer

fraction of chemical control expenditure for non ddt irs=
1-IF THEN ELSE(Time < 2018, SHARE OF IRS BUDGET FOR DDT(Time), FUTURE SHARE OF IRS BUDGET FOR DDT)
Units: Dmnl

FRACTION of households owning at least one itn
Units: Dmnl

FRACTION OF PROTECTIVE MEASURES EXPENDITURE FOR BEDNETS=
1
Units: Dmnl

FUTURE EM UNIT COST=
3000
Units: $/Square kilometer

FUTURE SHARE OF IRS BUDGET FOR DDT=
0
Units: Dmnl

households forgetting awareness=
Sensitized Population/SENSITIZATION DURATION
Units: Person/Year

INITIAL AREA COVERED BY SOURCE REDUCTION=
1
Units: Square kilometer

INITIAL BEDNETS IN HOUSEHOLDS=
0
Units: Bednet

irs coverage=
people protected by irs/population living in malaria risk areas
Units: Dmnl

irs effective coverage=
MIN(1,PROPORTIONAL REDUCTION IN BITES WHEN USING IRS*(population efficiently covered by irs/population living in malaria risk areas))
Units: Dmnl

irs mosquito percentual resistance=
IF THEN ELSE(Time<2018, IRS mosquito resistance, PROJECTED IRS RESISTANCE)
Units: Dmnl

IRS mosquito resistance:INTERPOLATE:
Units: Dmnl

ITN coverage:INTERPOLATE:
Units: 1

itn coverage at risk=
MIN(1,bednet covered population/population living in malaria risk areas)
Units: 1

itn coverage total=
MIN(1,bednet covered population/total population)
Units: Dmnl

larviciding expenditure=
em expenditure-em expenditure for infrastructure and household modifications
Units: $/Year

LARVICIDING UNIT COST=
1500
Units: $/Square kilometer/Year

MAXIMUM SENSITIZATION EFFECTIVENESS=
1
Units: Dmnl

net use=
itn coverage total*effect of household sensitizing on protective measures
Units: Dmnl

net use of population at risk=
itn coverage at risk*effect of household sensitizing on protective measures
Units: Dmnl

NON BEDNETS PROTECTIVE MEASURES PRICE PER PERSON PER YEAR=
20
Units: $/(Year*Person)

non ddt irs coverage=
Population Covered By Non Ddt Irs Application
Units: Person

non ddt irs expenditure=
irs expenditure*fraction of chemical control expenditure for non ddt irs
Units: $/Year

NON DDT IRS UNIT COST=
6
Units: $/(Year*Person)

people protected by irs=
ddt coverage+non ddt irs coverage
Units: Person

Population Covered By Ddt Application=
DELAY N(ddt expenditure/DDT PRICE PER PERSON PER YEAR,TIME FOR IRS DEPLOYMENT,0,3)
Units: Person

Population Covered By Non Bednets Protective Measures=
DELAY N(budget for non bednets protective measures/NON BEDNETS PROTECTIVE MEASURES PRICE PER PERSON PER YEAR, TIME FOR NON BEDNET PROTECTIVE MEASURES IMPLEMENTATION,0,1)
Units: Person

Population Covered By Non Ddt Irs Application=
DELAY N(non ddt irs expenditure/NON DDT IRS UNIT COST, TIME FOR IRS DEPLOYMENT,0,3)
Units: Person

population efficiently covered by ddt application=
ddt coverage*(1-DDT MOSQUITO RESISTANCE)
Units: Person

population efficiently covered by irs=
(population efficiently covered by ddt application+population efficiently covered by non ddt irs application)
Units: Person

population efficiently covered by non ddt irs application=
non ddt irs coverage*(1-irs mosquito percentual resistance)
Units: Person

population fraction covered by irs=
people protected by irs/total population
Units: Dmnl

population fraction efficiently covered by bednets=
net use of population at risk*PROPORTIONAL REDUCTION IN BITES WHEN USING NETS
Units: Dmnl

population fraction efficiently covered by non bednets protective measures=
(Population Covered By Non Bednets Protective Measures/population living in malaria risk areas)*effect of household sensitizing on protective measures

Units: Dmnl

PROJECTED IRS RESISTANCE=
0.2
Units: Dmnl

projected irs resistance timeline= WITH LOOKUP (
Time,
([(2010,0)-(2030,1)],(2010,0.2),(2015,0.2),(2020,0.2),(2025,0.2),(2030,0.2) ))
Units: Dmnl

proportion of households in malaria risk areas sensitized=
MIN(1,Sensitized Population/population living in malaria risk areas)
Units: Dmnl

PROPORTIONAL REDUCTION IN BITES WHEN USING IRS=
0.7
Units: Dmnl

PROPORTIONAL REDUCTION IN BITES WHEN USING NETS=
0.75
Units: Dmnl

protective measures effective coverage=
MIN(1, (population fraction efficiently covered by bednets+population fraction efficiently covered by non bednets protective measures
))
Units: Dmnl

sensitization=
sensitization expenditure/SENSITIZATION PRICE PER PERSON PER YEAR
Units: Person/Year

SENSITIZATION COVERAGE NECESSARY TO ACHIEVE MAXIMUM EFFECTIVENESS=
1
Units: Dmnl

SENSITIZATION DURATION=
1
Units: Year

SENSITIZATION PRICE PER PERSON PER YEAR=
1
Units: $/Person

Sensitized Population= INTEG (
sensitization-households forgetting awareness,
0)
Units: Person

SHARE OF EM EXPENDITURE FOR INFRASTRUCTURE AND HOUSEHOLD MODIFICATIONS TIME SERIES(
[(1970,0)-(2050,1.2)],(1970,1),(2005,1),(2050,0.5))
Units: Dmnl

SHARE OF IRS BUDGET FOR DDT(
[(1990,0)-(2010,1)],(1990,0),(2010,0))
Units: Dmnl

Source Reduction=
DELAY N(em expenditure for infrastructure and household modifications/em unit cost,
TIME FOR EM IMPLEMENTATION, 0.555, 1)
Units: Square kilometer/Year

source regeneration=
Area Covered By Mosquito Source Reduction/AVERAGE DURATION OF SOURCE REDUCTION INTERVENTIONS
Units: Square kilometer/Year

TIME FOR EM IMPLEMENTATION=
3
Units: Year

TIME FOR IRS DEPLOYMENT=
0.16
Units: Year

TIME FOR LARVICIDING IMPLEMENTATION=
0.16
Units: Year

TIME FOR NON BEDNET PROTECTIVE MEASURES IMPLEMENTATION=
0.16
Units: Year

total area covered by environmental management=
Area Covered By Mosquito Source Reduction+Area Covered By Larviciding
Units: Square kilometer

*******************************
7 Case Management
*******************************

access to public malaria treatment facilities=
EFFECT OF ACCESS TO BASIC HEALTH CARE ON ACCESS TO MALARIA TREATMENTS(average access to basic health care)
Units: Dmnl

asymptomatic population fraction=
(average duration of immunity and infectiveness-TIME TO RECOVER SYMPTOMATIC)/(TIME TO RECOVER ASYMPTOMATIC-TIME TO RECOVER SYMPTOMATIC)
Units: Dmnl

average malaria treatment unit cost=
PROPORTION OF SEVERE MALARIA CASES*TREATMENT COST OF SEVERE MALARIA ANEMIA+(1-PROPORTION OF SEVERE MALARIA CASES)*TREATMENT COST OF UNCOMPLICATED MALARIA CASE
Units: $/Person

EFFECT OF ACCESS TO BASIC HEALTH CARE ON ACCESS TO MALARIA TREATMENTS(
[(0,0)-(0.5,1)],(0,0),(0.1,0.35),(0.2,0.64),(0.3,0.85),(0.4,0.95),(0.5,1))
Units: Dmnl

effect of case management coverage on duration fraction of infectiousness=
EFFECT OF CASE MANAGEMENT COVERAGE ON DURATION FRACTION OF INFECTIOUSNESS TABLE
(fraction of symptomatic malaria cases attending health centers or dispensaries)
Units: Dmnl

EFFECT OF CASE MANAGEMENT COVERAGE ON DURATION FRACTION OF INFECTIOUSNESS TABLE(
[(0,0.6)-(1,1)],(0,1),(0.1,0.99),(0.2,0.98),(0.3,0.96),(0.4,0.93),(0.5,0.88),(0.6,0.82),(0.7,0.76),(0.8,0.71),(0.9,0.68),(1,0.67))
Units: Dmnl

effect of case management coverage on duration fraction of malaria mortality=
EFFECT OF CASE MANAGEMENT COVERAGE ON DURATION FRACTION OF MALARIA MORTALITY TABLE
(fraction of symptomatic malaria cases attending health centers or dispensaries)
Units: Dmnl

EFFECT OF CASE MANAGEMENT COVERAGE ON DURATION FRACTION OF MALARIA MORTALITY TABLE(
[(0,0)-(1,0.4)],(0,0.37),(0.2,0.3),(0.4,0.24),(0.6,0.18),(0.8,0.14),(1,0.12))
Units: Dmnl

EFFECT OF EDUCATION LEVEL ON PROMPT ATTENDANCE TO HEALTH CARE SERVICES(
[(0,0)-(0.8,1)],(0,0.2),(0.2,0.6),(0.4,0.8),(0.6,0.95),(0.8,1))
Units: Dmnl

expenditure needed for malaria case management=
malaria cases*average malaria treatment unit cost*fraction of symptomatic cases
Units: $/Year

fraction of symptomatic cases=
(1-asymptomatic population fraction)
Units: Dmnl

fraction of symptomatic malaria cases attending health centers or dispensaries=
MIN(1, target malaria treatment coverage*
access to public malaria treatment facilities*
fraction of symptomatic malaria cases willing to attend health care services)
Units: Dmnl

fraction of symptomatic malaria cases willing to attend health care services=
EFFECT OF EDUCATION LEVEL ON PROMPT ATTENDANCE TO HEALTH CARE SERVICES(average adult literacy rate)
Units: Dmnl

malaria case management expenditure=
fraction of symptomatic malaria cases attending health centers or dispensaries
*expenditure needed for malaria case management
*proportion of treated cases attending public malaria services
Units: $/Year

PROPORTION OF SEVERE MALARIA CASES=
0.125
Units: Dmnl

proportion of treated cases attending public malaria services= WITH LOOKUP (
Time,
([(2000,0)-(2016,0.9)],(2000,0.2),(2002,0.24),(2004,0.34),(2005.95,0.48),(2007.88,0.59),(2010,0.65),(2012,0.67),(2014,0.68),(2016,0.68) ))
Units: Dmnl

target malaria treatment coverage= WITH LOOKUP (
Time,
([(2000,0)-(2011,1)],(2000,0.1),(2001.65,0.11),(2003.03,0.14),(2004.07,0.22),(2005.11,0.4),(2006.02,0.6),(2006.96,0.77),(2007.97,0.89),(2008.98,0.95),(2009.92,0.98),(2011,1) ))
Units: Dmnl

TIME TO RECOVER ASYMPTOMATIC=
1
Units: Year

TIME TO RECOVER SYMPTOMATIC=
0.06
Units: Year

TREATMENT COST OF SEVERE MALARIA ANEMIA=
39
Units: $/Person

TREATMENT COST OF UNCOMPLICATED MALARIA CASE=
2
Units: $/Person

*******************************
8 Malaria Cost Accounting
*******************************

Accumulated Malaria Expenditure= INTEG (
malaria control funding,
0)
Units: $

Accumulated Real Gdp In Constant Usd10= INTEG (
real gdp in constant usd10, 0)
Units: $

Accumulated Reported Financing= INTEG (
reported financing, 0)
Units: $

average malaria treatment unit cost=
PROPORTION OF SEVERE MALARIA CASES*TREATMENT COST OF SEVERE MALARIA ANEMIA+(1-PROPORTION OF SEVERE MALARIA CASES)*TREATMENT COST OF UNCOMPLICATED MALARIA CASE
Units: $/Person

bednet expenditure=
integrated vector management interventions budget*normalized share of ivm budget for bednet
Units: $/Year

effective malaria control expenditure=
total funding for malaria control-malaria planning administration overhead expendture
Units: $/Year

em expenditure=
integrated vector management interventions budget*normalized share of ivm budget for em
Units: $/Year

EXCHANGE rate:INTERPOLATE:
Units: Ksh/Current$

FUTURE SHARE OF IVM BUDGET FOR BEDNET=
0.75
Units: Dmnl

FUTURE SHARE OF IVM BUDGET FOR EM=
0.08
Units: Dmnl

FUTURE SHARE OF IVM BUDGET FOR IRS=
0.05
Units: Dmnl

FUTURE SHARE OF IVM BUDGET FOR SENSITIZATION=
0.12
Units: Dmnl

GDP deflator:INTERPOLATE:
Units: Ksh/Ksh05

GDP deflator usd10:INTERPOLATE:
Units: Current$/$

GOVERNMENT contribution:INTERPOLATE:
Units: $/Year

integrated vector management interventions budget=
malaria prevention expenditure
Units: $/Year

irs expenditure=
integrated vector management interventions budget*normalized share of ivm budget for irs
Units: $/Year

malaria control funding=
total funding for malaria control
Units: $/Year

MALARIA external funding:INTERPOLATE:
Units: $/Year

malaria future funding=
MALARIA FUTURE FUNDING PER CAPITA*total population
Units: $/Year

MALARIA FUTURE FUNDING PER CAPITA=
5
Units: $/(Person*Year)

malaria planning administration overhead expendture=
total funding for malaria control*MALARIA PLANNING ADMINISTRATION OVERHEAD EXPENDTURE SHARE
Units: $/Year

MALARIA PLANNING ADMINISTRATION OVERHEAD EXPENDTURE SHARE=
0.1
Units: Dmnl

malaria prevention expenditure=
MAX(0.001, effective malaria control expenditure-malaria case management expenditure)
Units: $/Year

malaria total funding= WITH LOOKUP (
Time,
([(1980,0)-(2017,2e+008)],(1980,2.35785e+006),(1981,2.35785e+006),(1982,2.35785e+006),(1983,2.35785e+006),(1984,2.35785e+006),(1985,2.35785e+006),(1986,2.35785e+006),(1987,2.35785e+006),(1988,2.35785e+006),(1989,2.35785e+006),(1990,2.35785e+006),(1991,2.35785e+006),(1992,2.35785e+006),(1993,2.35785e+006),(1994,2.35785e+006),(1995,2.35785e+006),(1996,2.35785e+006),(1997,2.35785e+006),(1998,2.35785e+006),(1999,2.35785e+006),(2000,3.558e+006),(2001,4.343e+006),(2002,4.63643e+007),(2003,7.876e+006),(2004,1.44659e+007),(2005,3.59495e+007),(2006,7.10062e+007),(2007,5.369e+007),(2008,5.777e+007),(2009,4.899e+007),(2010,4.007e+007),(2011,1.071e+008),(2012,6.673e+007),(2013,3.341e+007),(2014,6.64829e+007),(2015,1.20757e+008),(2016,3.76311e+007),(2017,1.35416e+008) ))
Units: $/Year

malaria total funding data=
MALARIA external funding+GOVERNMENT contribution
Units: $/Year

normalized share of ivm budget for bednet=
IF THEN ELSE(Time<2018,SHARE OF IVM BUDGET FOR BEDNET(Time),FUTURE SHARE OF IVM BUDGET FOR BEDNET)/total percentage of budget requested
Units: Dmnl

normalized share of ivm budget for em=
IF THEN ELSE(Time<2018, SHARE OF IVM BUDGET FOR EM(Time), FUTURE SHARE OF IVM BUDGET FOR EM)/total percentage of budget requested
Units: Dmnl

normalized share of ivm budget for irs=
IF THEN ELSE(Time<2018,SHARE OF IVM BUDGET FOR IRS(Time),FUTURE SHARE OF IVM BUDGET FOR IRS)/total percentage of budget requested
Units: Dmnl

normalized share of ivm budget for sensitization=
IF THEN ELSE(Time<2018, SHARE OF IVM BUDGET FOR SENSITIZATION(Time), FUTURE SHARE OF IVM BUDGET FOR SENSITIZATION)/total percentage of budget requested
Units: Dmnl

real gdp in constant usd10=
((real gdp 0*GDP deflator)/EXCHANGE rate)/GDP deflator usd10
Units: $/Year

real share bednets=
bednet expenditure/total ivm expenditure
Units: 1

real share em=
em expenditure/total ivm expenditure
Units: 1

real share irs=
irs expenditure/total ivm expenditure
Units: 1

real share sensitization=
sensitization expenditure/total ivm expenditure
Units: 1

reported financing=
IF THEN ELSE( Time<2018, malaria total funding data, 0)
Units: $/Year

sensitization expenditure=
integrated vector management interventions budget*normalized share of ivm budget for sensitization
Units: $/Year

SHARE OF IVM BUDGET FOR BEDNET(
[(2000,0)-(2018,0.9)],(2000,0.285731),(2001,0.230815),(2002,0.807698),(2003,0.657351),(2004,0.638146),(2005,0.757885),(2006,0.741168),(2007,0.310637),(2008,0.426357),(2009,0.558227),(2010,0.362029),(2011,0.685772),(2012,0.580449),(2013,0.731442),(2014,0.774134),(2015,0.755002),(2016,0.638864),(2017,0.839684),(2018,0.541736))
Units: Dmnl

SHARE OF IVM BUDGET FOR EM(
[(2000,0)-(2018,0.3)],(2000,3.95532e-008),(2001,0.12813),(2002,0.0990982),(2003,0.0240381),(2004,0.132114),(2005,0.0971399),(2006,0.134293),(2007,0.100832),(2008,0.0701191),(2009,0.0947891),(2010,0.120189),(2011,0.11702),(2012,0.056671),(2013,0.175147),(2014,0.147304),(2015,0.159781),(2016,0.235523),(2017,0.0913983),(2018,0.262455))
Units: Dmnl

SHARE OF IVM BUDGET FOR IRS(
[(2000,0)-(2018,0.8)],(2000,0.714269),(2001,0.576989),(2002,0.0436549),(2003,0.306592),(2004,0.163683),(2005,0.096405),(2006,0.0573922),(2007,0.538116),(2008,0.468464),(2009,0.29959),

(2010,0.457687),(2011,0.138698),(2012,0.334545),(2013,0),(2014,0),(2015,0),(2016,0),(2017,0.0201723),(2018,0.0558326))
Units: Dmnl

SHARE OF IVM BUDGET FOR SENSITIZATION(
[(2000,0)-(2018,0.2)],(2000,1.97766e-008),(2001,0.0640652),(2002,0.0495491),(2003,0.0120191),(2004,0.0660569),(2005,0.04857),(2006,0.0671466),(2007,0.050416),(2008,0.0350595),(2009,0.0473946),(2010,0.0600947),(2011,0.0585102),(2012,0.0283355),(2013,0.0934115),(2014,0.078562),(2015,0.0852165),(2016,0.125612),(2017,0.0487457),(2018,0.139976))
Units: Dmnl

share of total malaria funding for prevention=
malaria prevention expenditure/total funding for malaria control
Units: Dmnl

total funding for malaria control=
IF THEN ELSE(Time<2018,
IF THEN ELSE(Time<2000,
malaria case management expenditure/(1-MALARIA PLANNING ADMINISTRATION OVERHEAD EXPENDTURE SHARE),
malaria total funding),
malaria future funding)
Units: $/Year

total ivm expenditure=
bednet expenditure+em expenditure+irs expenditure+sensitization expenditure
Units: $/Year

total malaria expenditure=
(malaria case management expenditure+total ivm expenditure)/
(1-MALARIA PLANNING ADMINISTRATION OVERHEAD EXPENDTURE SHARE)
Units: $/Year

total percentage of budget requested=
IF THEN ELSE(Time<2018, SHARE OF IVM BUDGET FOR SENSITIZATION(Time)+SHARE OF IVM BUDGET FOR EM(Time)+SHARE OF IVM BUDGET FOR BEDNET(Time)+SHARE OF IVM BUDGET FOR IRS(Time), FUTURE SHARE OF IVM BUDGET FOR SENSITIZATION+FUTURE SHARE OF IVM BUDGET FOR EM+FUTURE SHARE OF IVM BUDGET FOR BEDNET+FUTURE SHARE OF IVM BUDGET FOR IRS)
Units: Dmnl

total population=
Elderly Population+Infant Population+School Age Population+Working Age Population
Units: Person

*******************************

9 Malaria-free Impacts
******************************

asymptomatic infected population=
Malaria Infected And Partially Immune Population*asymptomatic population fraction
Units: Person

average adult literacy rate without malaria=
MIN(1,(Literate Working Age Population Without Malaria+Literate Elderly Population Without Malaria)/(Elderly Population+Working Age Population))
Units: Dmnl

becoming literate elderly without malaria=
Literate Working Age Population Without Malaria/WORKING AGE DURATION
Units: Person/Year

becoming literate working age without malaria=
(School Age Population Without Malaria)/SCHOOL AGE DURATION
Units: Person/Year

dropout without malaria=
School Age Population Without Malaria*dropout fraction
Units: Person/Year

effect of adult literacy rate on tfp without malaria=
relative adult literacy rate without malaria^ELASTICITY OF TFP TO ADULT LITERACY RATE
Units: Dmnl

effect of life expectancy on tfp without malaria=
relative life expectancy without malaria^ELASTICITY OF TFP TO LIFE EXPECTANCY
Units: Dmnl

elderly death rate without malaria= WITH LOOKUP (
average life expectancy without malaria,
([(0,0)-(80,1)],(0,1),(20,0.410881),(22.5,0.389394),(25,0.370157),(27.5,0.352756),(30,0.336902),(32.5,0.322367),(35,0.308965),(37.5,0.296544),(40,0.284989),(42.5,0.274432),(45,0.265855),(47.5,0.257306),(50,0.248818),(52.5,0.240404),(55,0.232093),(57.5,0.223906),(60,0.215861),(62.5,0.208031),(65,0.199094),(67.5,0.188776),(70,0.1778),(72.5,0.166073),(75,0.15349),(77.5,0.139955),(80,0.125409) ))
Units: Dmnl/Year

equivalent labor force lost to malaria=
labor force*labor fraction lost to malaria
Units: Person

equivalent working age population lost to malaria=

FRACTION OF WORKING TIME LOSS BECAUSE OF MALARIA*population fraction affected by malaria symptoms*Working Age Population
Units: Person

fraction of total factor productivity lost to malaria=
(total factor productivity without malaria-labor productivity)/total factor productivity without malaria
Units: Dmnl

FRACTION OF WORKING TIME LOSS BECAUSE OF MALARIA=
0.6
Units: Dmnl

fractional gdp lost to malaria=
1-(relative production/relative production without malaria)
Units: Dmnl

labor force=
Working Age Population*LABOR force participation rate
Units: Person

LABOR force participation rate:INTERPOLATE:
Units: Dmnl

labor fraction lost to malaria=
equivalent working age population lost to malaria/Working Age Population
Units: Dmnl

literate elderly deaths without malaria=
Literate Elderly Population Without Malaria*elderly death rate without malaria
Units: Person/Year

Literate Elderly Population Without Malaria= INTEG (
becoming literate elderly without malaria-literate elderly deaths without malaria,
initial literate elderly population)
Units: Person

literate school age deaths without malaria=
School Age Population Without Malaria*school age death rate without malaria
Units: Person/Year

literate working age deaths without malaria=
Literate Working Age Population Without Malaria*working age death rate without malaria
Units: Person/Year

Literate Working Age Population Without Malaria= INTEG (
becoming literate working age without malaria-becoming literate elderly without malaria-literate working age deaths without malaria,
initial literate working age population)

Units: Person

population affected by malaria symptoms=
Malaria Infected And Partially Immune Population-asymptomatic infected population
Units: Person

population fraction affected by malaria symptoms=
MAX(0,population affected by malaria symptoms/total population)
Units: Dmnl

relative adult literacy rate without malaria=
average adult literacy rate without malaria/INITIAL ADULT LITERACY RATE
Units: Dmnl

relative gdp lost from labor=
1-((1-labor fraction lost to malaria)^(1-capital share))
Units: Dmnl

relative gdp lost from labor and total productivity factor=
1-((1-labor fraction lost to malaria)^(1-capital share)*(1-fraction of total factor productivity lost to malaria))
Units: Dmnl

relative life expectancy without malaria=
average life expectancy without malaria/INITIAL AVERAGE LIFE EXPECTANCY
Units: Dmnl

relative production without malaria=
(relative capital^capital share)*(relative working force without malaria^(1-capital share))
*total factor productivity without malaria
Units: Dmnl

relative working force without malaria=
Working Age Population/INITIAL WORKING FORCE
Units: Dmnl

school age death rate without malaria= WITH LOOKUP (
average life expectancy without malaria,
([(0,0)-(80,1)],(0,1),(20,0.012801),(22.5,0.011582),(25,0.0104877),(27.5,0.00949433),(30,0.008588),(32.5,0.00775667),(35,0.00698867),(37.5,0.006277),(40,0.005614),(42.5,0.004957),(45,0.004342),(47.5,0.003779),(50,0.003262),(52.5,0.00278433),(55,0.00234067),(57.5,0.001931),(60,0.001546),(62.5,0.001189),(65,0.000933667),(67.5,0.000703333),(70,0.000508333),(72.5,0.00035),(75,0.000226667),(77.5,0.000136),(80,7.26667e-005) ))
Units: Dmnl/Year

School Age Population Without Malaria= INTEG (
school entrance rate-becoming literate working age without malaria-dropout without malaria-literate school age deaths without malaria,

INITIAL LITERATE SCHOOL AGE POPULATION)
Units: Person

total factor productivity without malaria=
effect of life expectancy on tfp without malaria*effect of adult literacy rate on tfp without malaria
Units: Dmnl

working age death rate without malaria= WITH LOOKUP (
average life expectancy without malaria,
([(0,0)-(80,1)],(0,1),(20,0.013531),(22.5,0.012284),(25,0.011165),(27.5,0.01015),(30,0.009223),(32.5,0.008369),(35,0.007584),(37.5,0.006853),(40,0.006174),(42.5,0.005598),(45,0.004943),(47.5,0.004339),(50,0.003783),(52.5,0.003269),(55,0.002794),(57.5,0.002351),(60,0.001937),(62.5,0.001549),(65,0.001173),(67.5,0.000896),(70,0.000659),(72.5,0.000462),(75,0.000306),(77.5,0.000186),(80,0.000102) ))
Units: Dmnl/Year

working days lost to malaria=
equivalent labor force lost to malaria*WORKING DAYS PER HEALTHY WORKER
Units: Days/Year

WORKING DAYS PER HEALTHY WORKER=
230
Units: Days/(Person*Year)
